# Quantifying and Realizing the Benefits of Targeting for Pandemic Response

**DOI:** 10.1101/2021.03.23.21254155

**Authors:** Sergio Camelo, Dragos Florin Ciocan, Dan A. Iancu, Xavier S. Warnes, Spyros I. Zoumpoulis

**Affiliations:** Stanford University; INSEAD

**Keywords:** Pandemic management, Targeted interventions, Curfews, Optimization, COVID-19

## Abstract

To respond to pandemics such as COVID-19, policy makers have relied on interventions that target specific population groups or activities. Because targeting is operationally challenging and contentious, rigorously quantifying its benefits and designing practically implementable policies that achieve some of these benefits is critical for effective and equitable pandemic control. We propose a flexible framework that leverages publicly available data and a novel optimization algorithm based on model predictive control and trust region methods to compute optimized interventions that can target two dimensions of heterogeneity: age groups and the specific activities that individuals normally engage in. We showcase a complete implementation focused on the Île-de-France region of France and use this case study to quantify the benefits of dual targeting and to propose practically implementable policies. We find that dual targeting can lead to Pareto improvements, reducing the number of deaths and the economic losses. Additionally, dual targeting allows maintaining higher activity levels for most age groups and, importantly, for those groups that are most confined, thus leading to confinements that are arguably more equitable. We then fit decision trees to explain the decisions and gains of dual-targeted policies and find that they prioritize confinements intuitively, by allowing increased activity levels for group-activity pairs with high marginal economic value prorated by social contacts, which generates important complementarities. Because dual targeting can face significant implementation challenges, we introduce two practical proposals inspired by real-world interventions — based on curfews and recommendations — that achieve a significant portion of the benefits without explicitly discriminating based on age.

## 1. Introduction

The COVID-19 pandemic has forced policy makers worldwide to rely on a range of large-scale population confinement measures in an effort to contain disease spread. In determining these measures, a key recognition has been that substantial differences exist in the health and economic impact produced by different individuals engaged in distinct activities. Targeting confinements to account for such heterogeneity could be an important lever to mitigate a pandemic’s impact, but could also lead to potentially contentious and discriminatory measures. This work is aimed at developing a rigorous framework to quantify the benefits and downsides of such targeted interventions in pandemic management, and applying it to the COVID-19 pandemic as a real-world case study.

Targeting has been implemented in several different ways during the COVID-19 pandemic. We include a detailed survey of COVID-19 targeted restrictions in Appendix Table EC.14. One real-world contentious example has been to differentiate confinements based on *age groups*, e.g., sheltering older individuals who might face higher health risks if infected, or restricting younger groups who might create higher infection risks. Such measures have been implemented in several settings – e.g., with stricter confinements applied to older groups in Finland (Tiirinki et al. 2020), Ireland (Harrison 2020), Israel (Magid 2020) and Moscow (Foy 2020), or curfews applied to children and youth in Bosnia and Herzegovina (Reuters Staff 2020) and Turkey (Kanbur and Ankgül 2020) – but some of the measures were deemed ageist and unconstitutional and were eventually overturned (Magid 2020, Reuters Staff 2020).

A different example of targeting extensively employed in practice has been to tailor confinements to specific *activities* conducted during a typical day. This has been driven by the recognition that different activities (or more specifically, population interactions in locations of certain activities) such as work, schooling, transport or leisure can result in significantly different patterns of social contacts and new infections. This heterogeneity has been recognized in numerous implementations that differentially confine activities through restrictions of varying degrees on schools, workplaces, recreation venues, retail spaces, etc. Additionally, some practical implementations even differentiated based on *both* age groups and activities, e.g., by setting aside dedicated hours when only the senior population was allowed to shop at supermarkets (Aguilera 2020), or by restricting only higher age groups from in-person work activities (Magid 2020).

As these examples suggest, targeted interventions have merits but also pose potentially significant downsides. On the one hand, targeting can generate improvements in both health and economic outcomes, giving policy makers an improved lever when navigating difficult trade-offs. Additionally, explicitly considering *multiple* dimensions of targeting simultaneously – such as activities *and* age groups – could overturn some of the prevailing insight that specific age groups should uniformly face stricter confinements. However, such granular policies are more difficult to implement, and could lead to discriminatory and potentially unfair measures.

Given that some amount of targeting is already in place in existing real-world policy implementations, it is critical to transparently model and quantify its benefits and downsides, as well as examining practical policies that can realize some of these benefits. This gives rise to several natural research questions: How large are the health and economic benefits of interventions that can engage in progressively finer targeting? Does finer targeting lead to significant synergies, and is there an interpretable mechanism through which this happens? Because finer targeting may be impractical, are there ways to capture some of its benefits with easier-to-implement policies?

### 1.1. Contributions

At a high level, the main contributions of this work are in providing a rigorous framework to quantify the potential benefits from targeted confinements and proposing practical implementations that can achieve some of these benefits.

#### The framework

To develop robust and practical insights on the benefits of targeting, we anchor our analysis on a modeling framework that leverages optimization and publicly available, real data. We embed an optimization framework within a multi-group SEIR epidemiological model that differentiates policies based on both population groups and activities, and balances the lost economic value with the cost of deaths. Since the resulting optimization problem is highly nonconvex, we design a novel algorithm — referred to as ROLD (Re-Optimization with Linearized Dynamics) — that can tractably produce high-quality approximate solutions through a linearization and optimization procedure inspired by model predictive control (Bemporad 2006, Camacho and Alba 2013) and trust region methods (Yuan 2015). We then propose a real-life implementation leveraging publicly available data on (i) real-time hospitalization, (ii) community mobility, (iii) social contacts, and (iv) socio-economic measures collected during the COVID-19 pandemic in Île-de-France, a region of France encompassing Paris with a population of approximately 12 million. The framework we develop is flexible, and could be extended to capture other targeted interventions such as testing or vaccinations, as well as additional considerations such as integrality requirements or fairness constraints.

#### Insights

We leverage this framework to address the core research questions posed earlier.

1. **An idealized benchmark for the benefits of targeting**. We first quantify the *potential* benefits that could be achieved in an idealized setting where dual targeting of both activities and age-groups is possible and perfectly enforceable. We find that dual targeting can lead to significant Pareto improvements. Specifically, the optimal ROLD policy that targets both age groups and activities Pareto-dominates the optimal ROLD policies that only target age groups, activities, or neither, i.e., it leads to lower economic costs without increasing the number of pandemic deaths. The gains from targeting are also super-additive: targeting on both dimensions gains more than the sum of gains from unilateral targeting. In addition, the optimized dual-targeted policy also Pareto-dominates a number of benchmarks resembling policies implemented in practice. Finally, although not an explicit objective of the optimization, the dual-targeted ROLD policy also generally reduces the time in confinement for most population groups (and particularly for those groups that are most heavily confined) relative to less fine-grained policies.
2. **Interpretability**. We shed light on the mechanism and the cases when targeting leads to benefits. In term of mechanism, we find that optimized dual-targeted policies impose less confinement on group-activity pairs that generate a relatively high economic value prorated by activity-specific social contacts. This leads to complementary confinement schedules for different groups that reduce both the number of deaths and economic losses, with the important added benefit of not completely confining any group. We confirm these insights by running ROLD on a wide range of problem instances and generating a large dataset, which we use to train decision trees to predict the ROLD decisions based on simple, transparent features. The key feature that explains the targeting is the “econ-to-contacts-ratio,” i.e., the *ratio of marginal economic value to total contacts* generated for a given (group, activity) pair. To understand *when* targeting leads to benefits, we train separate decision trees to predict the gains from dual targeting over less or no targeting. We find that *R*_0_, the cost of death, the heterogeneity in the econ-to-contacts-ratio values, and the severity of the disease are the most important features, and that some of these can have a non-monotonic impact on the magnitude of the gains. In particular, our analysis leads to practical insights about the value of refined targeting depending on the pandemic’s reproduction number and the policy maker’s weighting of health over economic outcomes.
3. **Implementability**. Finally, we propose two new policies that do not explicitly target age and are thus simpler to implement and less contentious, but that nonetheless retain some of the superior health and economic outcomes of idealized dual targeting. Both policies are inspired by real-world pandemic response measures. We first consider curfew policies that restrict activities at targeted times of day, *uniformly* for all age groups. Because different age groups perform distinct activities throughout the day, curfews have the potential to implicitly differentiate based on age. In the second proposal, we consider *recommending*, as opposed to enforcing, targeted activity levels separately for each age group, while accounting for imperfect population compliance. The results show that both proposals can significantly improve over activity-based targeting. Specifically, considering the gap between a dual-targeted and an activity-based targeted policy, we find that a curfew policy can close 24.3% (median value) of that gap and a recommendation-based policy can lead to significant improvements even at relatively modest compliance levels, closing 10.4% (median value) of the gap at 40% compliance and 19.2% at 60% compliance.

## 2. Literature Review

The literature on pandemic response, particularly in the COVID-19 context, is already vast, so we focus our literature review on three key dimensions that our work most closely relates to.

### Targeting

Paralleling our aforementioned real-life examples, several papers have studied targeted interventions. Kucharski et al. (2020), Prem et al. (2020), Di Domenico et al. (2020), Cipriano et al. (2021) recognize the importance of heterogeneity in the social contacts generated through activities and examine several interventions limiting them. Though some of the models here are age differentiated, targeting only happens through activities. Population group targeting, either through confinements, testing or vaccinations, has been investigated in Bastani et al. (2021), Acemoglu et al. (2020), Matrajt et al. (2021), Goldstein et al. (2021), Bertsimas et al. (2020), Favero et al. (2020), Birge et al. (2020), Chang et al. (2020), Evgeniou et al. (2020), Giordano et al. (2021). By enforcing stricter confinements for higher risk groups (e.g., older populations when considering mortality risk or younger populations when considering the risk of new infections), such targeted policies have been shown to generate potentially significant improvements in health outcomes, and even in economic value if optimally tailored (Acemoglu et al. 2020). A potential benefit of our approach is that, by exploiting complementarities between group and activity targeting, these higher risk groups may experience less confinement for the same level of aggregate deaths and economic losses.

### Optimization of interventions in epidemiological models

Our work relates to research that combines epidemiological modeling and optimization techniques to design improved interventions. In general, an epidemic is modeled by a compartmental model, where interventions change the parameters that describe the epidemic with the goal to minimize the health (and economic) burden. Although an analytical characterization of the optimal solution is possible in special cases (Brandeau et al. 2003), the problem is generally intractable and, similar to our framework, research has focused on proposing heuristic algorithms and approximations for solving the general problem (Zaric and Brandeau 2001, 2002). The paper that is most related to ours is Bertsimas et al. (2020), which also proposes a multi-group SEIR formulation, in conjunction with an iterative coordinate descent algorithm to optimize vaccine allocations for COVID-19 in a differentiated fashion. Although taking different approaches to doing so, both the algorithm proposed there and our ROLD heuristic crucially depend on solving linearized versions of the true SEIR dynamics which are tractable via commercial solvers. However, the model of Bertsimas et al. (2020) focuses on vaccine allocation decisions, whereas ours captures the dynamics of differential confinements and also allows activity-based targeting. Bose et al. (2021), Pataro et al. (2021), Morris et al. (2021) also borrow from the optimal control literature, but the models there are simpler than our own and do not capture targeting. Birge et al. (2020) use formal optimization for location-based targeting, but in a one-shot model that does not differentiate age groups or activities and does not account for time in the calculation of health or economic impact. Last, several studies from the operations research community have proposed optimization models to support the allocation of ventilators during epidemics (Huang et al. 2017, Adelman 2020, Mehrotra et al. 2020, Bertsimas et al. 2021). Our paper is also related to a large stream of work that derives prescriptive insights for managing the COVID-19 pandemic. Kaplan (2020) summarizes modeling studies that supported local decisions on event crowd-size restrictions, hospital surge planning, and timing of activity restrictions during COVID-19 response. Several papers simulate a small number of candidate policies for social distancing, e.g., full lockdown versus school-only lockdown (Kucharski et al. 2020, Prem et al. 2020, Di Domenico et al. 2020, El Housni et al. 2020, Favero et al. 2020, Bertsimas et al. 2021), compare a number of current and counterfactual lockdown policies that differ in their schedule of relaxations (Boloori and Saghafian 2023), or restrict the candidates to a simple parametric class for which exhaustive search is computationally feasible (e.g., trigger policies based on hospital admissions as in Duque et al. 2020 or confirmed cases as Ahn et al. 2021). These approaches do not use formal optimization and, when considering a more complex policy space like in our targeting model, could lead to significantly sub-optimal results and misleading conclusions. Navabi-Shirazi et al. (2022) use multicriteria optimization to select the mode (remote, in-person, hybrid) of university courses and assign classrooms, under severely reduced capacities due to COVID-19 social distancing measures. The study of Fotouhi et al. (2021) helps policy makers design curbside restrictions in meal delivery operations that reduce curbside crowding, thus increasing public safety during a pandemic, yet enable delivery companies to retain their profitability.

### COVID-19 forecasting and SEIR model calibration

Several studies have focused on forecasting the health burden of COVID-19 in the presence (or relaxation) of policy interventions. Agent-based simulation has been used to study the effect of government interventions, including vaccination of various efficacy and coverage levels along with non-pharmaceutical interventions (such as reduced mobility, school closings, and use of face masks; Patel et al. 2021, Alagoz et al. 2021); vaccination at different times (Rosenstrom et al. 2022); and social distancing measures implemented at different times and adhered to in different degrees (Alagoz et al. 2020). Linas et al. (2022) employ the COVID-19 policy simulator of Chhatwal et al. (2020) to project COVID-19 mortality as U.S. states relaxed non-pharmaceutical interventions. Li et al. (2022) propose an epidemiological prediction model to quantify the impact of government interventions (mass gathering restrictions, school closings, stay at home) and their timing on the spread of COVID-19. Mandal et al. (2021) show how seroprevalence data could guide a test-and-isolate strategy, for fully lifting COVID-19 restrictions in Indian megacities.

Our work is also related to several other papers that have estimated SEIR parameters, particularly in the COVID-19 context. A number of papers estimate SEIR epidemiological parameters stratified by age groups – we use the estimates from the Île-de-France study in Salje et al. (2020). Another stream of papers, focusing on forecasting COVID-19 spread, estimate when and how the underlying SEIR parameters evolve in response to government interventions and changes in individual behavior, such as Perakis et al. (2021). Lastly, we relate to the literature that has used Google and other mobility data to inform COVID-19 response strategies or to estimate the realized reductions in social contacts during COVID-19 (Dutta et al. 2021, Ilin et al. 2021, Wellenius et al. 2020, Cot et al. 2021, Xiong et al. 2020), as well as the literature on social contacts estimation (Béraud et al. 2015, Prem et al. 2017).

## 3. Model and Optimization Problem

We develop a controlled, multi-group SEIR model that includes time-dependent confinements that can be targeted based on *age groups* and types of *activities* that individuals engage in. The framework is flexible, captures resource constraints such as hospital or ICU capacity, and can be extended to capture other targeted interventions such as testing or vaccinations, as well as additional restrictions that make targeting more fair or practical such as integrality requirements, fairness constraints, etc. We discuss several of the potential extensions in Section 9.

### 3.1. Some Notation

We denote scalars by lower-case letters, as in *v*, and vectors by bold letters, as in ***v***. We use square brackets to denote the concatenation into vectors, ***v*** := [*v*_0_, *v*_1_]. For a time series of vectors ***v***_1_, …, ***v***_*n*_, we use ***v***_*i*:*j*_ := [***v***_*i*_, …, ***v***_*j*_] to denote the concatenation of vectors ***v***_*i*_ through ***v***_*j*_. Lastly, we use ***v***^⊤^ to refer to the transpose of ***v***.

### 3.2. Epidemiological Model and Controls

We rely on a modified version of the discretized SEIR (Susceptible-Exposed-Infectious-Recovered) epidemiological model (Anderson and May 1992, Prem et al. 2020, Salje et al. 2020) with multiple population groups that interact with each other. In our case study we use nine groups *g* ∈ 𝒢 determined by age and split in 10-year buckets, with the youngest group capturing individuals with age 0-9 and the oldest capturing individuals with age 80 or above. Time is discrete, indexed by *t* = 0, 1, …, *T* and measured in days. We assume that no infections are possible beyond time *T*.

#### Compartmental Model and States

Figure 1 represents the compartmental model and the SEIR transitions for a specific group *g*. For a population group *g* in time period *t*, the compartmental model includes states *S*_*g*_(*t*) (susceptible to be infected), *E*_*g*_(*t*) (exposed but not yet infectious), *I*_*g*_(*t*) (infectious). *I*_*g*_(*t*) is further subdivided into *I*_*j,g*_(*t*) for *j* ∈ {*a, ps, ms, ss*} to model different degrees of severity of symptoms: asymptomatic, paucisymptomatic, with mild symptoms, or with severe symptoms; thus, *I*_*g*_(*t*) = Σ_*j*∈{*a,ps,ms,ss*}_ *I*_*j,g*_(*t*). The model also has states *R*_*g*_(*t*) (recovered but not confirmed as having had the virus), 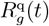(recovered and confirmed as having had the virus), and *D*_*g*_(*t*) (deceased). Individuals with severe symptoms will need hospitalization, either in general hospital wards (*H*_*g*_(*t*)) or in intensive care units (*ICU*_*g*_(*t*)). All the states represent the number of individuals in a compartment of the model in the *beginning* of the time period.

**Figure 1.**
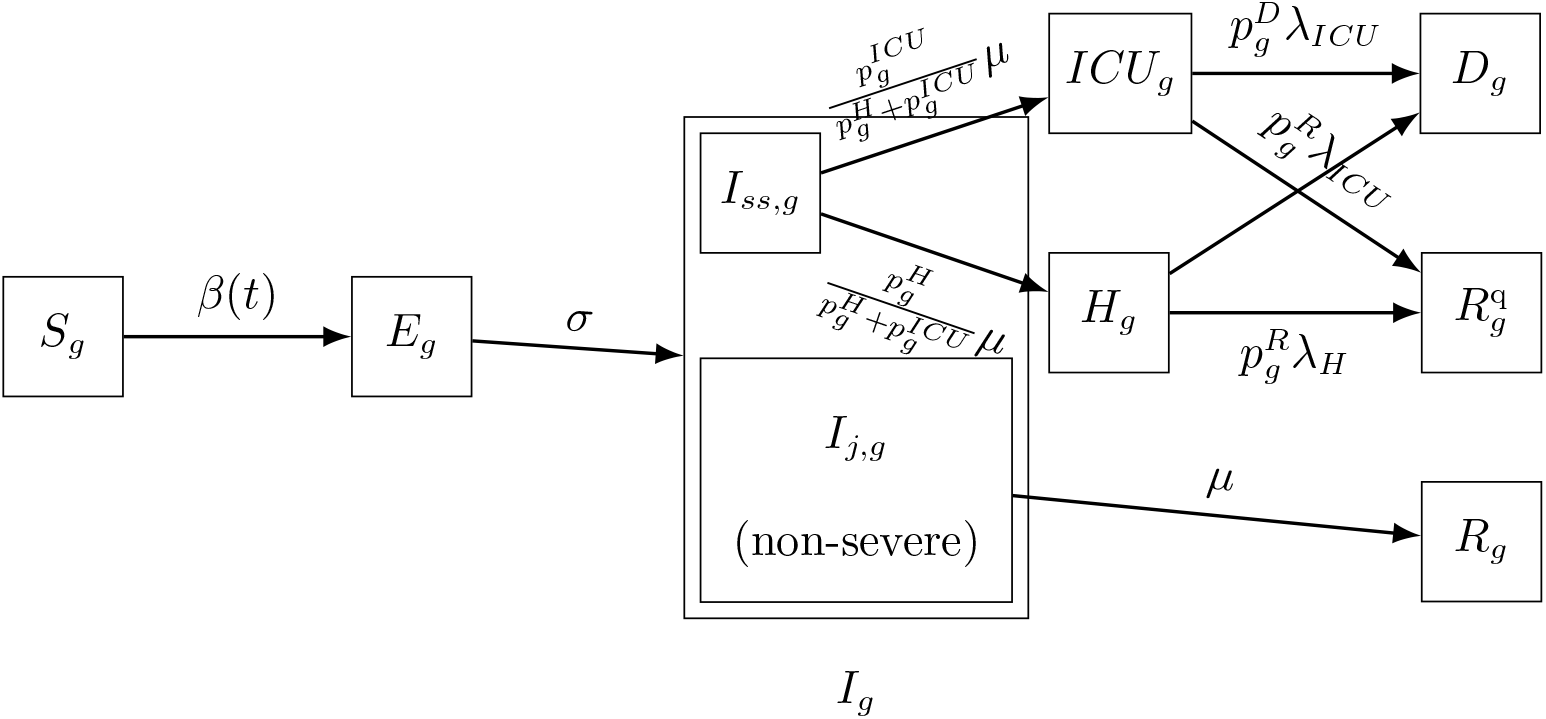
Compartmental SEIR model for a specific group *g* with transition rates.

Susceptible individuals get infected and transition to the exposed state at a rate determined by the number of social contacts and the transmission rate *β*(*t*). Exposed individuals transition to the infectious state at a rate *σ* and infectious individuals transition out of the infectious state at a rate *μ*. We assume that an infectious individual in group *g* exhibits symptoms of degree *j* with probability *p*_*j,g*_. An infectious individual needs to be hospitalized in general hospital wards or ICU with probability 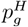 and 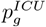, respectively, where 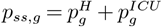. On average, patients treated in general hospital wards (ICU) spend 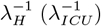 days in the hospital (ICU). An infectious individual with severe symptoms in group *g* deceases (recovers) with probability 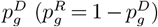.

We keep track of all living individuals in group *g* who are not confirmed to have had the disease *N*_*g*_(*t*) := *S*_*g*_(*t*) + *E*_*g*_(*t*) + *I*_*g*_(*t*) + *R*_*g*_(*t*), and let ***X***_*t*_ = [*S*_*g*_(*t*), *I*_*g*_(*t*), …, *D*_*g*_(*t*)]_*g*∈𝒢_ denote the full state of the system (across groups) at time 0 ≤ *t* ≤ *T*. We denote the number of compartments by |𝒳|, so the dimension of ***X***_*t*_ is |𝒢||𝒳| × 1.

#### Controls

Individuals interact in activities 𝒜 = {work, transport, leisure, school, home, other}. These interactions generate *social contacts* that drive the rate of new infections.

We control the SEIR dynamics by adjusting the confinement intensity in each group-activity pair over time: we let 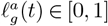 denote the activity level allowed for group *g* and activity *a* at time *t*, expressed as a fraction of the activity level under a *normal course* of life (i.e., no confinement). In our study we take 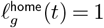, meaning that the number of social contacts at home is unchanged irrespective of confinement policy.1. We denote the vector of all activity levels for group *g* at *t* by 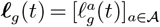, and we also refer to *ℓ*_*g*_(*t*) as confinement decisions when no confusion can arise.

We propose a parametric model to map activity levels to social contacts. We use *c*_*g,h*_(*ℓ*_*g*_, *ℓ*_*h*_) to denote the mean number of total daily contacts between an individual in group *g* and individuals in group *h* across all activities when their activity levels are *ℓ*_*g*_, *ℓ*_*h*_, respectively. Varying the activity levels changes the social contacts according to

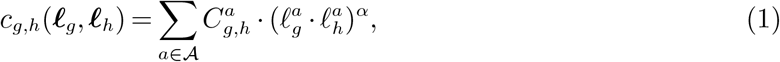

where 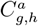 denote the mean number of daily contacts in activity *a* under normal course (i.e., without confinement) and *α* ∈ ℝ is a social mixing parameter that captures the elasticity of social contacts to activity levels. This parametrization is similar to a Cobb-Douglas production function (Mas-Colell et al. 1995), using the activity levels as inputs and the number of social contacts as output. We retrieve values for 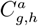 from the data tool of Wille et al. (2020), which is based on the French social contact survey data in Béraud et al. (2015), and we estimate *α* from health outcome data (French Government 2020) and Google mobility data (Google 2020), as described in Section EC.4.2.

Let ***u***_*t*_ = [*ℓ*_*g*_(*t*)]_*g*∈𝒢_ denote the vector of all decisions at time *t* ∈ {0, 1, …, *T* − 1}, i.e., the confinement decisions for all the groups. We denote the number of different decisions for a given group at a given time by |𝒰|. Then the dimension of ***u***_*t*_ is |𝒢||𝒰| × 1.

### 3.3. Resources and Constraints

We use *K*^H^(*t*) (*K*^ICU^(*t*)) to denote the capacity of beds in general hospital wards (ICU) on day *t*. When the patient inflow into the hospital or the ICU exceeds the remaining number of available beds, then the policy maker needs to decide how many patients to turn away from each group. Although our framework allows optimizing over such decisions, we choose to not consider this dimension of targeting because it can be extremely contentious in practice. Instead, we implement a proportional rule that allocates any remaining hospital and ICU capacity among patients from all age groups proportionally to the number of cases requiring admission from each group. More formally, with 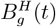 denoting the number of patients from age group *g* who are denied admission to general hospital wards in period *t*, the proportional rule is:

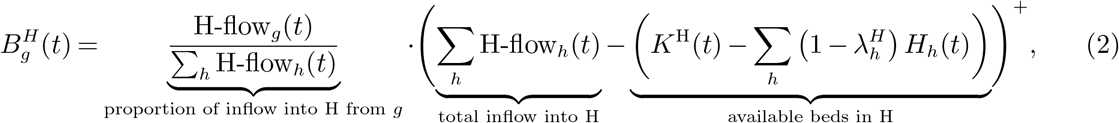

where 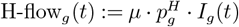, and a similar rule holds for patients requiring admission to the ICU. We assume that all patients that are denied admission die immediately.

We can now write a complete set of discrete dynamical equations for the controlled SEIR model ((EC.1)-(EC.9) in Appendix EC.1) and summarize these using the function

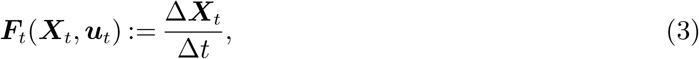

where Δ***X***_*t*_ := ***X***_*t*+1_ − ***X***_*t*_. Additionally, we also include the following constraints:

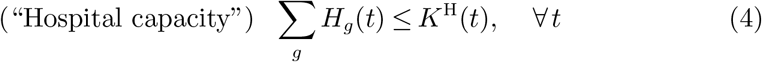

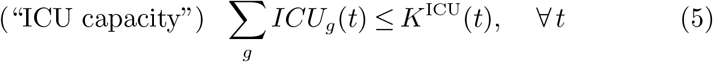

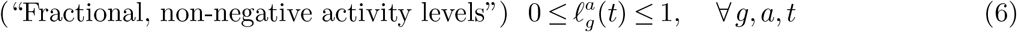

We denote by 𝒞(***X***_*t*_) the feasible set described by (4)-(6)for the vector of decisions ***u***_*t*_ at time *t*.

### 3.4. Objective

Our objective captures two criteria. The first quantifies the total deaths directly attributable to the pandemic, which we denote by Total Deaths(***u***_0:*T* −1_) :=Σ_*g*∈𝒢_ *D*_*g*_(*T*) to reflect the dependency on the specific policy ***u***_0:*T* −1_ followed. The second criterion captures the economic losses due to the pandemic, denoted by Economic Loss(***u***_0:*T* −1_). These stem from three sources: (a) lost productivity due to confinement, (b) lost productivity during the pandemic due to individuals being quarantined, hospitalized, or deceased, and (c) lost value after the pandemic due to deaths (as deceased individuals no longer produce economic output even after the pandemic ends).

To model (a), we assign a daily economic value *v*_*g*_(*ℓ*) to each individual in group *g* that depends on the activity levels *ℓ*:= [*ℓ*_*g*_]_*g*∈𝒢_ across all groups and activities. For the working age groups, *v*_*g*_(*ℓ*) comes from wages from employment and is a linear function of group *g*’s activity level in work 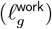 and of the average activity levels in leisure, other and transport for the entire population (equally weighted). This reflects that the value generated in some industries, like retail, is impacted by confinements across all these three activities. For the school age groups, *v*_*g*_(*ℓ*) captures future wages from employment due to schooling and depends only on the group’s activity level in school 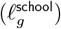. For (b), we assume that an individual who is in quarantine, hospitalized, or deceased, generates no economic value. At the same time, we assume that individuals in 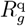 generate economic value as they would under no confinement. For (c), we determine the wages that a deceased individual would have earned based on their current age until retirement age under the prevailing wage curve, and denote the resulting amount of lost wages with 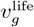.

The overall economic loss is the difference between the economic value that would have been generated during the pandemic under a “no pandemic” scenario (*V*) and the value generated during the pandemic, plus the future economic output lost due to deaths.

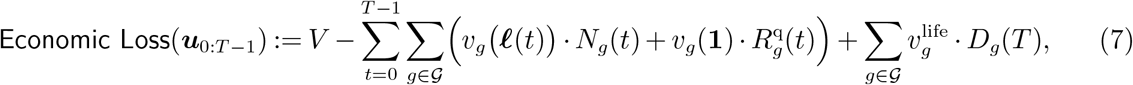

All the details of the economic modelling are deferred to Appendix EC.2.

To allow policy makers to weigh the importance of the two criteria, we associate a cost *χ* to each death, which we express in multiples of GDP per capita. Our framework can capture a multitude of policy preferences by considering a wide range of *χ* values, from completely prioritizing economic losses (*χ* = 0) to completely prioritizing deaths (*χ* → ∞).

### 3.5. Optimization Problem

The optimization problem we solve is to find control policies for confinement that minimize the sum of mortality and economic losses2. subject to the constraints that (i) the state trajectory follows the SEIR dynamics, and (ii) the controls and states respect the capacity and feasibility constraints discussed above. Formally, we solve:

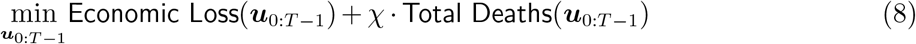

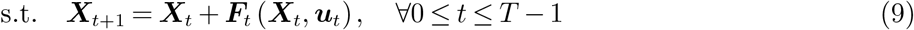

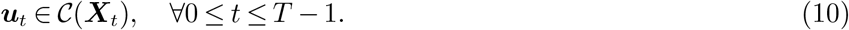

## 4. Algorithm: Re-Optimization with Linearized Dynamics

Solving problem (8)-(10) to optimality unfortunately requires solving intractable optimization problems. To see this, note that the key term in the dynamics of any SEIR-type model is the rate of new infections, which involves multiplying the current susceptible population with the infected population. This introduces non-linearity in the state trajectory; for instance, our dynamic for the evolution of the susceptible population in group *g* from (EC.2) reads:

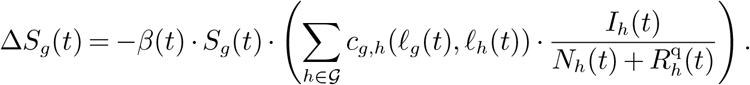

Expanding out *S*_*g*_(*t*) produces a complex, non-convex dependency on the past decisions *ℓ*(*τ*) for 0 ≤ *τ* ≤ *t* − 1, which makes the resulting problem intractable via convex optimization.

With this in mind, we focus on developing heuristics that can tractably yield good policies, and we propose an algorithm called Re-Optimization with Linearized Dynamics, or ROLD, that builds a control policy by incrementally solving linear approximations of the true SEIR system.

### 4.1. Linearization and Optimization

The key idea is to solve the problem in a shrinking-horizon fashion, where at each time step *k* = 0, …, *T* we linearize the system dynamics and objective (over the remaining horizon), determine optimal decisions for all *k*, …, *T*, and only implement the decisions for the current time step *k*.

We first describe the linearization procedure. Recall that the true evolution of our dynamical system is given by (3). The typical approach in dynamical systems is to linearize the system dynamics around a particular “nominal” trajectory. More precisely, assume that at time *k* we have access to a nominal control sequence 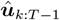 and let 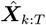 denote the resulting nominal system trajectory under the true dynamic (3) and under 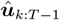. We approximate the original dynamics through a Taylor expansion around 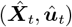:

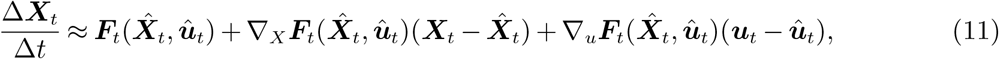

where ∇_*X*_***F***_*t*_ and ∇_*u*_***F***_*t*_ denote the Jacobians with respect to ***X***_*t*_ and ***u***_*t*_, respectively. Note that these Jacobians are evaluated at points on the nominal trajectory, so (11) is indeed a linear expression of ***X***_*t*_ and ***u***_*t*_. By induction, every state ***X***_*t*_ under dynamic (11) will be a linear function of ***u***_*τ*_ for *τ < t*, and all the constraints will also depend linearly on the decisions.

In a similar fashion, we also linearize the objective (8). Since *v*_*g*_(*ℓ*(*t*)) is linear in ***u***_*t*_ for all *t* = 0, …, *T* − 1, the objective contains *bilinear* terms and can be written compactly as:

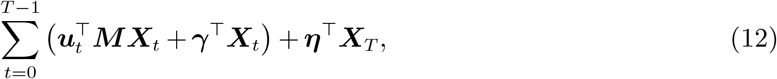

for some matrix ***M*** with dimensions |𝒢|| 𝒰| × |𝒢|| 𝒳|, and vectors ***γ*** and ***η*** of dimensions |𝒢|| 𝒳| × 1 (detailed expressions are available in Appendix EC.3). By linearizing this using a Taylor approximation, we consider the following objective instead:

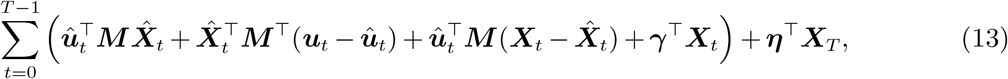

which depends linearly on all the decisions ***u***_0_, …, ***u***_*T* −1_.

#### Linearization-optimization procedure

We use the following heuristic to obtain an approximate control at time *k*, for *k* = 0, …, *T* − 1:

1. Given the current state ***X***_*k*_ and a nominal control sequence 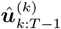 for all remaining periods, calculate a nominal system trajectory 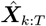 under the true dynamic in (3). (The nominal control sequence is set to a solution obtained by a gradient descent method at *k* = 0, and to the algorithm’s own output from period *k* − 1 for periods *k >* 0, per Step 4 below.)
2. Use (11) to approximate the state dynamic around the nominal trajectory 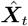 and use (13) to approximate the objective-to-go function over the remaining periods *t* ∈ {*k*, …, *T* }.
3. Solve the linear program to obtain decision variables 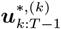 that maximize the linearized objective-to-go subject to all the relevant linearized constraints.
4. Set the nominal control sequence for the next time step as 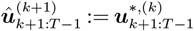.
5. Update the states using the optimal control 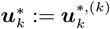 and the true dynamic in (3), i.e. 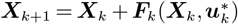.

The linearization-optimization procedure described above is run for all periods *k* = 0, …, *T* − 1 sequentially to output a full control policy 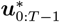.

#### Trust region implementation

In our experiments, we have found that the linearized model described in (11) may diverge significantly from the real dynamical system when the optimized controls 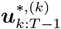 determined in Step 3 diverge sufficiently from the nominal controls 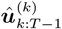 considered in the linearization in Step 2. This can lead to a large sensitivity in performance to the initialization used in the very first step; for example, if the Taylor approximation were constructed around a policy of full confinement, the linearized model could systematically underestimate the number of infections and deaths created when considering more relaxed confinements.

We overcome this by employing an iterative procedure inspired by a trust region optimization method. The key idea is to avoid the large approximation errors by running the linearization-optimization procedure iteratively within each time step *k*, with each iteration only being allowed to take a small step towards the optimum within a trust region of an *E*-ball around the nominal control sequence 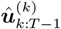, and the updated optimized control sequence of each iteration being used as a nominal sequence for the next iteration. This leads to a procedure that is much more robust to the initial guess of control sequence, albeit at the expense of increased computation time.

Further algorithmic details for ROLD are provided in Appendix EC.3.

## 5. Île-de-France Calibration and Experimental Setup

To complete the description of our framework, we now summarize our approach for calibrating our model using real-world publicly available data on (i) community mobility, (ii) social contacts, (iii) health outcomes, and (iv) economic output for the Île-de-France region of France.

### 5.1. Parametrization and Calibration

We adopt values for disease progression parameters for Île-de-France directly from the study by Salje et al. (2020); we adopt values for the number of social contacts across different age groups and activities from the study of Béraud et al. (2015) focused on France, using the tool by Wille et al. (2020); and we use Google mobility data (Google 2020) for the Île-de-France region to approximate the mean effective lockdowns for all activities during the horizon of interest. We then estimate any remaining parameters by minimizing the sample-average approximation of an error metric derived by comparing our model’s predictions (on several sample paths) with time-series data from the French Public Health Agency (French Government 2020) on hospital and ICU utilization and deaths. We calibrate our economic model using data on full time equivalent wages and employment rates from the French National Institute of Statistics and Economic Studies, and sentiment surveys on business activity levels during confinement from the Bank of France. We provide all the details for calibration and parameter specification in Appendix EC.4. We report experimental results from sensitivity and robustness analyses on the fitted parameters in Appendix EC.6.

### 5.2. Model Validation

Table EC.5 in the Appendix summarizes the parameter values obtained from our calibration procedure, and Figure 2 compares the fitted model’s predictions with the reported values for hospital beds utilization, ICU beds utilization, and cumulative deaths. To further validate our model, we also assess the goodness of fit *out of sample*. In particular, for each date *t* from a set of four dates in 2020, we calibrate the SEIR model using data up until day *t* − 14, we simulate the calibrated model up until day *t*, and we compare the model predictions with the reported data in the interval [*t* − 13, *t*]. Figure EC.1 shows that the predictions of the calibrated SEIR model for all three metrics of interest stay close to the reported values in the out-of-sample validation set. As expected, the fit worsens when the model is estimated with less data (e.g., it is worst for the earliest of the four dates shown).

**Figure 2.**
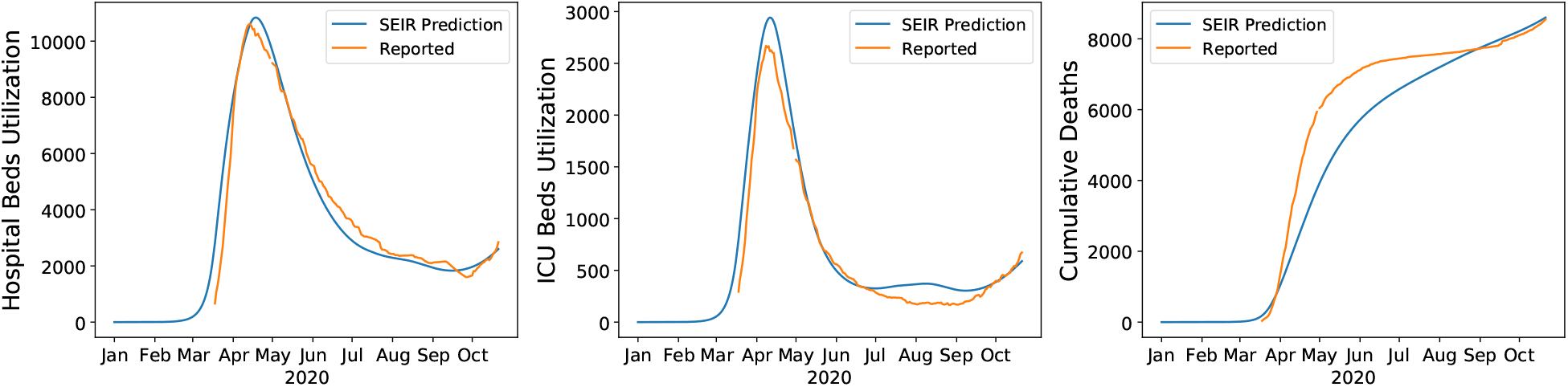
Predictions of the fitted SEIR model vs. reported values by the French Public Health Agency for Hospital Beds Utilization, ICU Beds Utilization and Cumulative Deaths.

### 5.3. Experimental and Optimization Setup

We run experiments over a range of values for our model parameters, summarized in Table 1. For parameters for which multiple values are used in our experiments, the “Baseline Value” column reports the values used in results in the main paper and the Appendix, unless specified otherwise. In particular, we use a baseline capacity of 2900 ICU beds in Île-de-France, and experiment with ICU capacities that range from 2000 to 3200 beds.3. We use an infinite capacity for general hospital wards. We optimize decisions starting on October 21 2020. We allow confinement decisions to change every two weeks.

**Table 1.** Parameter values for experimental and optimization setup. The parameters *ν*^other activities^, *r, f*_*g*_ and *θ* related to our economic model are defined in Appendix EC.2

| Parameter Description | Notation | Values in Experiments | Baseline Value |
| --- | --- | --- | --- |
| Cost of death | $\chi$ | 30 values in $[0, 1000] \times \text{GDP per capita of France}$ | |
| GDP per capita of France |  | €37199.03 |  |
| ICU capacity | $K^{\text{ICU}}$ | 2000, 2300, 2600, 2900, 3200 | 2900 |
| Hospital capacity | $K^{\text{H}}$ | ∞ | |
| Sensitivity of econ. value on confinement | $\nu^{\text{other activities}}$ | 0, 0.1, 0.2 | 0.1 |
| Discount rate (used to calculate $v_g(\ell)$ and $v_g^{\text{life}}$) | $r$ | 0.03 | |
| Fraction going to school | $f_g$ | 1 for $g = 0-9$ y.o.<br>0.907 for $g = 10-19$ y.o. | |
| Mult. factor for value of schooling | $\theta$ | 0.5, 1, 5 | 0.5 |
| Starting time for optimization |  | October 21 2020 |  |
| Optimization horizon | $T'$ | {90, 180, 360} days | 90 days |
| Frequency of confinement decisions |  | 14 days |  |

#### Optimization Horizon

We use an optimization horizon of *T*′ = 90 days in the experiments reported in the main paper (and allow up to 360 days in additional experiments). To optimize confinements using ROLD and other policies on days 0, 1, …, *T*′− 1, we set the total time horizon to be *T* = *T*′ + 14 and further constrain the policies to be fully open on days *T*′, …, *T*′ + 13, while assuming no infections are possible starting on day *T*′. We do this to mitigate possible end-of-horizon effects: any deaths and loss of economic value between day *T*′ + 1 and *T*′ + 14 will still count towards the objective, so ROLD cannot quite allow for too many infections towards the end of the *T*′-day optimization horizon. We discuss further techniques for mitigating end-of-horizon effects in Section 9.

#### ROLD Variants

To quantify the benefits of targeting, we consider several ROLD policies that differ in the level of targeting allowed, which we compare over a wide range of values for *χ*, from 0 to 1000× the annual GDP per capita in France.4. For each *χ* value, we calculate all the ROLD policies of interest, and we record separately the economic losses and the number of deaths generated by each policy. The four versions of ROLD we consider are no targeting whatsoever (NO-TARGET), targeting age groups only (AGE), targeting activities only (ACT), or targeting both (AGE-ACT, or simply ROLD when no confusion can arise). To obtain each policy, we run suitably constrained versions of the ROLD optimization problem initialized using the solution of a gradient descent algorithm (details in Appendix EC.3.5).

## 6. How Large Are the Potential Gains from Dual Targeting?

We next apply our framework to the Île-de-France context to quantify the magnitude of the gains from targeting in an idealized setting where dual targeting can be implemented. This will then serve as a useful benchmark against which to assess more implementable targeted policies.

To isolate the benefits of each type of targeting, we compare the four versions of ROLD that differ in the level of targeting allowed, as described in Section 5.3. Figure 3a records each policy’s performance in several problem instances, where each problem instance corresponds to a different value for the cost of death *χ*. A striking feature is that each of the targeted policies actually *Pareto-dominates* the NO-TARGET policy, and the improvements are significant: relative to NO-TARGET and for same number of deaths, economic losses are reduced by EUR 0-2.9B (0%-36.4%) in AGE, by EUR 0.6B-2.2B (6.5%-52.3%) in ACT, and by EUR 2.3B-5.4B (24.4%-80.6%) in AGE-ACT. This Pareto-dominance is unexpected since it is not explicitly required in our optimization procedure, and it underlines that any form of targeting can lead to significant improvements in terms of *both* health and economic outcomes.

**Figure 3.**
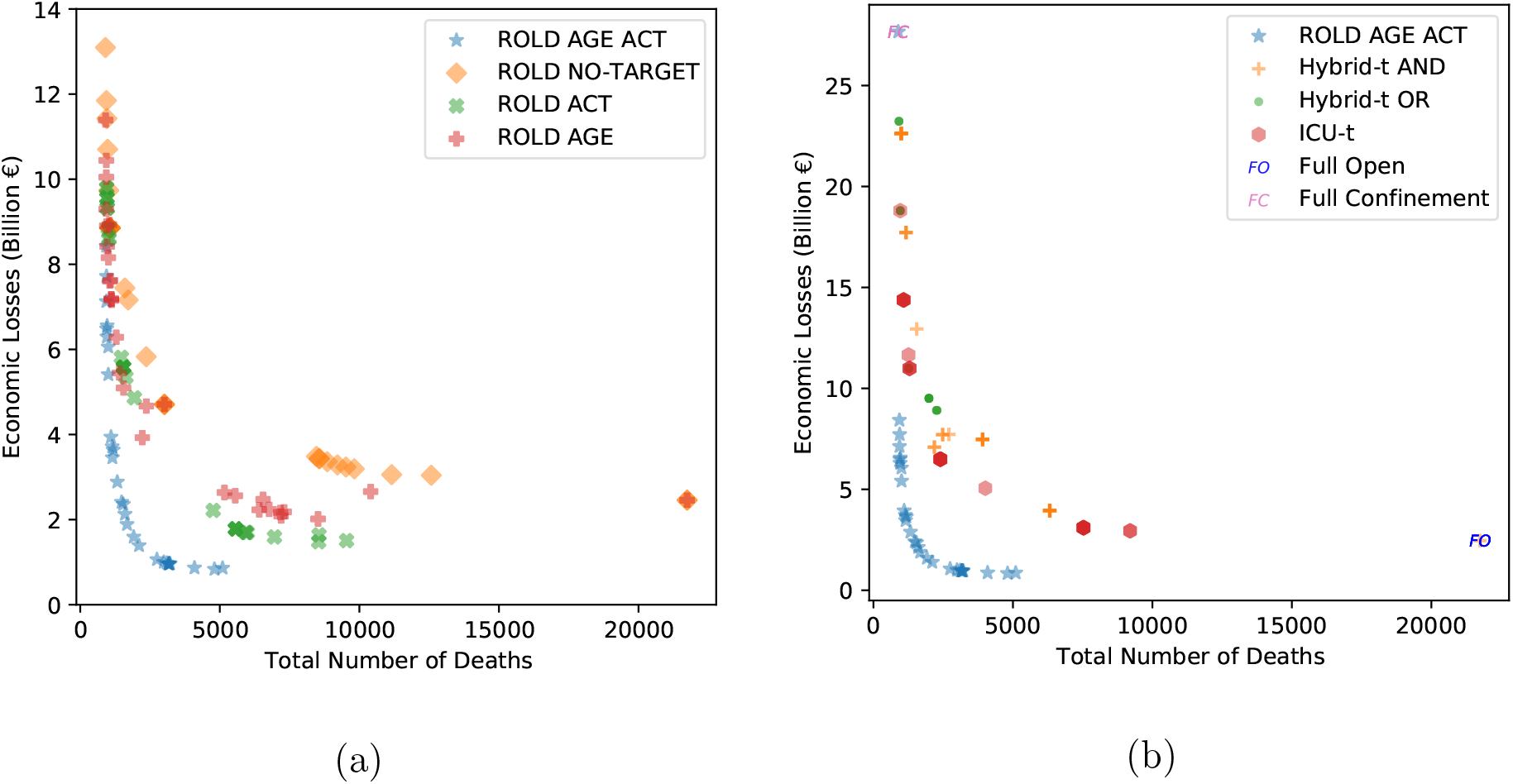
The total number of deaths and the economic losses generated by targeted ROLD policies and the benchmark policies. Panel **(a)** compares the four versions of ROLD that differ in the level of targeting allowed. Panel **(b)** compares the ROLD policy that targets age groups and activities with the benchmark policies. Each marker corresponds to a different problem instance parameterized by the cost of death *χ*. We include 30 distinct values of *χ* from 0 to 1000×, and panel (b) also includes a very large value (*χ* = 10^16^×, for which ROLD AGE-ACT recovers the full confinement policy).

When comparing the different types of targeting, neither AGE nor ACT Pareto-dominate each other, and neither policy dominates in terms of the total loss objective (Figure 4a). In contrast and crucially, AGE-ACT Pareto-dominates *all* other policies (and also dominates in terms of the total loss objective). Moreover, targeting both age groups and activities leads to super-additive improvements in almost all cases: for the same number of deaths, AGE-ACT reduces economic losses by more than AGE and ACT *added together* (Figure 5). This suggests that substantial complementarities may be unlocked through *dual* targeting that may not be available under less granular targeting. These results are robust under more problem instances (Appendix Section EC.6.1).

**Figure 4.**
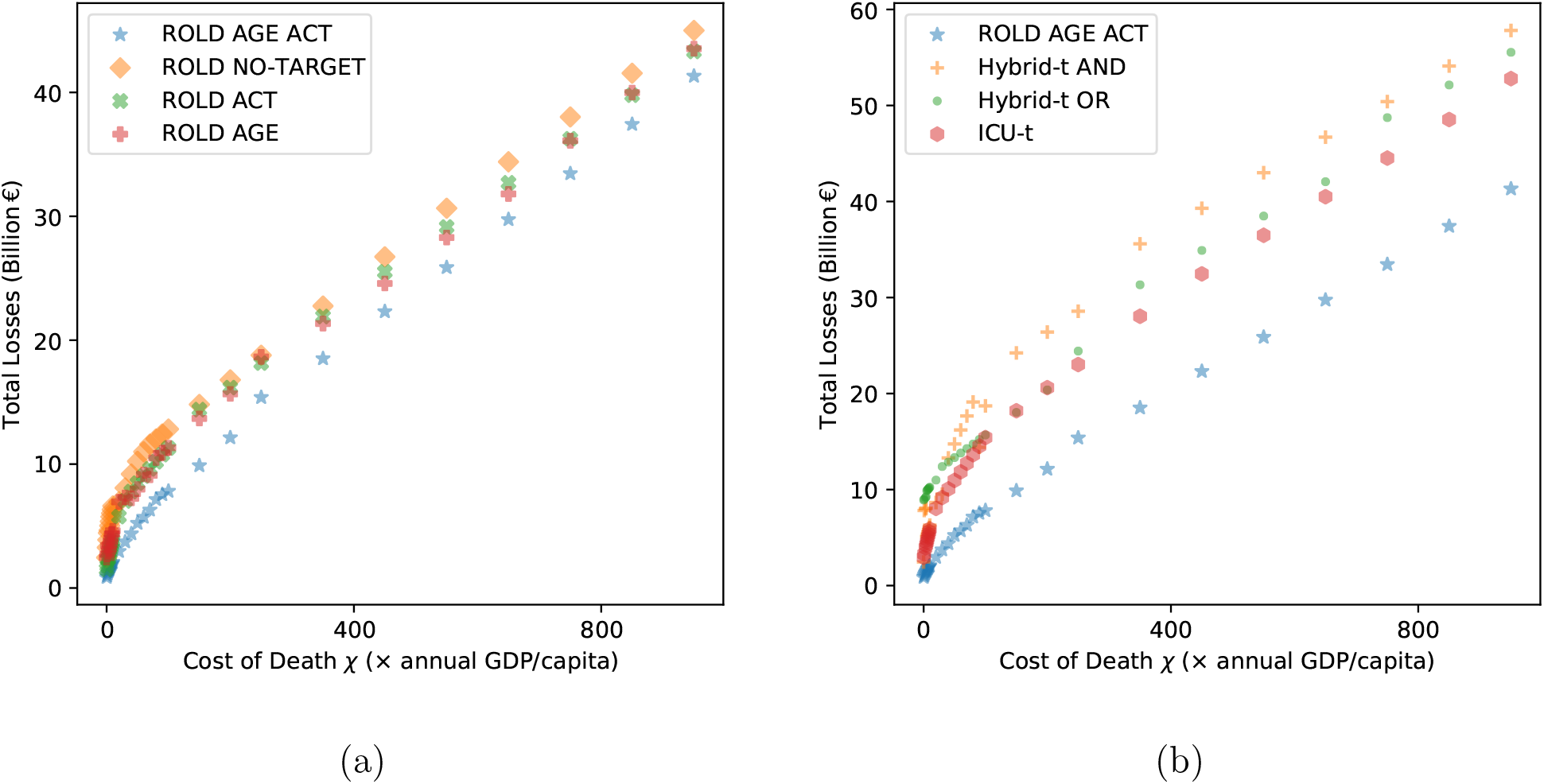
Total losses generated by targeted ROLD policies and by the benchmark policies, at different values of the cost of death *χ*. Panel **(a)** compares the four versions of ROLD that differ in the level of targeting allowed. Panel **(b)** compares the ROLD policy that targets age groups and activities with the benchmark policies. Each marker corresponds to a different problem instance parameterized by the cost of death *χ*. We include 30 distinct values of *χ* from 0 to 1000×.

**Figure 5.**
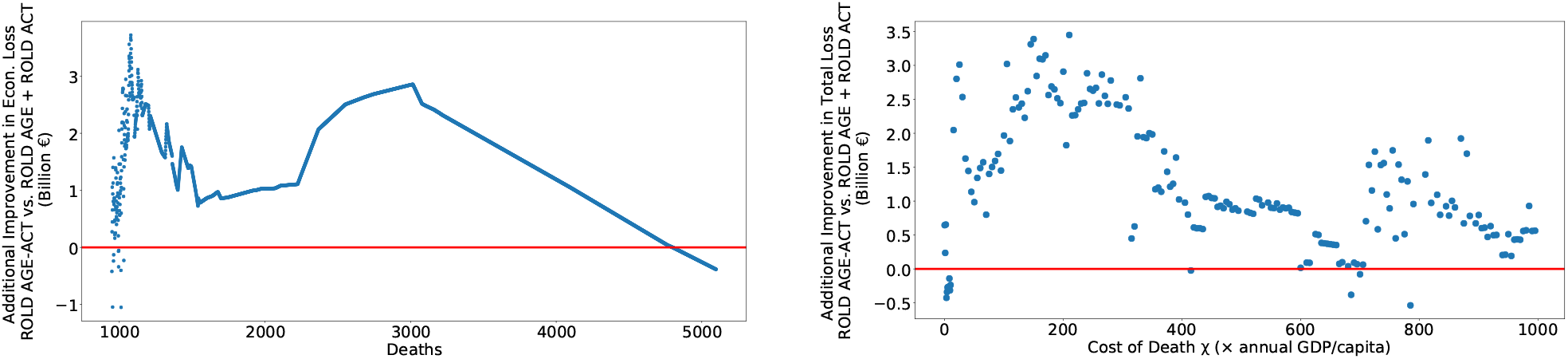
The super-additivity of ROLD AGE-ACT. The figures compare the improvement from AGE-ACT with the sum of the improvements from AGE and ACT. All improvements are with respect to NO-TARGET. The left panel compares the improvements in economic losses for the same number of deaths. The right panel compares the improvements in total losses for the same cost of death *χ*.

To gain additional perspective on these idealized gains, we also compare ROLD AGE-ACT with various practical benchmark policies in Figures 3b, 4b. Benchmarks ICU-t and Hybrid-t AND/Hybrid-t OR mimic implementations in the U.S. Austin area (Duque et al. 2020) and, respectively, France (Lehot and Borgne 2020). These policies switch between a stricter and a relaxed confinement level based on conditions related to hospital admissions and occupancy and the rate of new infections (Appendix EC.5). We optimize the parameters of these benchmark policies and compare with the best-performing model in each class. We also consider two extreme benchmarks corresponding to enforcing “full confinement” (FC) or remaining “fully open” (FO), which can be expected to perform well when completely prioritizing the number of deaths (i.e., *χ* → ∞) or the economic losses (i.e., *χ* = 0), respectively.

ROLD Pareto-dominates all these benchmarks, decreasing economic losses by EUR 5.8B-16.9B (66.6%-84.0%) relative to Hybrid-t AND, by EUR 7.3B-11.8B (55.5%-85.7%) relative to Hybrid-t OR, and by EUR 5.7B-11.8B (55.5%-81.9%) relative to ICU-t for the same number of deaths. Additionally, ROLD meets or exceeds the performance of the two extreme policies: for a sufficiently large *χ*, ROLD exactly recovers the FC policy, resulting in 890 deaths and economic losses of EUR 27.7B; for a sufficiently low *χ*, ROLD actually Pareto-dominates the FO policy, reducing the number of deaths by 16,642 (76.6%) and reducing economic losses by EUR 1.6B (64.9%). The latter result, which may seem surprising, is driven by the natural premise captured in our model that deaths and illness generate economic loss because of lost productivity; thus, a smart sequence of confinement decisions can actually improve the economic loss relative to FO. Among all the policies we tested, ROLD AGE-ACT was *the only one* capable of Pareto-dominating the FO benchmark, confirming that dual targeting is critical and powerful.

Another possible benefit of finer targeting is that it could reduce the time in confinement for all groups. To check this, we calculate the time-average activity level for each age group under each ROLD policy, averaged over the activities relevant to that age group and weighting each activity by the participation rate of that age group in that activity (details provided in Appendix EC.6.2). The results are visualized in Figure 6. The dual-targeted AGE-ACT policy has higher population-wide activity levels relative to all other policies and maintains higher activity levels for most age groups, with systematically higher levels for the age groups that are *most confined*. In particular, for the AGE-ACT policy the group with the lowest median activity level (of 81.7%) is the 40-49 y.o., whereas under the AGE (ACT) policy, that group is the 80+ (60-69) y.o., with median activity level of 34.2% (75.3%). This is a reasonable requirement when considering the “fairness” of confinement policies.5. Moreover, even when AGE-ACT confines certain age groups more than other policies, it never does so by much: its activity levels are within 5% (in absolute terms) from the levels achieved by ACT (respectively, AGE and NO-TARGET) for *every* age group in 96% (57%, 98%) of all instances, and within 10% from ACT (respectively, AGE and NO-TARGET) for every age group in 98% (88%, 98%) of all instances.

**Figure 6.**
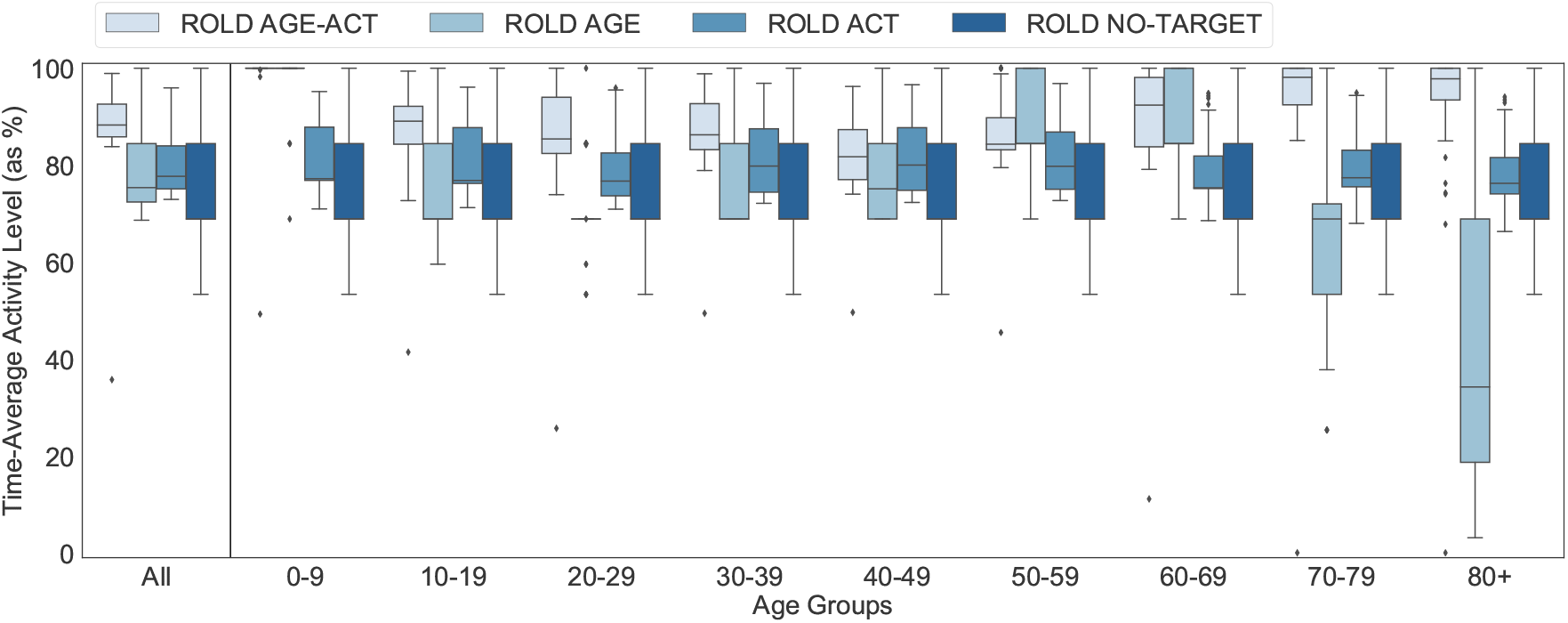
Time-average activity level for the targeted ROLD policies. Each boxplot depicts the time-average activity level under the respective policy averaged over the activities relevant to that age group and weighting activities by participation rates, for different problem instances parameterized by *χ*. The first boxplot on the left depicts activity levels aggregated over all the age groups.

These results suggest that dual targeting has the potential to improve outcomes along multiple dimensions: in terms of *both* health and economic outcomes, and by generating more socially acceptable confinements, through a reduced and more “fair” distribution of confinement time.

## 7. How Do Gains Arise from Dual Targeting?

That targeting can generate such improvements along multiple metrics is quite unexpected because this is not something that the ROLD framework explicitly optimizes for (recall that the ROLD objective only involves a total loss metric). To better understand how gains could arise from targeting, we now examine the structure of the optimal dual-targeted ROLD decisions and explain these as well as their associated gains in terms of a set of interpretable features.

### 7.1. Dual Targeting Generates Complementarities

By examining the optimal ROLD AGE-ACT decisions, we first note that the policy maintains high activity levels for (age group, activity) pairs with high economic value and few social contacts. This can be visualized in Figure 7, which plots the activity levels for different (age group, activity) pairs divided into three buckets (high, medium, low) depending on their econ-to-contacts-ratio. This is defined as the marginal economic value divided by the total social contacts generated by a group in an activity, i.e.,

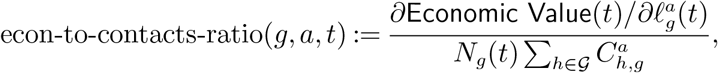

where 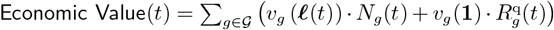. Similarly, Figure 8 shows that ROLD prioritizes activity in transport, school, work, other, and then leisure, in that order, in accordance with the relative econ-to-contacts-ratio of these activities.6.

**Figure 7.**
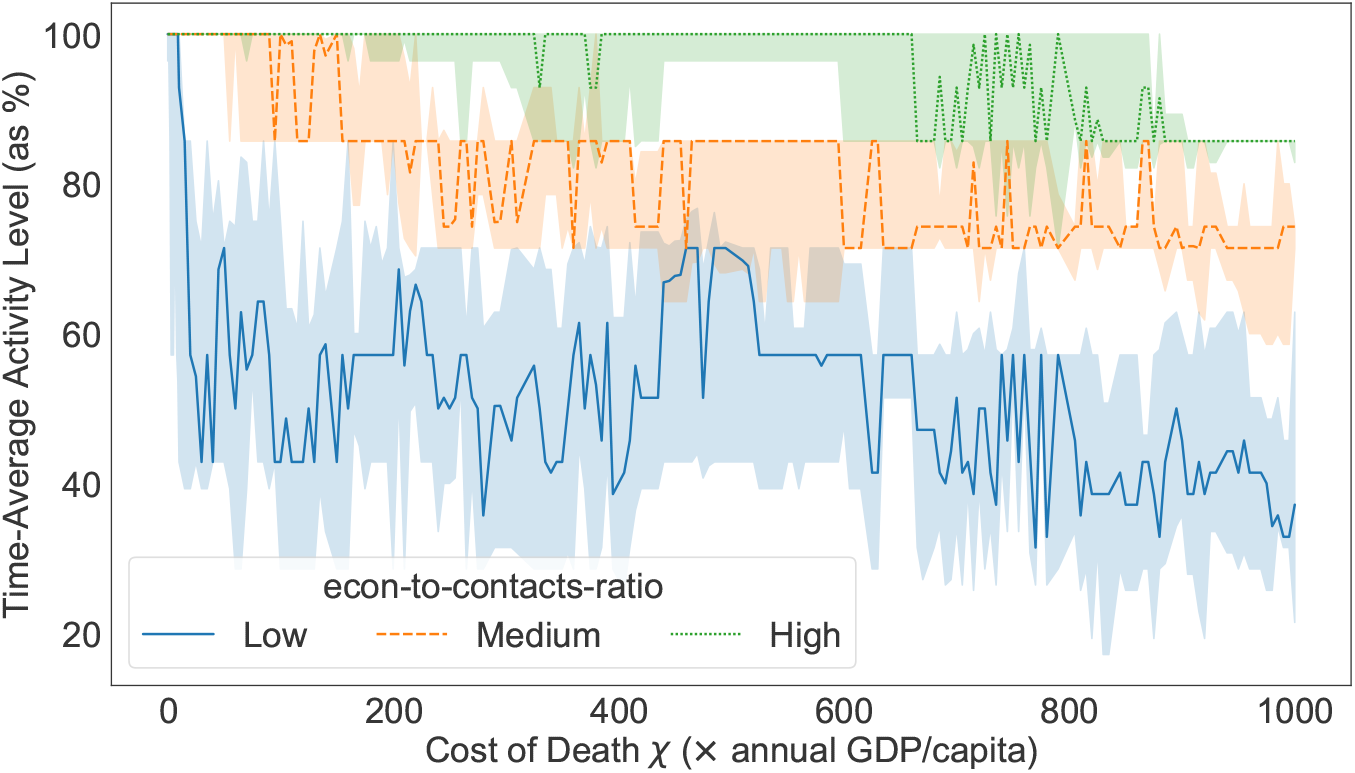
The time-average activity levels of optimized ROLD AGE-ACT policies for different (age group, activity) pairs, bucketed in three equally-sized buckets according to their econ-to-contacts-ratio. The lines indicate the median for each bucket while the bands indicate interquartile ranges, where for each value of the cost of death *χ*, the statistics are taken over the relevant (age group, activity) pairs.

**Figure 8.**
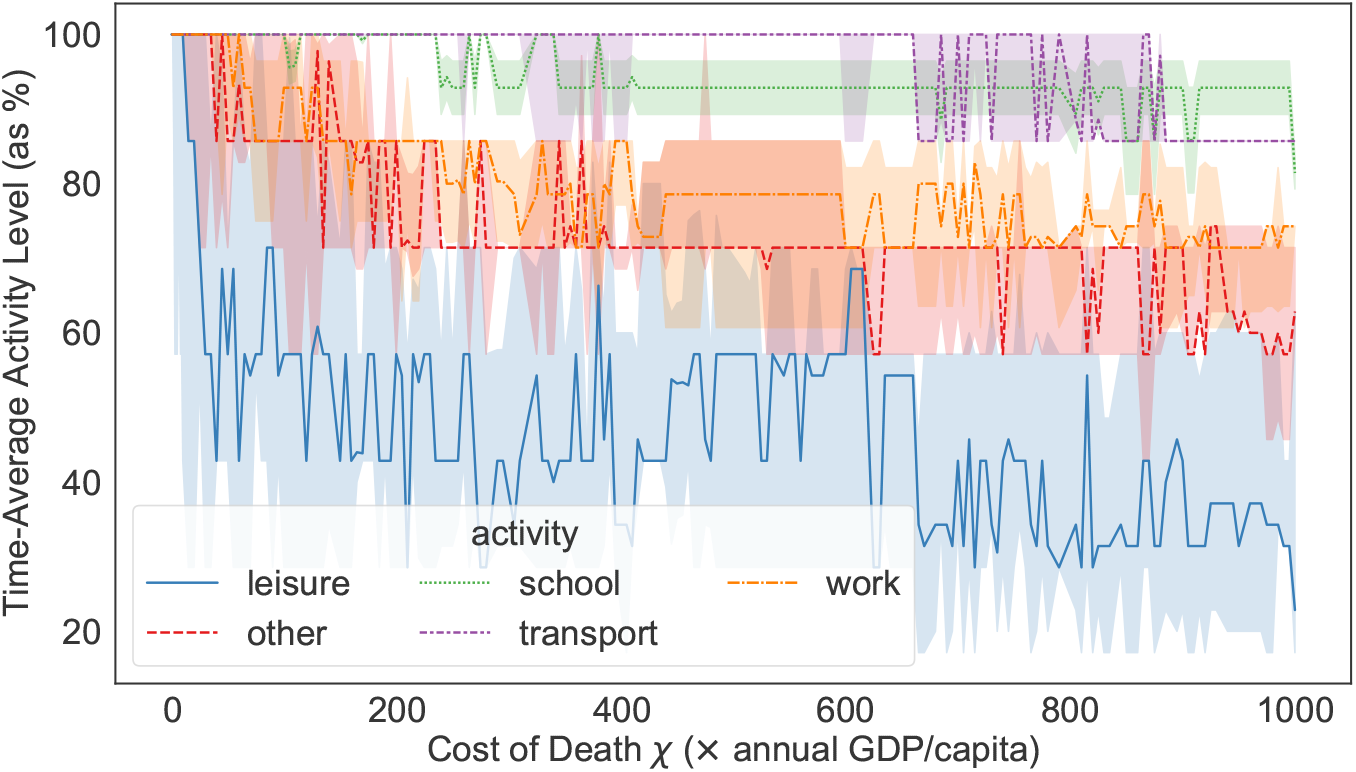
The time-average activity levels of optimized ROLD AGE-ACT policies for different activities. The lines indicate the median for each activity, while the bands indicate interquartile ranges, where for each value of the cost of death *χ* the statistics are taken over the relevant age groups for that activity.

To understand how *complementarities* arise in this context, note that the ability to separately target age groups and activities allows the ROLD policy to fully exploit the fact that distinct age groups may be responsible for the largest econ-to-contacts-ratio in different activities. As an example, the 20-69 y.o. groups have the highest ratio in work, whereas the 0-19 y.o. and 70+ y.o. groups have the highest ratio in leisure. Accordingly, ROLD coordinates confinements to account for this (see Figure 9): groups 20-69 y.o. remain more open in work but face confinement in leisure for up to the first ten weeks, whereas groups 0-19 and 70+ y.o. remain quite open in leisure but have reduced activity in work. These complementary confinement schedules allow ROLD to reduce both the number of deaths and economic losses, with the important added benefit that no age group is completely confined. (For an additional visualization, Figure EC.4 in the Appendix depicts the ROLD activity levels for a specific value of the cost of death *χ*.)

**Figure 9.**
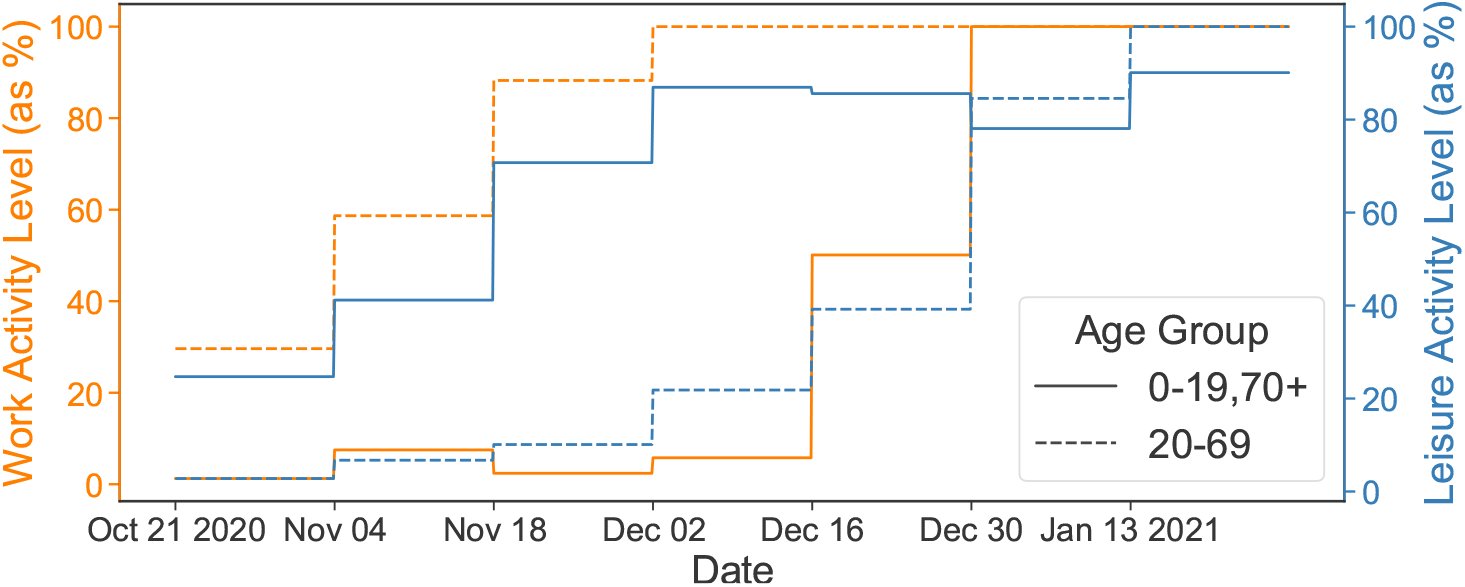
The activity levels of optimized ROLD AGE-ACT policies for work and leisure, and for the 20-69 y.o., vs. the 0-19 and 70+ y.o. age groups. The lines indicate mean activity levels, where for each time *t* the mean is taken across different values for the cost of death *χ* and across the respective relevant age groups. (The figure is best viewed in color.)

### 7.2. Understanding Dual-Targeted Policies and Their Gains

To gain a better understanding of the ROLD policy and the potential benefits of dual targeting, we also take a different approach and train interpretable machine learning models (regression trees) to predict (i) the optimal ROLD decisions and (ii) the gains from dual targeting, as a function of several salient epidemiological and economic features.

More specifically, we generate problem instances with randomly drawn values from a wide range for the transmission rate *β* (equivalently, for the basic reproduction number *R*_0_), the disease severity (“severity-scaling” — a scaling factor for the probabilities of an infectious individual having severe symptoms {*p*_*ss,g*_}_*g*∈𝒢_), the social mixing parameter *α* (or equivalently, the elasticity of social contacts to activity levels7.), the cost of death *χ*, the ICU capacity *K*^ICU^, and the economic model parameters *ν*^work^, *ν*^other activities^, *ν*^fixed^. We create 14,039 problem instances by drawing each parameter independently and uniformly at random from a specified range. Details for the parameter variation are provided in Appendix EC.6.4 and in particular in Table EC.9.

#### Interpreting the dual-targeted policy

We create a training set of 5,524,785 samples — with one sample for every 14-day period, each age group *g*, and each activity *a* relevant to that age group. We fit a decision tree to predict the ROLD AGE-ACT-optimized activity levels 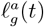 based on 20 features. We include as features several important problem parameters (e.g., the basic reproduction number *R*_0_, the cost of death, the elasticity of social contacts to activity levels, the ICU capacity) as well as derived features based on parameters and SEIR state values (e.g., econ-to-contacts-ratio, ICU utilization and admission rates, infection rates). Some of these features are targeted, meaning they take different values for different age groups or activities (e.g., econ-to-contacts-ratio), whereas others are non-targeted, such as *R*_0_, the time *t* or the cost of death *χ*. The details of the fitting procedure are in Appendix EC.6.4.1, and the full set of features is in Tables EC.10 and EC.11.

Figure 10 displays the depth-four tree obtained by fitting using the entire feature set, together with the resulting root-mean-squared-error (RMSE). This simple tree predicts the optimal ROLD activity levels reasonably well (RMSE of 0.31) and confirms our core insight that the econ-to-contacts-ratio is the most salient feature when targeting confinements, as it is used as a split variable in the root node of the tree and in several sub-trees, with higher ratios always leading to higher activity levels. The tree also confirms that *R*_0_ and time are relevant, with the optimal ROLD policy enforcing stricter confinements for larger values of *R*_0_ and in earlier time periods. Another relevant feature is R-perc, which quantifies the total number of individuals in a recovered state as a fraction of the overall population; this is a natural measure of the level of herd immunity and the ROLD policy leverages it intuitively, allowing more activity at larger levels of herd immunity.

**Figure 10.**
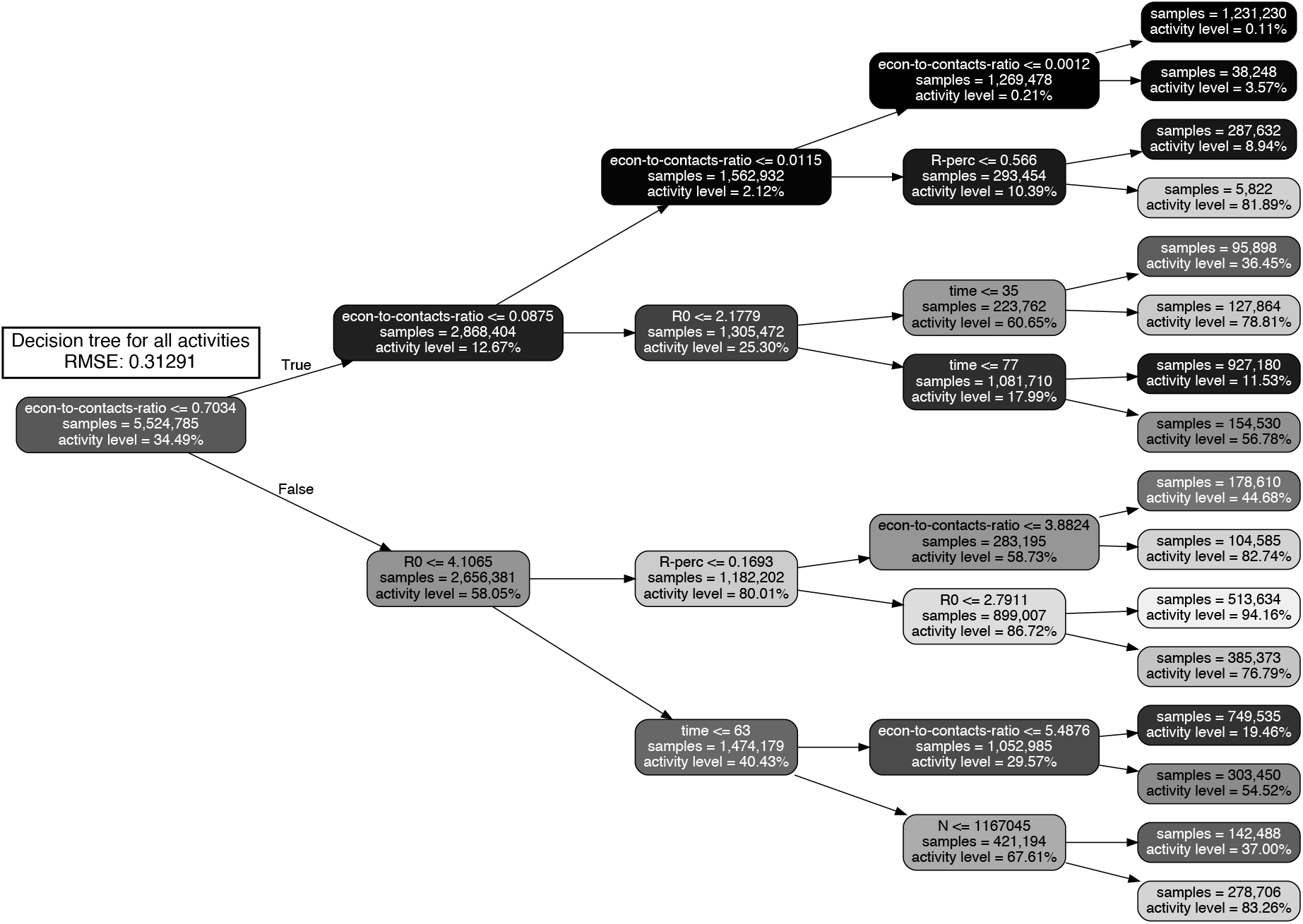
Decision tree of depth four approximating the optimized ROLD AGE-ACT decisions trained on 14,039 problem instances with an optimization horizon of *T*′ = 90 days (a total of 5,524,785 samples). Nodes are color-coded based on activity level, with darker colors corresponding to stricter confinement.

We also quantify the importance of the various features by calculating their permutation importance scores, a commonly used metric that measures importance as the increase in model prediction error after the values of the respective feature are randomly permuted. For trees of depth four, the scores for econ-to-contacts-ratio, *R*_0_, time, R-perc(*t*), and *N*_*g*_(*t*) are respectively 0.163, 0.065, 0.037, 0.008, 0.007; and all the other features have scores of zero. The econ-to-contacts-ratio feature thus has by far the largest permutation importance score of all features. Figure EC.6 in the Appendix confirms similar results for a tree of depth 10, and Figure EC.7 documents an additional exercise that further shows econ-to-contacts-ratio as the most salient of all targeted features. Appendix EC.7 contains additional results to complement this section, including a theoretical justification for the salience of the econ-to-contacts-ratio derived in a simplified model.

#### Interpreting the gains from dual targeting

For each of the 14,039 problem instances, we retrieve the optimal policies under ROLD AGE-ACT, ACT, and NO-TARGET, and calculate the relative gain of ROLD AGE-ACT over the less targeted policies ROLD NO-TARGET and ROLD ACT in terms of the total loss objective. We then fit decision trees to predict these relative gains. We include as features several problem parameters (e.g., the basic reproduction number *R*_0_, the cost of death, the disease severity, the elasticity of social contacts to activity levels, the ICU capacity) and derived features based on statistics of the econ-to-contacts-ratio across age groups and activities. Specifically, because each sample in the training set corresponds to an entire problem instance (including all time periods and all age groups and activities), we first calculate econ-to-contacts-ratio values at the start of the pandemic, denoted as time *t*_0_:

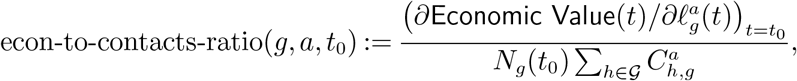

and use these to calculate two kinds of economic features: (i) for each activity other than home, we take the standard deviation of econ-to-contacts-ratio across all age groups that are active in that activity, which we label econ-to-contacts-ratio-activity-stdev; (ii) for each age group, we take the standard deviation of econ-to-contacts-ratio across all activities (other than home) in which that age group is active, which we label econ-to-contacts-ratio-agegroup-stdev. (Relevant activities for each age group are listed in Table EC.8.) The full set of features is summarized in Table EC.12 and the details of the fitting procedure can be found in Appendix EC.6.4.2.

To appreciate how the various features influence the gains, consider Figure 11, which displays permutation importance scores corresponding to depth-10 trees for explaining the gains of the ROLD AGE-ACT policy over ROLD NO-TARGET and ROLD ACT; and Figures EC.8 and EC.9 in the Appendix, which depict trees of depth five explaining the same gains.

**Figure 11.**
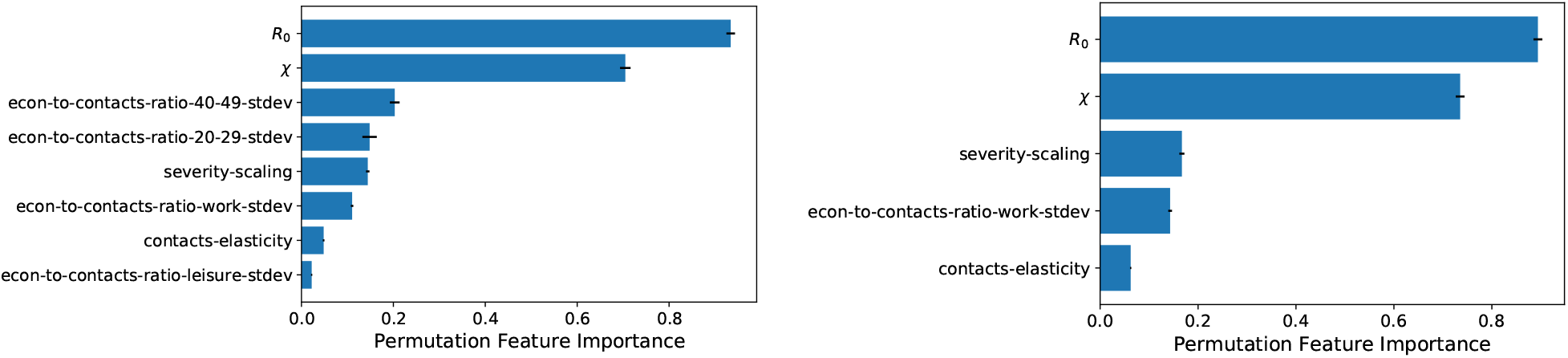
Permutation feature importance for the depth-10 decision trees explaining the relative gains of ROLD AGE-ACT over ROLD NO-TARGET (left) and ROLD ACT (right).

The most salient features for predicting the gains of dual targeting are *R*_0_, the cost of death *χ*, the heterogeneity in the econ-to-contacts-ratio values, the disease severity, and the elasticity of social contacts to activity levels. The two most important features, *R*_0_ and *χ*, impact gains in a similar, *non-monotonic* fashion, with moderate values leading to larger gains than very low or very high values. This can be best viewed in the partial dependence plots shown in Figure 12. This is explainable because all policies tend to enforce similar activity levels at very low or very high values of *R*_0_ or *χ* (that is, high activity values at sufficiently low values of *R*_0_ or *χ*, and vice-versa), which limits the benefits that could be generated through more refined targeting.

**Figure 12.**
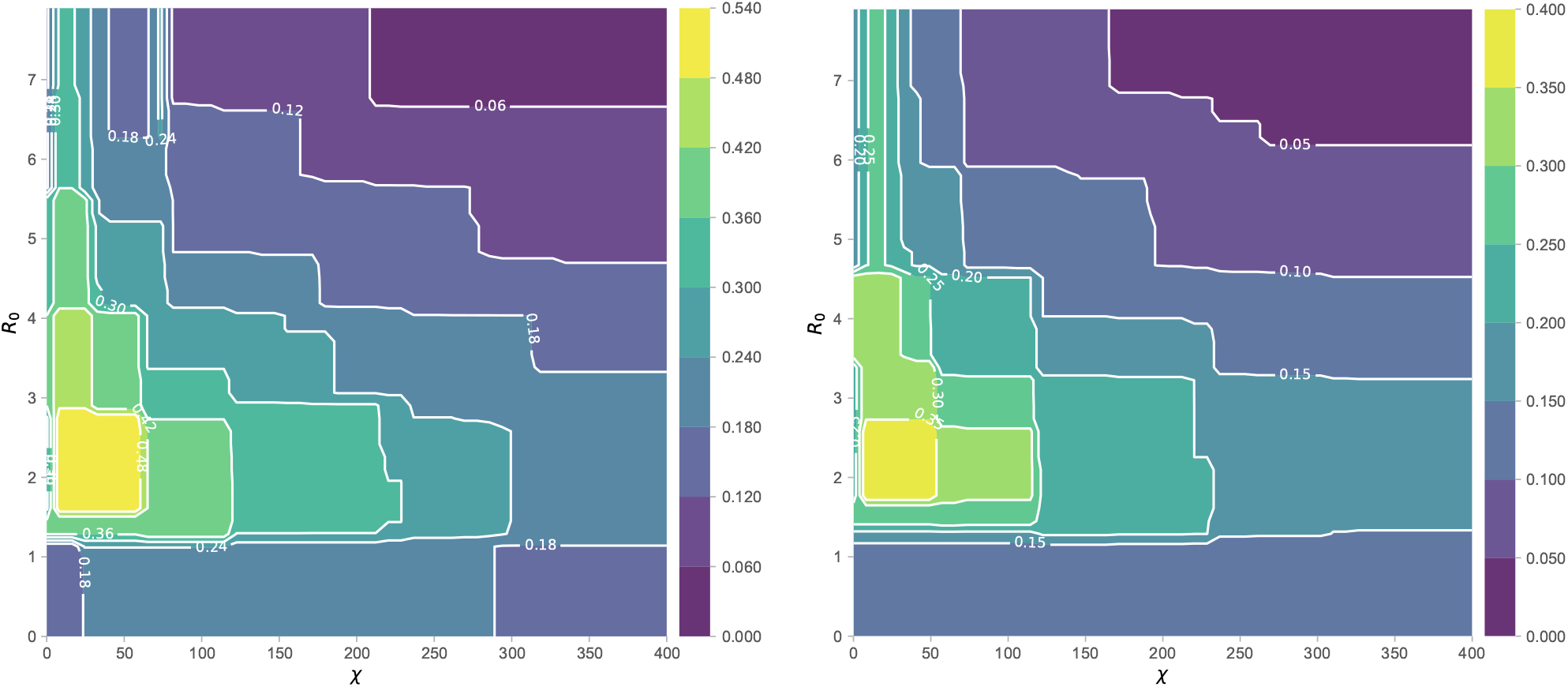
Partial Dependence Plots for the depth-10 decision trees showing the relative gains of ROLD AGE-ACT over ROLD NO-TARGET (left) and ROLD ACT (right) as a function of *R*_0_ and *χ*. The color scale depicts the magnitude of the relative gains. (The figure is best viewed in color.)

Figure 12 also leads to practical insights. Policy makers faced with pandemic variants with very high basic reproduction number *R*_0_ and who heavily prioritize health over economic outcomes should not concern themselves with targeting. The gains are also generally smaller in regimes with low *R*_0_ — although then, dual targeting could lead to more substantial gains for policy makers who weigh more heavily (albeit not too heavily) health over economic outcomes. Lastly, in regimes with moderate values of *R*_0_, all policy makers would gain substantially from targeting, and especially those who weigh economic outcomes more heavily.

The gains from dual targeting increase as the heterogeneity in the econ-to-contacts-ratio values (captured by the standard deviation) increases. This aligns well with the intuition developed in Section 7.1 that dual targeting exploits such heterogeneity to prioritize which (age group, activity) pairs to keep open. Intuitively, the gains relative to no-targeting are explained by heterogeneity across *both* age groups and activities, with the 40-49 and 20-29 y.o. emerging as the most relevant groups and work, leisure as the most relevant activities; in contrast, the gains relative to activity-based targeting are best explained by heterogeneity *across age groups* alone (in the work activity). Generally, the gains decrease as the disease severity increases, reflecting that a higher probability of severe symptoms limits the improvements available from finer targeting. In the regime of small *R*_0_, where the gains from targeting are limited, lower severity predicts even smaller gains. This is consistent with all policies recommending high activity levels, which leaves little room for improvements from targeting.

## 8. Towards Practically Implementable Policies

The previous sections established dual targeting as an idealized benchmark. However, discriminating activity levels based on age can be challenging in practice, from both an operational and an ethical/legal standpoint. We thus study two policy variations inspired by real-world interventions that are simpler to implement than dual targeting and that retain some of its superior health and economic outcomes.

In the first proposal, we consider *curfews* for different activities, implemented *uniformly* over all age groups. These draw inspiration from real-life policies during COVID-19: as an example, France implemented population-wide curfews during the first half of 2021 that started at a time varying from 6 p.m. to 11 p.m. and lasted until 6 a.m., while maintaining school and work activities largely de-confined (Reuters 2021). This is effectively implementing restrictions similar to ROLD AGE-ACT: under a 6 p.m. curfew, a typical member of the 20-65 y.o. group would be engaged in work until the start of the curfew, so their leisure and other activities would be quite restricted; in contrast, since most individuals aged above 65 are not in active employment, they would not face such restrictions in these last two activities. More generally, since separate age groups spend different parts of the day in different activities, curfews are a potential way to *implicitly* target age groups, but are operationally simpler and less contentious than the ROLD AGE-ACT policy.

In the second proposal, we consider *recommending*, as opposed to enforcing, targeted activity levels separately for each age group, while accounting for imperfect population compliance. Such recommendations have been deployed in the pandemic response of many countries, with governments encouraging specific at-risk groups such as the elderly to limit their activities outside of home (see the examples in Table EC.15). Such policies are thus more readily implementable than fully targeted confinements, and our goal with the exercise is to determine the compliance levels needed to ensure a sufficiently good performance relative to activity-based targeting.

### 8.1. Optimized Curfews

We focus on *practical* curfew policies: we do not allow targeting by age groups, we consider a single start and end-time within a 24-hour day (further constrained to occur at the hour or half-hour mark), we assume that activity levels are completely restricted during the curfew and unrestricted otherwise, and we allow curfews to change at most once every two weeks.

To formalize our model, we break the 24-hour day into 48 slots of 30 minutes each, starting at the hour and half-hour mark. For day *t*, we denote the *τ* -th time slot of the day by *t*_*τ*_, for *τ* = 1, …, 48, with *τ* = 1 denoting the 12 a.m.-12.30 a.m. time window. We encode the curfew for activity *a* as starting at the beginning of slot 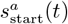 and ending at the end of slot 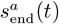. For each activity *a* ∈ 𝒜 \ {home}, we let *ℓ*^*a*^(*t*_*τ*_) denote the activity level allowed during time slot *τ* of day *t*, which we take to be 0 during the curfew and 1 outside of the curfew; we set *ℓ*^home^(*t*) = 1 throughout.

To model curfews, we refine our social mixing model to take into account the participation rates of different age groups in distinct activities throughout the day. To that end, we requested and obtained time-use survey data from the French National Archive of Data from Official Statistics (ADISP, INSEE 2010) that include detailed information of how survey participants of different ages spent every 10-minute interval of their day, from which we estimate participation rates in the different activities (details of our procedure are discussed in Appendix EC.8.1). Let 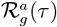 denote the participation rate of age group *g* in activity *a* during time slot *τ* of a generic day, so that 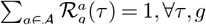. As in our base model, let *c*_*g,h*_(*ℓ*(*t*)) denote the mean number of total daily contacts between an individual in group *g* and individuals in group *h* across all activities during day *t*, when the activity levels are *ℓ* (*t*). We then set

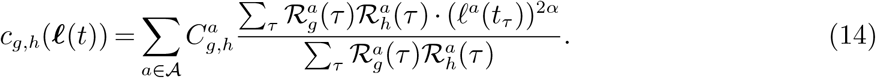

This expression extends our base-case social mixing model from Section 3 by capturing the distinct participation rates of the age groups through a weighted-average combination of contacts during the day. Note that if we removed the differentiation between time slots, i.e., 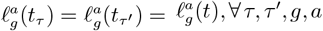, then (14) would collapse to our familiar social mixing model in (1) (for the case when all age groups have the same activity levels).

To determine the optimized curfew policy, which we henceforth refer to as CURFEW, we solve an optimization problem analogous to the one in (8)-(10), with the following modifications. First, the decision variables are the “start” and “end” time of the curfew in each activity on each day *t*, 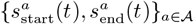, allowed to change once every 14 days. Second, the calculation of Economic Loss in the objective replaces *ℓ*(*t*) in (7) with *ℓ*^eff^(*t*), where 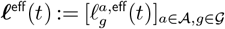 and

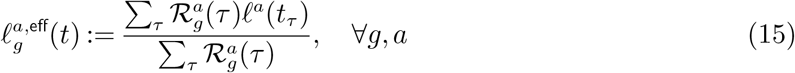

denotes the (weighted-average) effective activity level of group *g* in activity *a* during day *t*, accounting for the group’s participation rates. Third, the SEIR dynamics in constraint (9) are modified to reflect the revised social mixing in (14). Lastly, we solve the optimization problem through a gradient descent procedure, and round its optimal solution to the closest hour or half-hour mark (details are provided in Appendix EC.8.2).

Figure 13 shows that optimized curfews can significantly outperform ROLD ACT in terms of the total loss objective: CURFEW closes 24.3% (median value) of the performance gap between ROLD AGE-ACT and ROLD ACT, and the improvement can reach up to 65.1% of the gap for moderate values of the cost of death *χ*, namely *χ* = 145. As a side remark, we conjecture that these improvements are conservative estimates of what is achievable because the optimization procedure used for CURFEW is significantly less sophisticated than the one used for ROLD AGE-ACT.

**Figure 13.**
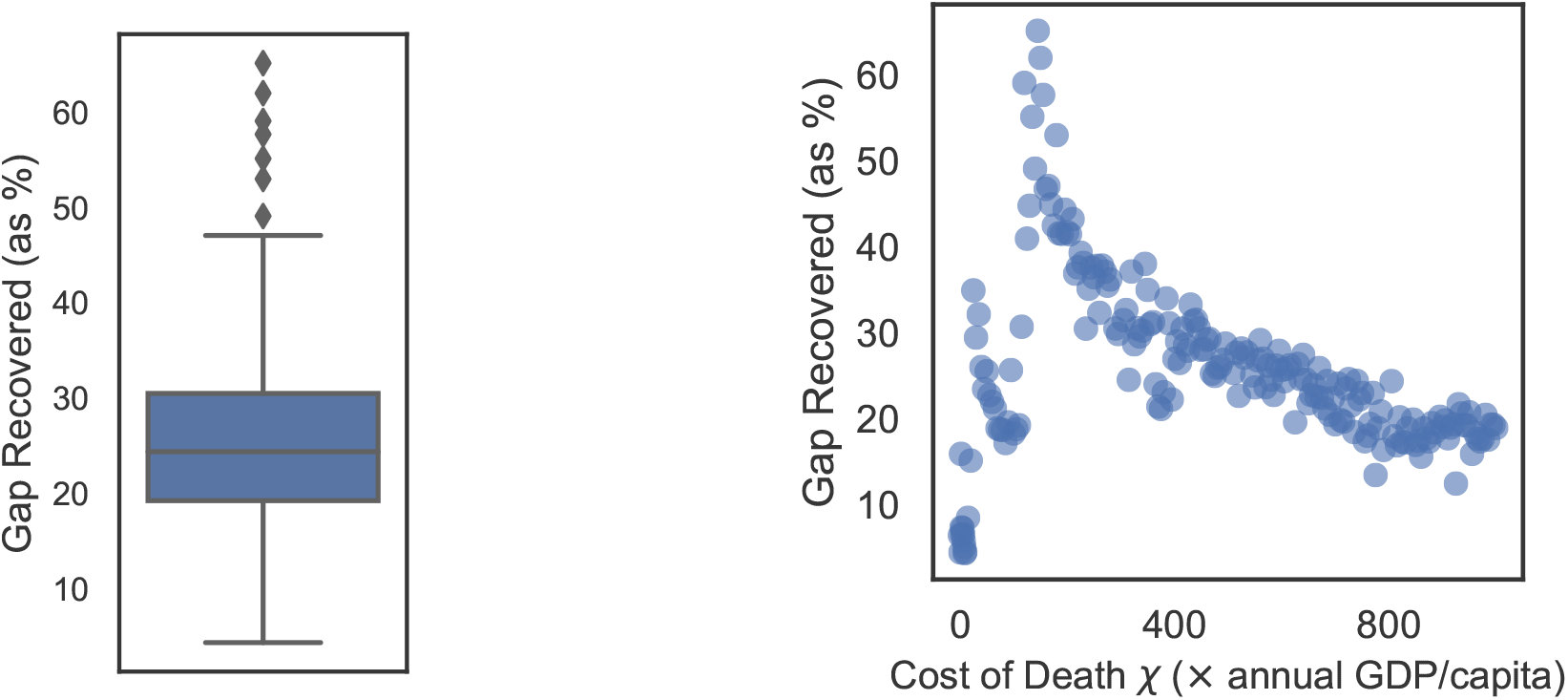
The performance of the CURFEW policy expressed as a fraction of the gap in total loss between ROLD ACT and AGE-ACT that it recovers. We include 199 distinct values of *χ*, from 0 to 1000×.

That CURFEW can recover a substantial portion of AGE-ACT’s performance despite not explicitly targeting age groups suggests that time-of-day targeting implicitly allows some differentiation by age. To examine this, we calculate the time-average activity levels of all the relevant age groups engaged in a given activity (per Table EC.8), based on which we determine the range of activity levels encountered across groups. Figure 14 plots the magnitude of these ranges for the CURFEW policy. When comparing these with ROLD ACT, which by definition has zero range in each activity, it can be seen that CURFEW effectively differentiates restrictions by age in each activity.

**Figure 14.**
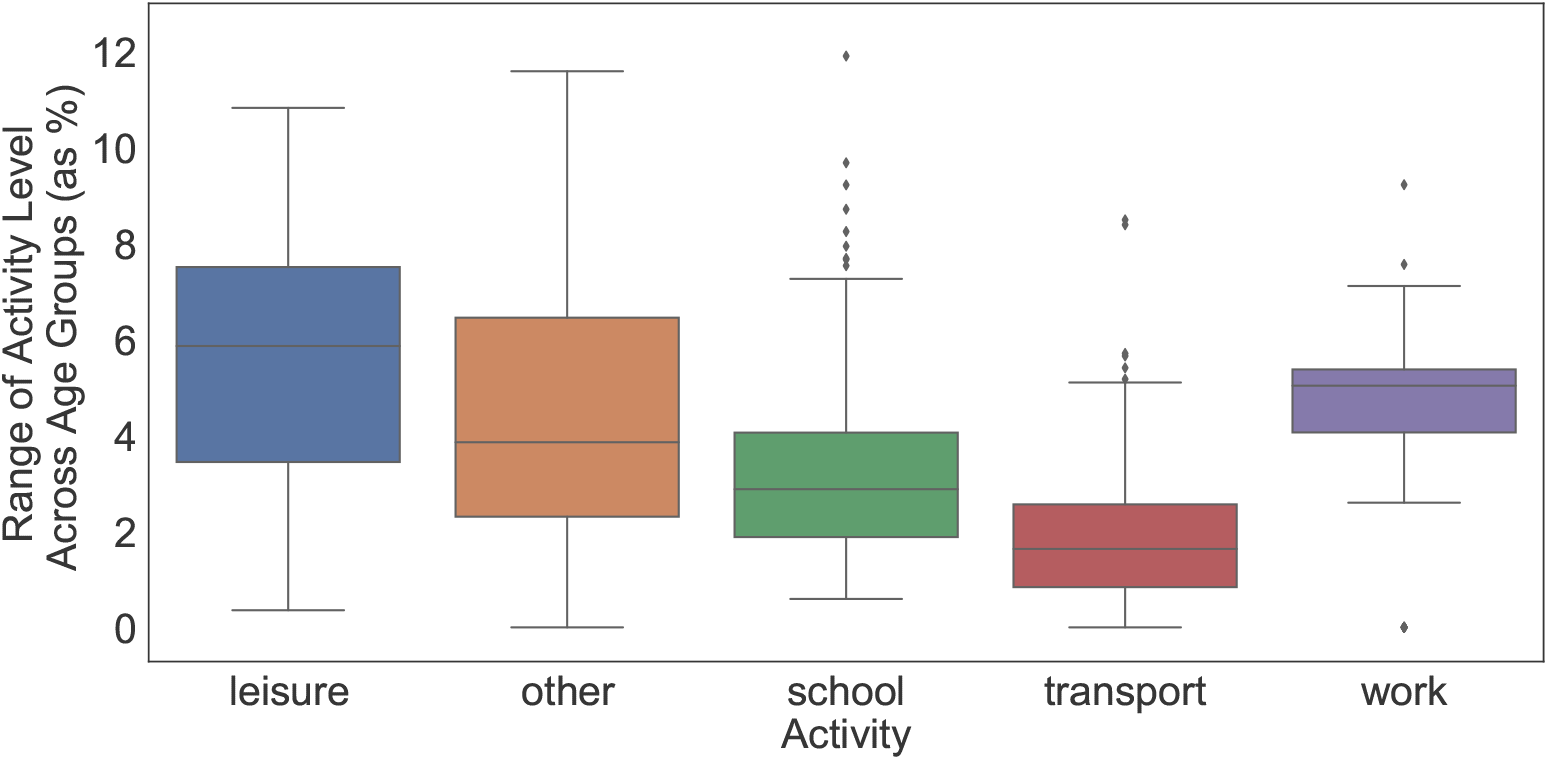
The range of time-average activity levels (defined as maximum minus minimum level over all relevant age groups) for the CURFEW policy, in several problem instances parametrized by *χ*. In comparison, the ROLD ACT policy has equal activity levels for all age groups, and thus zero range for each activity.

Overall, these results highlight the benefits of the CURFEW policy, which can significantly improve performance relative to ROLD ACT without incurring the operational or ethical implementation challenges of ROLD AGE-ACT.

### 8.2. Dual-Targeted Recommendations with Limited Compliance

We next examine a practically-appealing implementation of dual targeting that phrases ROLD’s output as *recommendations* instead of mandatory restrictions and that also accounts for imperfect compliance, whereby only a subset of the population would follow the recommendations. More formally, we capture compliance through a parameter *p*, with 0 ≤ *p* ≤ 1, such that a *p* fraction of the population of each age group adheres to the recommended age group-targeted activity levels, and the remaining 1 − *p* fraction does not adhere, but must still abide by a population-wide restriction in each activity (which is assumed enforceable, as in ROLD ACT). We refer to the resulting policy as the ROLD COMP(*p*) policy.

More precisely, ROLD COMP(*p*) still optimizes for *ℓ* via the optimization problem (8)-(10), but builds in limited compliance into this optimization by adding the following constraints:

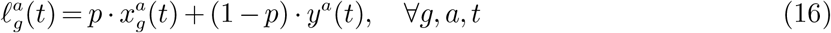

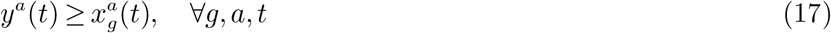

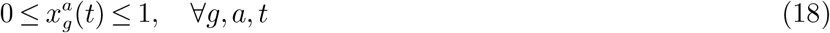

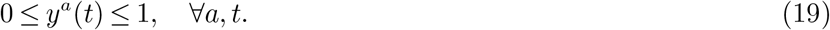

This confinement policy can be interpreted as a linear combination of two policies: an age-and-activity targeted policy, 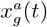, which the policy maker can only recommend; and an activity-only targeted policy, *y*^*a*^(*t*), which the policy maker enforces. The linear weights reflect the percentage of the population that would be compliant with the recommendation 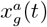, versus those who would only abide by the enforceable activity levels *y*^*a*^(*t*). Constraint (17) then expresses that the enforceable activity level cannot be more restrictive than the recommended one.

Figure 15, which shows ROLD COMP(*p*)’s performance for different compliance levels *p*, confirms the practical promise of the approach. The left panel, which depicts the fraction of the performance gap (in terms of the total cost objective) between ROLD ACT and AGE-ACT recovered by ROLD COMP(*p*), suggests that significant improvements are already available at relatively modest compliance levels. Specifically, the median gap recovered is 10.4% at 40% compliance and 19.2% at 60% compliance. As *p* increases, the performance naturally improves, and the relationship between performance and compliance level is overall convex, with larger marginal gains occurring at higher compliance levels.8. The right panel suggests that ROLD COMP(*p*) Pareto-dominates ROLD ACT even for small compliance levels when restricting to moderate values of *χ*, and Pareto-dominates over all tested *χ*’s for sufficiently large levels of compliance. We also consider a variation of limited compliance in which we allow each age group to have a group-specific compliance fraction calibrated from empirical data (Ganslmeier et al. 2022) and the resuls are very similar (Appendix EC.8.4).

**Figure 15.**
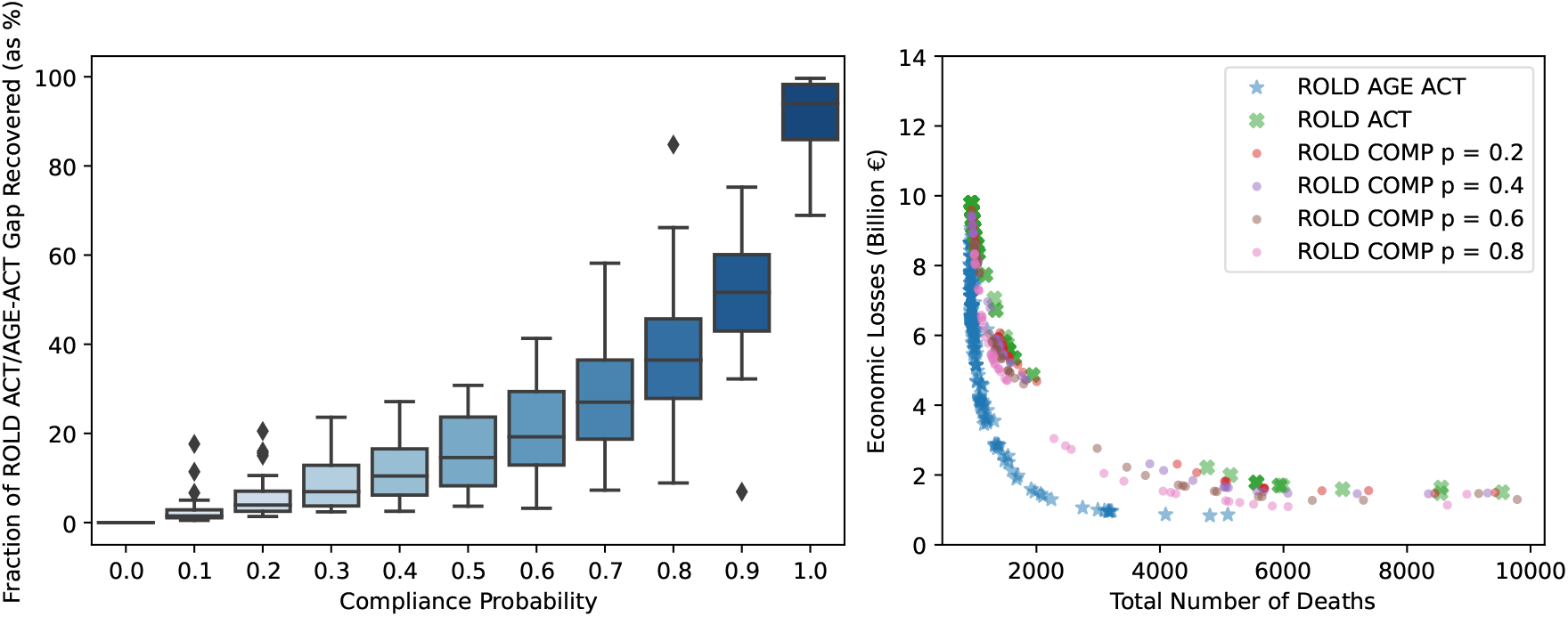
The performance of limited compliance policies for different compliance levels. In the left panel, each boxplot summarizes the fraction of the ROLD ACT/AGE-ACT performance gap (in terms of the total cost objective) that ROLD COMP(*p*) recovers, in 201 problem instances parametrized by the cost of death *χ* (with values from 0 to 1000×). The right panel illustrates the total number of deaths and the economic losses generated by ROLD COMP(*p*) for different compliance levels *p*, compared to ROLD AGE-ACT and ROLD ACT; each marker corresponds to a different value of *χ*.

These results suggest that *recommendations* for reducing activity levels according to the dual-targeted ROLD AGE-ACT policy, accompanied by reasonable levels of compliance in the population, can lead to significant health and economic benefits compared to less targeted policies.

## 9. Discussion

Our study established that dual targeting could in principle lead to significant benefits and that certain simple, practical policies can achieve some of these benefits. Dual targeting can improve both health *and* economic outcomes and it can maintain higher activity levels for most age groups (and particularly for those most confined), thus making confinements more socially acceptable. Such benefits are generated by heavily leveraging complementarities through a simple “bang-for-the-buck” rule: impose less confinement on group-activity pairs with a high econ-to-contacts-ratio, i.e., high marginal economic value prorated by activity-specific social contacts. The gains afforded by targeting are generally largest for moderate values of the basic reproduction number *R*_0_ and the cost of death, high heterogeneity in the econ-to-contacts-ratio, and low disease severity. Because dual targeting can face significant implementation challenges, we introduced two practical proposals — based on curfews and recommendations — that achieve some of the benefits without explicitly discriminating based on age.

### Future work

The algorithmic developments in our proposed framework for quantifying the possible benefits of dual targeting could be improved in several ways. One is end-of-horizon effects: ROLD may allow for increased infections as it nears the end of the horizon, since these may not progress to deaths. A policy maker may want to adjust ROLD’s behavior to prevent this, for example through a thorough sensitivity analysis of the optimal policy with respect to the problem horizon, or by imposing penalties on the remaining infections, or by running ROLD in a rolling (non-shrinking) horizon fashion. Secondly, as seen with emerging COVID-19 variants, the underlying epidemic environment could change during a typical ROLD time horizon. To track and respond to such changes, a potential approach could be to re-estimate the model parameters on the fly as ROLD advances, or to adopt a robust model that explicitly allows for mis-specification. One could also consider second-order approximations of the dynamics and objective and leverage (non-convex) quadratic programming to deal with the resulting optimization problems. Lastly, work could also be devoted to deriving a theoretical justification for the importance of the econ-to-contacts-ratio by generalizing our simple analysis in Appendix EC.7, or by deriving tractable upper bounds for the performance of controlled SEIR models that could be used to benchmark interventions.

Our modeling choices were constrained by available data, but further data collection may allow for more precise modeling. Our social mixing model relied on social contact matrices by age group and activity, which are available from surveys conducted in a number of countries; additional data may allow refining the population group or activity definitions. Additionally, the available social contacts data only record interactions in the same activity, whereas contacts occur as individuals are engaged in different activities (e.g., a services industry professional interacts during work with individuals who are engaged in leisure activities). A more refined mixing model that captures such interactions would be desirable, provided that data are available to calibrate it. Similarly, we worked with economic data reported by industry activities and for coarse population groups, but a dataset that splits economic value into fine-grained (group, activity) contributions would be more suitable. Lastly, a more detailed economic model would include cross effects, e.g., children in school producing value in conjunction with educational staff engaged in work.

Although we focus on confinements, a direction for future research is to see how to combine these with other types of targeted interventions. The framework is sufficiently flexible to accommodate interventions such as contact tracing, mass testing and also vaccinations, although a careful implementation is beyond the scope of this article.

Finally, to address fairness concerns related to targeting in a principled way, one could impose fairness requirements based on the interventions’ actions and/or outcomes, e.g., requiring that the health or economic losses or the time in confinement faced by different groups should satisfy certain axiomatic fairness properties (Young 1994).

## Data Availability

The time-use survey data was obtained after special request from the French National Archive of Data from Official Statistics (ADISP). All other data referred to in the manuscript are public and URL references to the datasets are included in the manuscript.

**This page is intentionally blank. Proper e-companion title page, with INFORMS branding and exact metadata of the main paper, will be produced by the INFORMS office when the issue is being assembled**.

## E-companion to Quantifying and Realizing the Benefits of Targeting for Pandemic Response

### EC.1. Dynamics of the Controlled SEIR Epidemic Model

We write down a set of discrete time dynamics for the controlled SEIR model. We use notation Δ*Z*(*t*) to denote *Z*(*t* + 1) − *Z*(*t*). For all groups *g* ∈ 𝒢, we have:

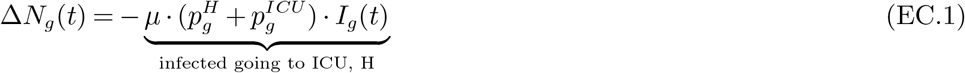

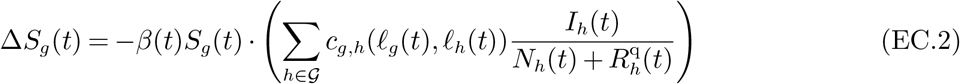

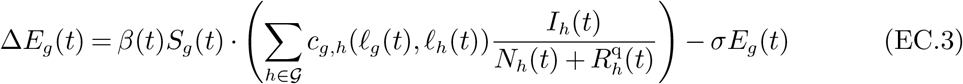

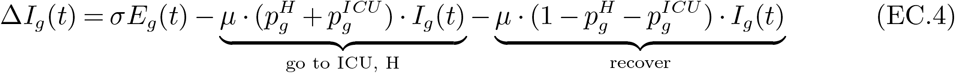

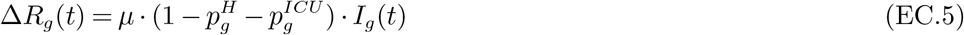

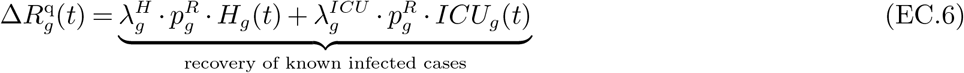

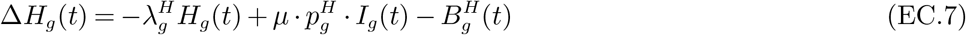

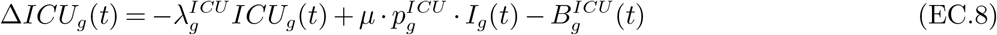

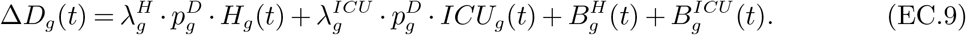

In (EC.5) and (EC.6), note that we do not have terms for the population turned away from hospital/ICU which may eventually recover. Instead, we assume the turned away patients will go into the deceased state. In (EC.9), we are assuming that if a patient is turned away from the ICU, they transition into deceased, instead of being allocated a hospital bed if one is available.

We now provide justification for how we account for social contacts and, in particular, for the expressions in (EC.2) and (EC.3). We note that individuals in 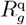 can interact with members of *S*_*g*_, *E*_*g*_, *I*_*g*_ and *R*_*g*_. Fix a person *i* in age group *g* ∈ 𝒢, in state *S*_*g*_. Then:

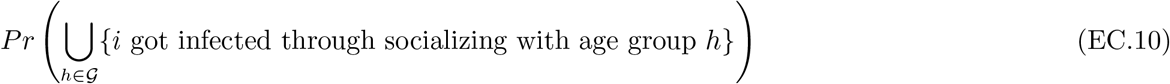

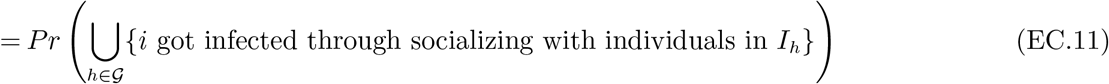

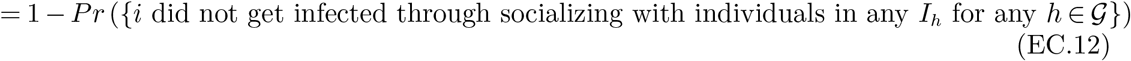

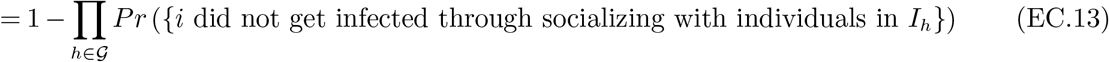

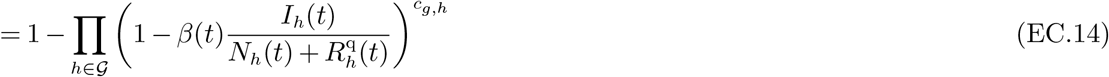

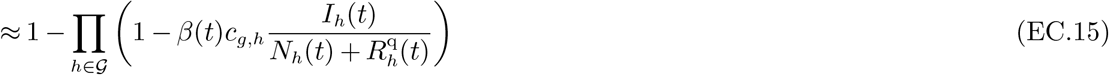

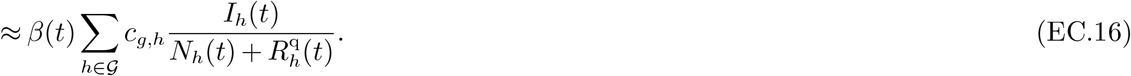

In (EC.14) we use the following reasoning. Having fixed person *i* in age group *g*, (a) a contact with a randomly chosen individual in group *h* will result in person *i* getting infected with probability 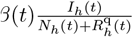, and (b) the number of person *i*’s contacts with individuals in age group *h* is given by *c*_*g,h*_ = *c*_*g,h*_(*ℓ*_*g*_(*t*), *ℓ*_*h*_(*t*)). Finally, person *i* getting infected as the result of a contact with someone from group *h* is considered to be an independent event across different contacts, therefore we raise the probability of no infection from a contact to the power of the number of contacts. (EC.15) and (EC.16) follow from linear approximations.

By taking the expectation of random variable

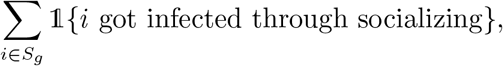

we retrieve the expressions in (EC.2) and (EC.3).

### EC.2. Details of the Economic Model

As discussed in Section 3.4, economic losses come from three separate sources:

#### Effect of confinement

To account for confinement in the non-quarantined population, we make the economic value generated per day by an individual in group *g* in the remaining (non-quarantined) SEIR chambers explicitly depend on the enforced confinement in the population. Recall that for a group *g*, the activity levels *ℓ*_*g*_ specify the level of each activity allowed for that group as compared to normal course, and *ℓ* = [*ℓ*_*g*_]_*g*∈𝒢_. We denote the economic value generated by a member of *g* per day by *v*_*g*_(*ℓ*). We remark that *v*_*g*_(**1**) corresponds to the economic value generated by an individual under normal circumstances.

The *v*_*g*_(*ℓ*) specific to a group can be of two types: (a) wages from employment and (b) future wages from employment due to schooling. Naturally, depending on the age group, both, one, or neither of these will actually contribute to economic value. Distinguishing whether the specific group is comprised of school age, employable or retired population, we define

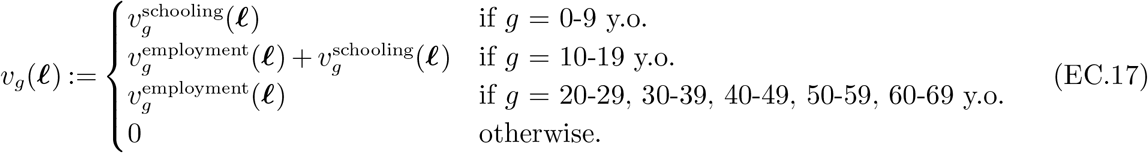

We break down the definitions of 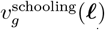 and 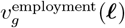 below:

- **Value from employment** 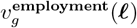. The value generated from employment is a function of the confinement level in the work activity, but also of the confinement levels in leisure, transport, as well as other activities. As an example, we expect the economic value generated by those employed in restaurants, retail stores, etc. to depend on foot traffic levels, which in turn are driven by the confinement levels in leisure, transport and other activities across all groups.

Our model for employment value is a linear parametrization of these confinement decisions; specifically, 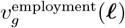 is linear in 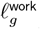 and the weighted average of *ℓ*^transport^, *ℓ*^leisure^ and *ℓ*^other^ across these three activities and all groups *g* ∈ *𝒢*:

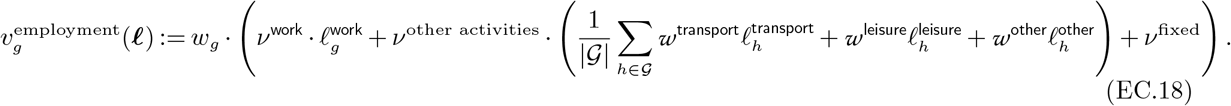

Additionally, *ν*^work^, *ν*^other activities^ and *ν*^fixed^ are activity level sensitivity parameters such that *ν*^work^ 1 + *ν*^other activities^ 1 + *ν*^fixed^ = 1; under fully open activity, they induce a multiplier of 1 in (EC.18). Then *w*_*g*_ measures the overall daily employment value of a typical member of group *g* under no confinement, and is equal to 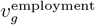(**1**). The weights *w*^transport^, *w*^leisure^, *w*^other^ capture the relative importance of each of these three activities for employment value. For our baseline setting, we take *w*^transport^ = *w*^leisure^ = *w*^other^ = 1*/*3. We let these parameters vary in our analyses in Section 7.2.

We estimate the coefficients of this model from data, as we describe in detail in Section EC.4.

- **Value from schooling** 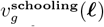. A day of schooling for the individuals in relevant groups results in economic value, equal to a day of wages that members of these groups would gain in the future. We use the salary of the 20-29 year-old group multiplied by a factor, and we discount for a number of years corresponding to the difference between the midpoint of the age group and the beginning of the 20-29 year-old group. For instance, for the 0-9 year olds we discount over 15 years, and for 10-19 year olds we discount over five years. The discounting factor we apply is thus

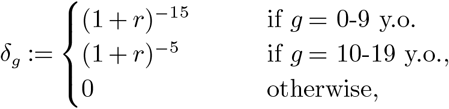

where *r* is the discount rate. We further multiply the wage by *f*_*g*_, which is the fraction of group *g* that is in school.9. Lastly, we also use a multiplicative factor *θ* for sensitivity analysis: *θ* reflects that an additional day of schooling may have a multiplier effect in future wages, as well as the fact that schooling can be continued online during lockdowns. We provide ranges for *θ* in Section 5.3.

Thus, the definition for value of school days is

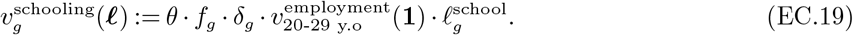

#### Effect of quarantine, illness and deaths during the pandemic

We capture the economic effect of quarantine, hospitalization, and death during the pandemic by assuming that if at some time period an individual in group *g* is in one of the SEIR chambers *H*_*g*_, *ICU*_*g*_, *D*_*g*_, then they generate no economic value. At the same time, we assume that individuals in 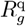 generate economic value as they would under no confinement.

#### Effect of lost future wages due to death

We account for a deceased individual’s lost wages which they would have earned from their current age until retirement age, given the prevailing wage curve under normal circumstances, {*v*_*g*_(**1**)}_*g*∈𝒢_. For group *g*, we set the current age to the midpoint of the age group. We discount the resulting cash flows by an annualized interest rate. We denote the resulting lost wages amount by 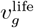.

For instance, for someone in age group 30-39 y.o., we calculate this cash flow by10.

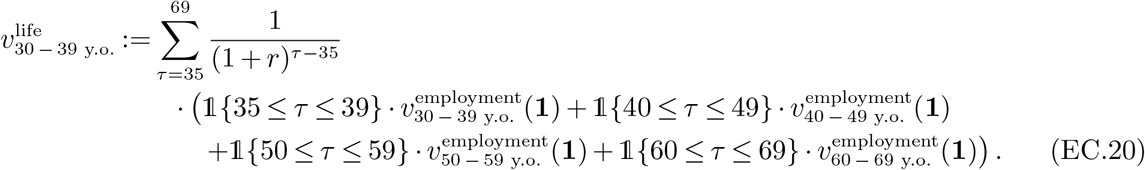

Last, we define a quantity *V* which represents the economic value that would be generated across all groups *g* ∈ 𝒢, during the time of the pandemic, under a “no pandemic” scenario. More precisely, to calculate *V* we assume that at time *t* = 0 all the infected population is instantaneously healed and able to generate full economic value *v*_*g*_(**1**). Thus,

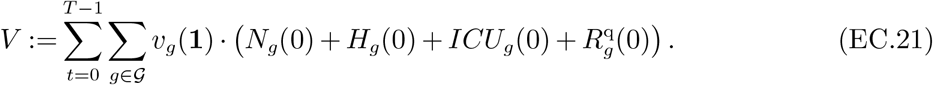

Note that this term is a constant and does not depend on the policy followed by the policy maker.

### EC.3. Algorithmic Details for ROLD

In this section, we clarify the algorithmic details of the linearization-optimization procedure described in Section 4. We first focus on how we build a linear model given *k*, ***X***_*k*_ and **û**_*k*:*T* −1_.

#### EC.3.1. Linearized Dynamics

In Step 2, the algorithm builds an approximation of the state dynamics that is linear in the controls ***u***_*k*_, …, ***u***_*T* −1_. Here, we compute the coefficients for each ***u***_*t*_ explicitly. We introduce the notation:

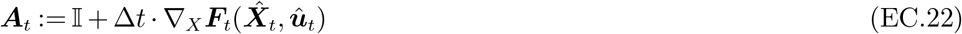

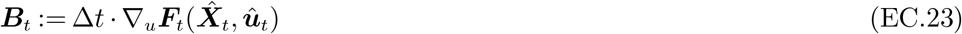

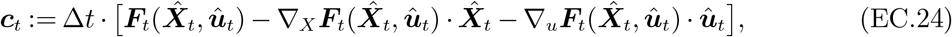

where matrix ***A***_*t*_ has dimensions |𝒢||*𝒳*| *×* |𝒢||*𝒳*|, matrix ***B***_*t*_ has dimensions |𝒢||*𝒳*| *×* |𝒢|| *𝒰*|, and vector ***c***_*t*_ has dimensions |𝒢||*X* | *×* 1. With this, we have

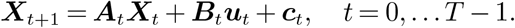

We can then express the state ***X***_*t*_ as11.

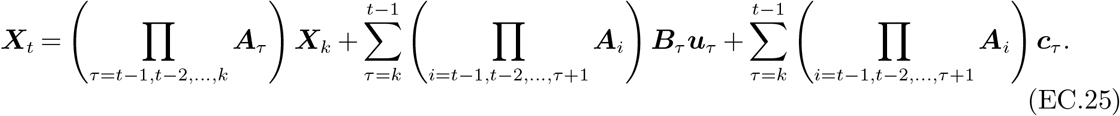

It is now possible to express both the objective and the constraints linearly in the decisions ***u***_*t*_.

#### EC.3.2. Constraint Coefficients

We can write each of the constraints (4) and (5) in the form

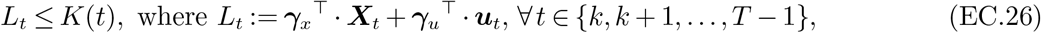

for some (time-invariant) ***γ***_*x*_, ***γ***_*u*_. Since ***X***_*t*_ is linear in ***u***_*k*_, …, ***u***_*t*−1_, to represent one such constraint we just need to store the coefficients corresponding to all decision variables (i.e., ***u***_*k*_, …, ***u***_*T* −1_) and the free terms/constants that appear in *L*_*t*_.

In particular, in the LHS ***γ***_*x*_^⊤^*·* ***X***_*t*_ + ***γ***_*u*_^⊤^*·* ***u***_*t*_ of such a constraint, the decision ***u***_*τ*_, for *k* ≤*τ* ≤*t*, will have coefficients

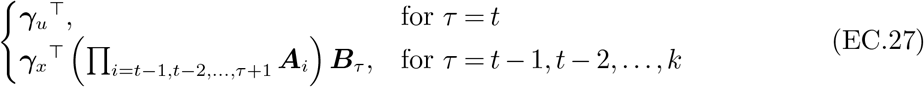

To make calculations efficient, we note that the coefficients can be obtained recursively as in the CalculateConstraintCoefficients function defined in Algorithm 1.

#### EC.3.3. Objective Coefficients

Up to constants, the objective in (13) can be written as

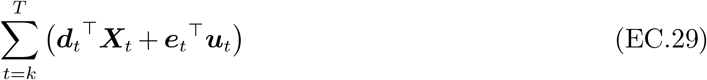

with

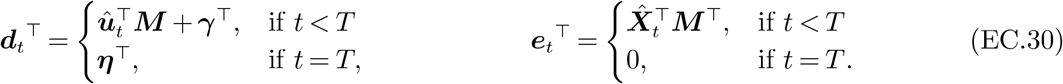

##### Algorithm 1

CalculateConstraintCoefficients

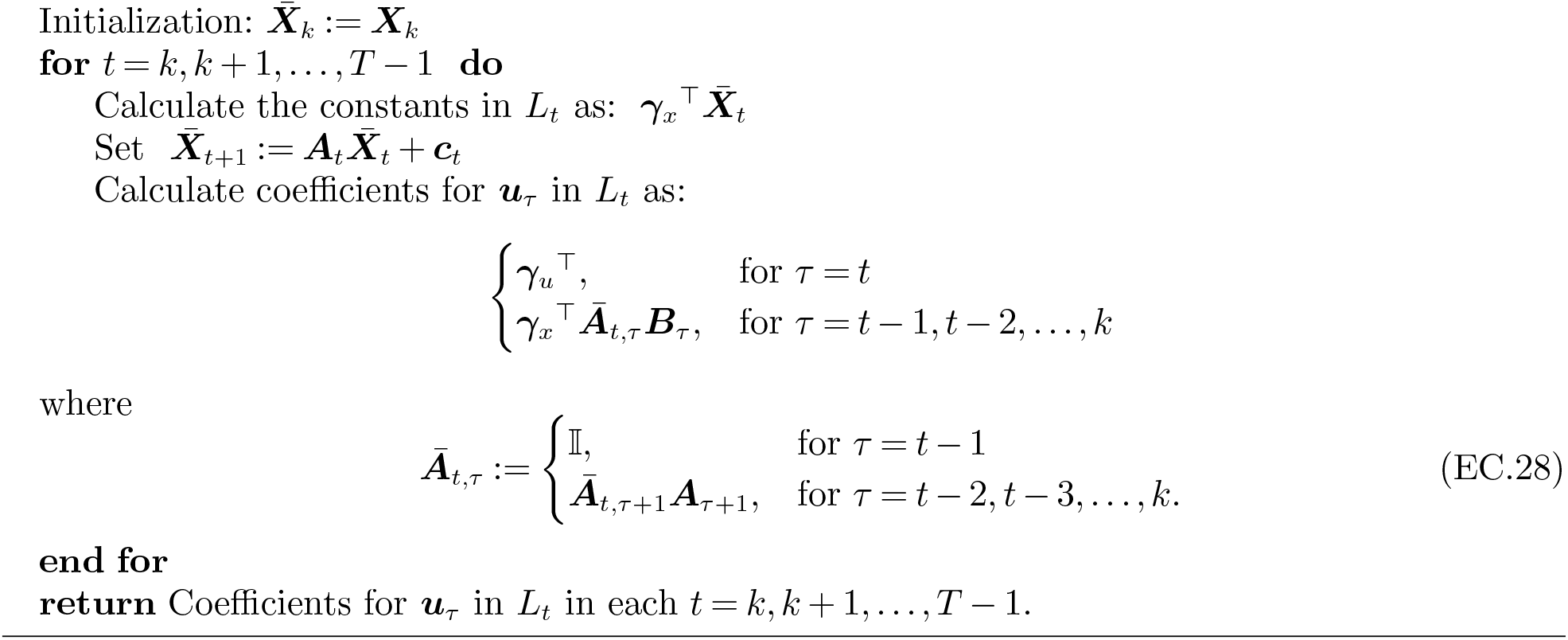

In (13) the decisions ***u***_*t*_, for *k* ≤ *t* ≤ *T* − 1, will have objective coefficients:

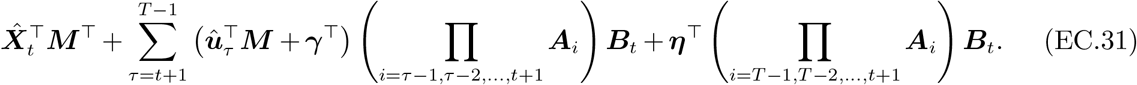

This allows calculating the coefficients recursively, just as we did for the constraints. The detailed function CalculateObjectiveCoefficients is defined in Algorithm 2.

##### Calculation of *M, γ* and *η*

Expanding the objective (8), we have:

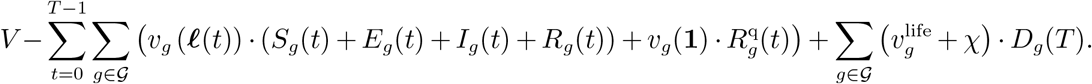

From this equation and the definitions of *v*_*g*_ (*·*) in Appendix (EC.2) above, we can write ***M*** (where the rows are indexed by the controls and the columns indexed by the compartments) as

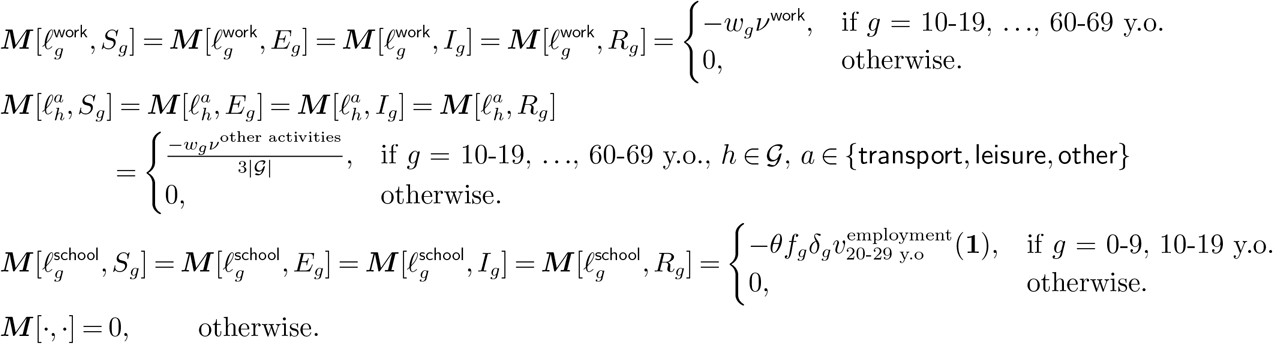

###### Algorithm 2

CalculateObjectiveCoefficients

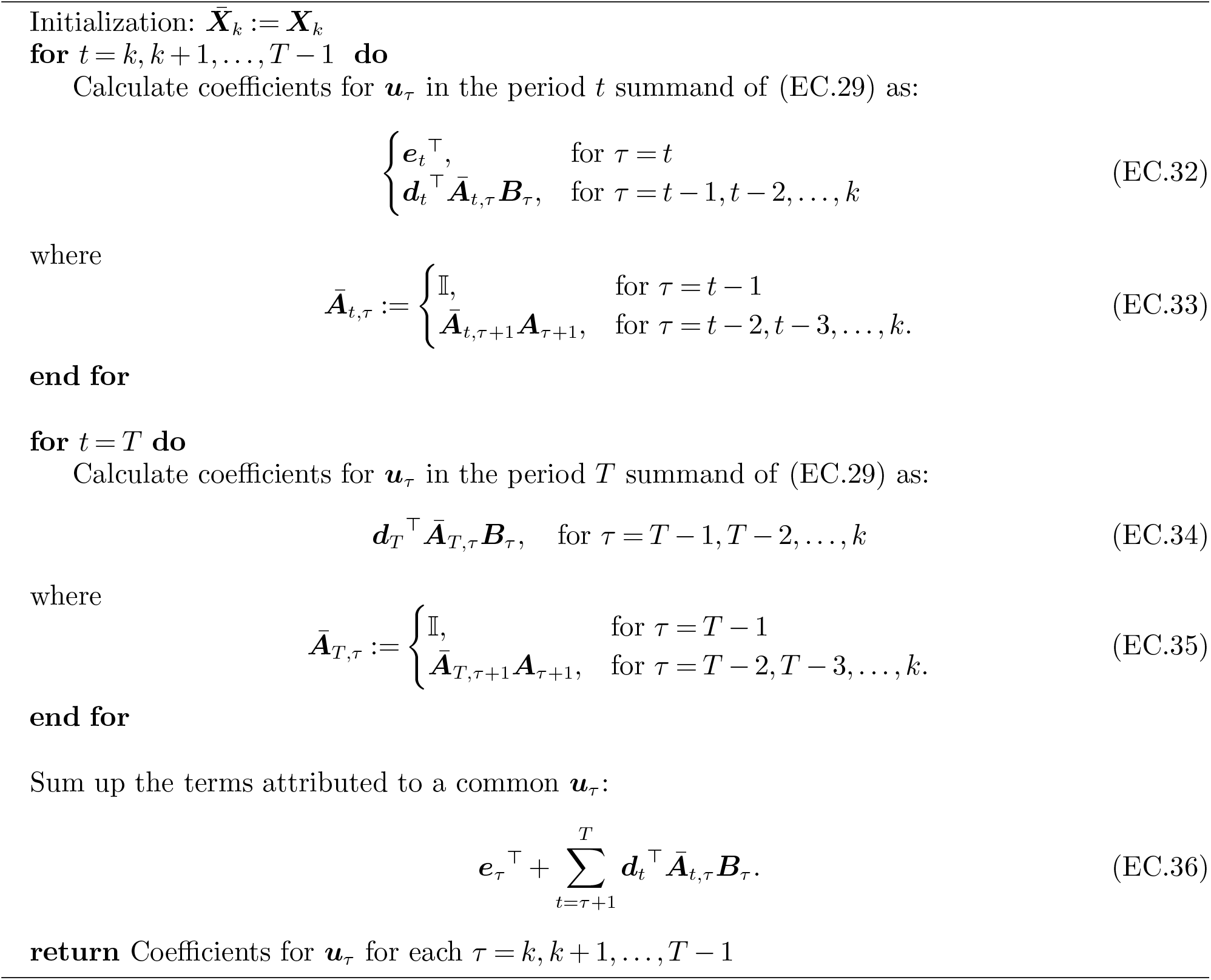

Similarly, we can write ***γ*** (indexed by the compartments for each group) as

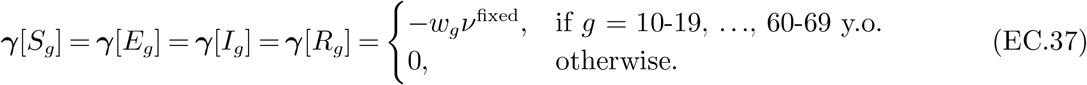

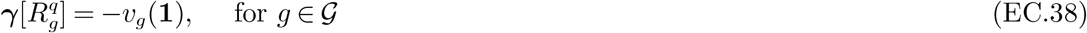

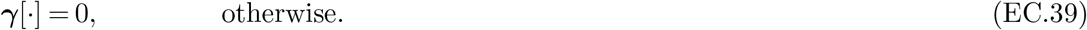

Finally, ***η*** (indexed by the compartments for each group) is

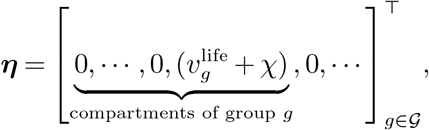

where the only non-zeros are in the indices corresponding to compartment *D*_*g*_ of each group *g*.

#### EC.3.4. Specifics of the Iterative Linearization-Optimization Procedure

Having defined functions CalculateConstraintCoefficients and CalculateObjectiveCoefficients, the Linearization-Optimization function which is the main subroutine of ROLD is described in Algorithm 3. This function builds the linear approximation for the remaining trajectory of the system, and optimizes it via an LP in a trust region of an infinity-norm *ϵ*-ball around the initial nominal control 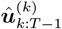. We denote that *E*-ball by 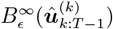.

##### Algorithm 3

Linearization-Optimization

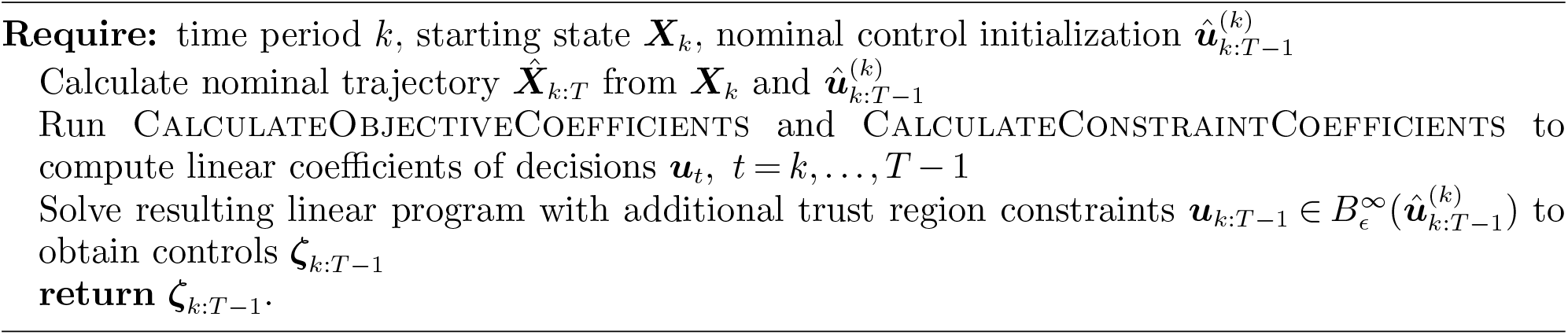

The full ROLD procedure is given in Algorithm 4. Within each period *k*, ROLD calls the Linearization-Optimization function iteratively up to a termination condition, using the output control to initialize the nominal control and the trust region for the next call of the function. This still requires to choose an initialization of the *k* = 0 nominal control 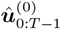; in our experiments we initialize this with a heuristic solution generated via a gradient method.

For the termination condition, we combine a fixed upper bound on the number of iterations with a condition that we do not repeat the control sequences produced by Linearization-Optimization, in order to avoid cycles. The fixed upper bound on the number of iterations is set so as to ensure that for each *k*, every confinement decision in ***u***_*k*:*T* −1_ can be changed to any value in [0, 1] with *ϵ*-length steps, i.e., the upper bound is at least 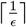. We further multiply the allowed number of iterations by a multiple *mult* ≥ 1, fixing the upper bound to be *mult* 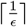.

We experimented with different values of *E* between 0.01 and 0.5 and values of *mult* between 1 and 5. As expected, lower values of *ϵ* resulted in a more stable and higher performing heuristic. Higher values of *mult* improved the heuristic only up to around *mult* = 2, after which point the non-cycling termination condition was triggered almost always. On the other hand, reducing *ϵ* had a significant impact on the run-time of the linearization algorithm. We chose the combination of *ϵ* and *mult* that gave us the best trade-off between the quality of the solution and the total run-time. In particular, for all the runs presented we take *ϵ* = 0.05 and *mult* = 2, resulting in an upper bound of 40 runs for the inner loop.

#### EC.3.5. Initialization for ROLD

In this subsection we flesh out the details for the nominal control initialization 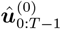. Our implementation of the initialization varies depending on the ROLD variant (i.e., the level of targeting):

- ROLD NO-TARGET: ROLD is initialized at a solution of a gradient descent method that is constrained to have the same activity levels across all age groups and activities, which can vary through time.
- ROLD AGE: A gradient descent method that is constrained to have the same activity levels across all activities for a given age group (and the activity levels are allowed to vary through time) is initialized at the ROLD NO-TARGET solution. Then ROLD is initialized at the solution of the gradient descent method.
- ROLD ACT: A gradient descent method that is constrained to have the same activity levels across all age groups in a given activity (and the activity levels are allowed to vary through time) is initialized at the ROLD NO-TARGET solution. Then ROLD is initialized at the solution of the gradient descent method.
- ROLD AGE-ACT: We keep the best solution out of:

##### Algorithm 4

ROLD

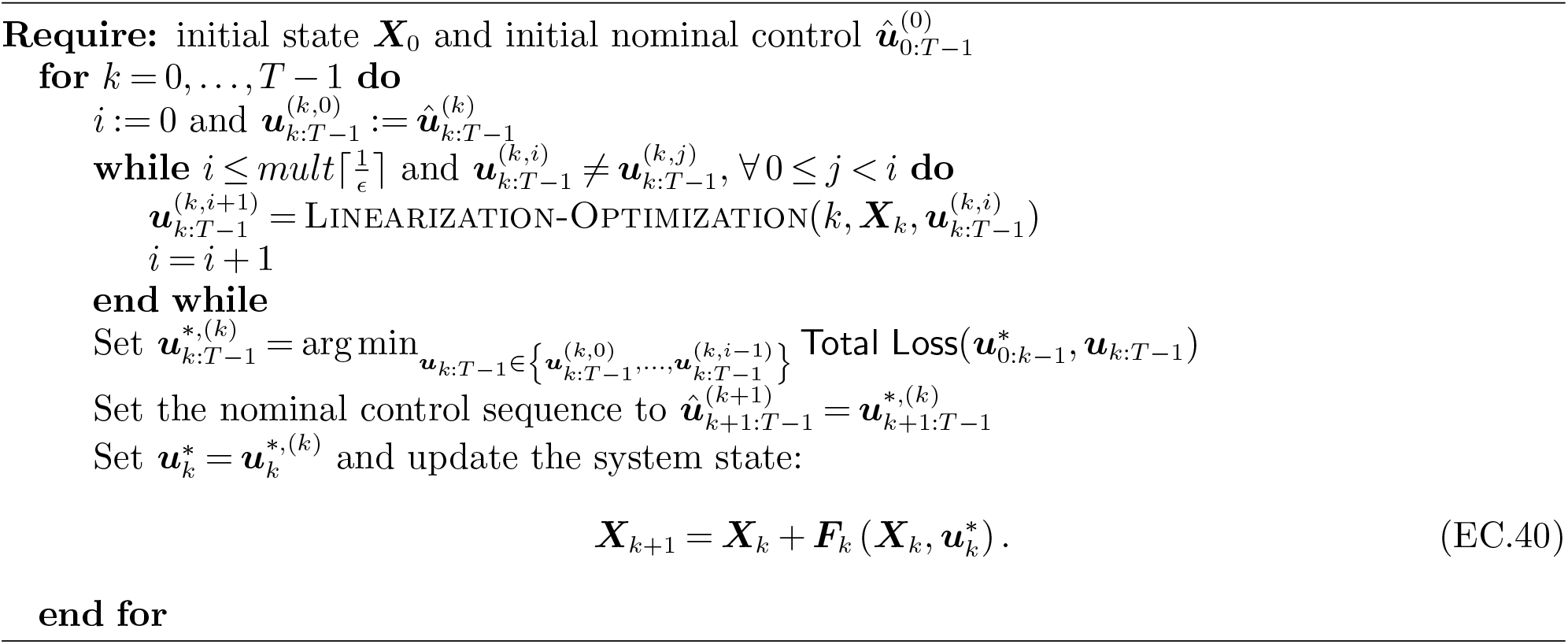

i. A gradient descent method that can vary activity levels across activities, age groups, and time is initialized at the ROLD AGE solution. Then ROLD is initialized at the solution of the gradient descent method.
ii. A gradient descent method that can vary activity levels across activities, age groups, and time is initialized at the ROLD ACT solution. Then ROLD is initialized at the solution of the gradient descent method.

### EC.4. Details on Parametrization and Calibration of the Model for Île-de-France

#### EC.4.1. Basic SEIR Model Parameters

The SEIR model parameters that are constant across age groups are summarized in Table EC.1. The age-group specific parameters are reported in Table EC.2. We start with the parameters as reported in Salje et al. (2020)12., and then we allow the values of parameters 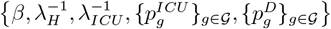 to change through time so as to model changes in how hospitals manage COVID-19 patients and changes in mandates for using masks and other measures that reduce transmission (details are presented in Section EC.4.2).

For *R*_0_ and *λ*_*H*_, the reported uncertainty ranges are 95% confidence intervals. For *σ*^−1^ (i.e., the mean stay in compartment *E*), the uncertainty range is calculated as 4 *±*0.8*·* 0.6 days, where 0.6 days is half the width of the 95% confidence interval for the incubation period reported in Bi et al. (2020), and 0.8 accounts for the fact that the stay in compartment *E* is 4*/*5 of the mean incubation time in Salje et al. (2020). For *μ*^−1^ (i.e., the mean stay in an infectious state), the uncertainty range is calculated as 4 *±*0.43, where 0.43 is half the width of the 95% confidence interval for the serial interval reported by Du et al. (2020).13. For the average stay in the ICU, we add to the mean stay of 20.46 days for Île-de-France, another 1.5 day, which is the mean time spent in hospital prior to ICU admission (Salje et al. 2020).

##### Calculating the transmission rate *β* from *R*_0_

We obtain *β* by linearizing the dynamics for *E*_*g*_, *I*_*g*_ around a point where *S*_*h*_ ≈ *N*_*h*_, *I*_*h*_ ≈ 0, ∀ *h*. More precisely, we have:

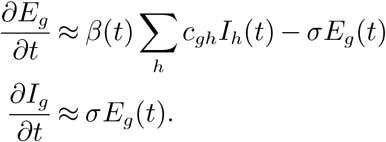

**Table EC.1.** SEIR model parameters

| Variable | Description | Value<br>[Uncertainty Range] | Reference |
| --- | --- | --- | --- |
| $R_0$ | Basic reproduction number | 2.9<br>[2.8 - 2.99] | Salje et al. (2020) |
| $\sigma^{-1}$ | Latency period | 4.0 days<br>[3.52 - 4.48] | Salje et al. (2020), Bi et al. (2020) |
| $\mu^{-1}$ | Infectious period | 4.0 days<br>[3.57 - 4.43] | Salje et al. (2020), Du et al. (2020) |
| $(\lambda^H)^{-1}$ | Average time spent in Hospital | 14.94 days<br>[14.55 - 15.32] | Salje et al. (2020) |
| $(\lambda^{ICU})^{-1}$ | Average time spent in ICU | 21.96 days<br>[21.38 - 22.6] | Salje et al. (2020) |

**Table EC.2.**
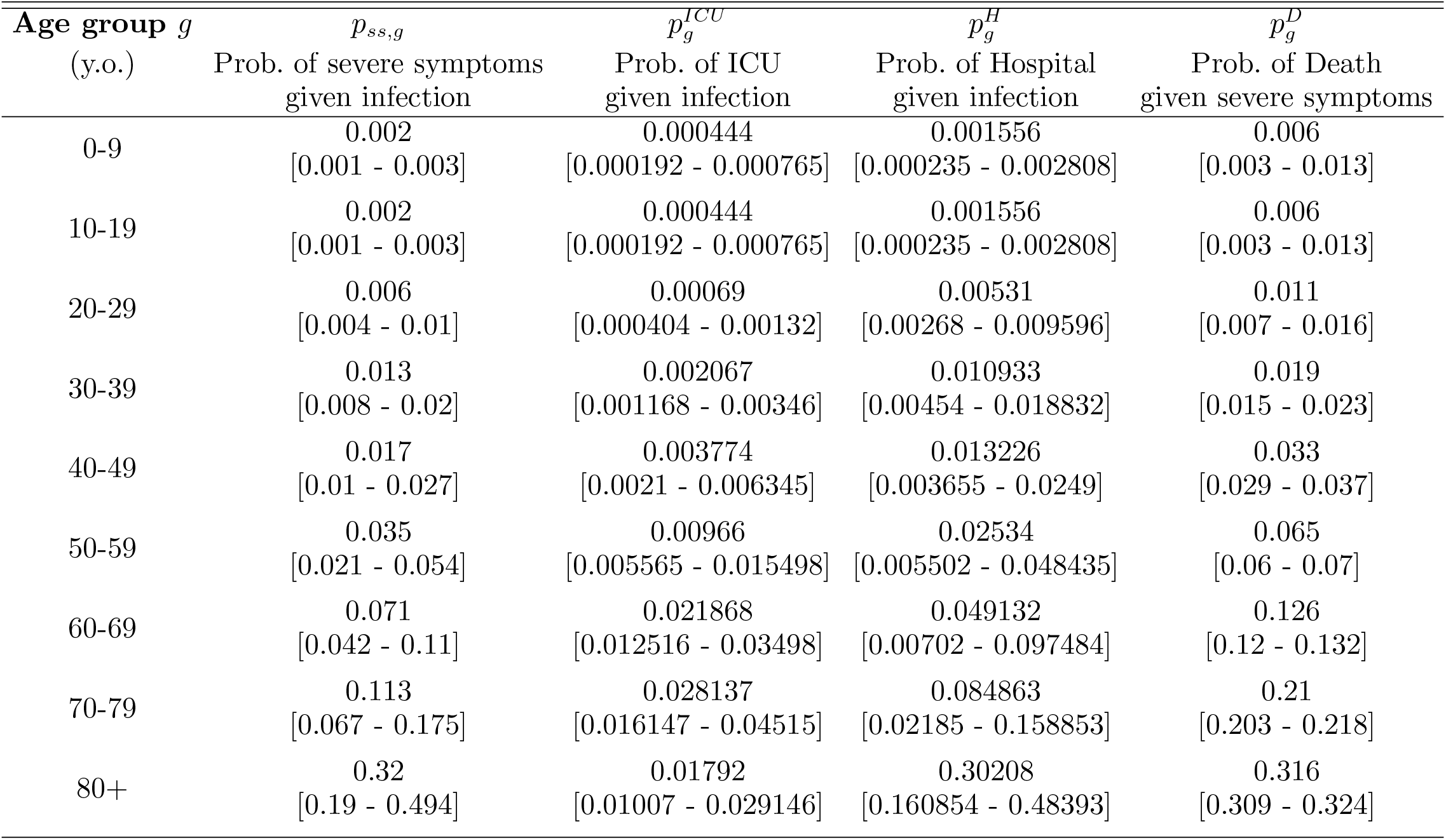
Age-group specific SEIR model probability parameters

Then, with ***Y*** (*t*) := [*E*_1_ (*t*), *E*_2_ (*t*), …, *E*_|*𝒢*|_, *I*_1_ (*t*), …, *I*_|*𝒢*|_]^*T*^, we can write 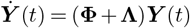, where

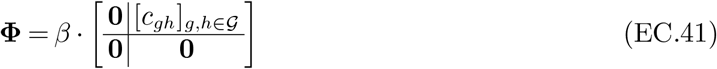

and

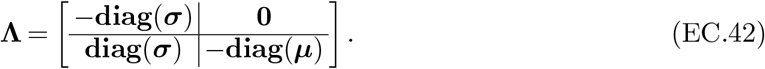

Then *R*_0_ can be identified as the spectral radius (i.e., the largest absolute value of the eigenvalues) of the matrix −**ΦΛ**^−1^ (Diekmann et al. 2010, Perasso 2018). Since the eigenvalues of a matrix *β ·* ***A*** are simply *β* multiples of the eigenvalues of ***A***, we can therefore determine *β* as *R*_0_ divided by the spectral radius of the matrix (−**Φ***/β*) **Λ**^−1^.

#### EC.4.2. Epidemiological Model Parameter Fitting Using Health Outcomes and Mobility Data

We use data on health outcomes from the French Public Health Agency (French Government 2020), as well as Google mobility data (Google 2020), to estimate the unknown parameters in our model. The data on health outcomes includes counts for individuals who are in the hospital, in the ICU, and who have died, by age group, and is maintained and updated daily by the French Public Health Agency (Santé publique France). The Google mobility data reports changes in activity at different places compared to a baseline, and is calculated using aggregate and anonymized data. For both health outcomes and mobility, we use data specific to the Île-de-France region.

The calibration exercise has two purposes: (a) to further refine the SEIR parameters reported in the literature to the data observed in Île-de-France and (b) to estimate our other parameters for which we do not have existing references. We then use the values of the estimated parameters in our experiments and simulations.

We first describe the set of parameters to be estimated, which we denote by 𝒫.

- Date of patient zero. We assume that the SEIR process starts with an infected individual of the 40-49 y.o. age group (Mohammad 2020). We wish to estimate the date when this infection occurs.
- Epidemiological parameters. We use the epidemiological parameters of Salje et al. (2020) to initialize the SEIR model. We allow these parameters to change in time in order to model changes in the way hospitals manage COVID-19 patients, as well as changes in mandates for using masks and other measures that reduce transmission. We assume that on date *d*, each parameter from the set 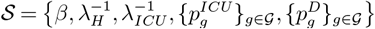 changes with respect to its initial value (as reported in Section EC.4.1), according to the relationship

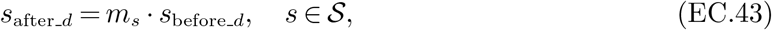

where *m*_*s*_ is a multiplier pertaining to parameter *s*. We assume the same multiplier 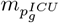 for all groups *g* ∈ *𝒢*, and similarly for 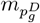. We seek to determine the date of change *d* as well as the multipliers *m*_*s*_, *s* ∈ *𝒮*.
- Confinement patterns. To estimate activity levels for the activities in our social mixing model, we use Google mobility data (Google 2020). The mobility data reports changes of activity (visits and length of stay) for each day, compared to a baseline value. The baseline used corresponds to the median value for the corresponding day of the week, during the five-week period January 3 - February 6 2020. We fix the home activity level to be equal to 1, throughout time. We estimate the level of the other activities using the corresponding activities from the Google mobility data, as shown in Table EC.3.

**Table EC.3.** Mapping between the activities in our model and the activities in the Google mobility data What remains to be estimated in the calibration is the weight parameter *α*_other_, as well as the school activity levels. We calibrate the level of schooling activity for four different time periods until October 21 2020. These periods are chosen to reflect (i) the dates when the French government closed down schools, and (ii) the French school calendar and summer recess.
- Social mixing parameter. To reduce the number of parameters to be calibrated, we simplify our mixing model in (1) by constraining *α*_1_ = *α*_2_. We seek to determine this mixing parameter.

| Activity in Our Model | Activity in Google Mobility Data |
| --- | --- |
| work | Workplaces |
| transport | Transit stations |
| leisure | Retail & recreation |
| other | $\alpha_{\text{other}} \cdot \text{Retail \& recreation} + (1 - \alpha_{\text{other}}) \cdot \text{Grocery \& pharmacy}$ |

We next describe the details of the fitting procedure that we set up in order to retrieve an optimal parameter fitting. The mixing dynamics of the SEIR model are driven by the vector of activity levels *ℓ*_*g*_ of each age group (as described in Section 3.2). Data on activity levels can be noisy; we model this uncertainty by assuming the vector of activity levels is a random vector, distributed as follows:

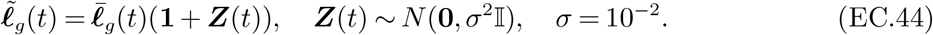

The value 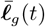 is obtained from the Google activity data at time *t*; this dataset does not differentiate activity by age, so 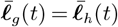 for all *g, h* ∈ *𝒢*; in other words, all groups are assigned the same activity level. Recall that *H*_*g*_(*t*), *ICU*_*g*_(*t*), *D*_*g*_(*t*) denote the hospital utilization, ICU utilization and cumulative number of deaths according to the SEIR model, respectively. We denote these quantities with 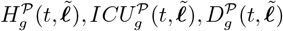 to emphasize the dependence of the SEIR process on parameters 𝒫 and the vector of activity levels 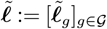. We aggregate these quantities over all age groups:

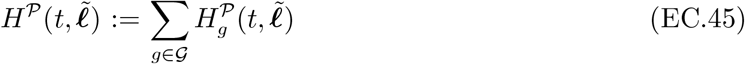

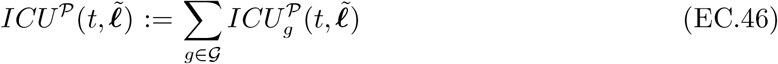

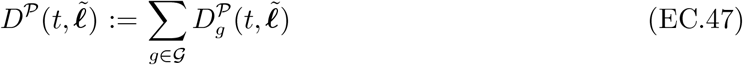

Denote by *H*^*obs*^(*t*), *ICU*^*obs*^(*t*), *D*^*obs*^(*t*) the general ward hospital beds utilization, ICU beds utilization, and cumulative deaths, respectively, at time *t*, as observed in the real data for Île-de-France from the French Public Health Agency (French Government 2020). We calculate the relative fitting error of the SEIR model at time *t* for each of these three quantities as

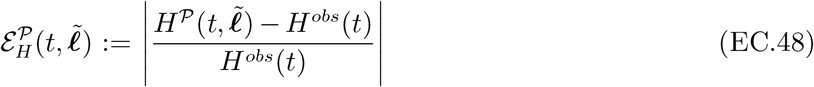

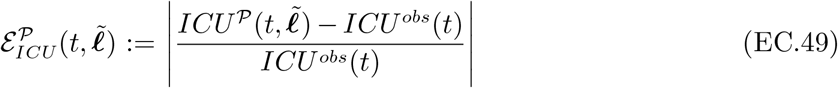

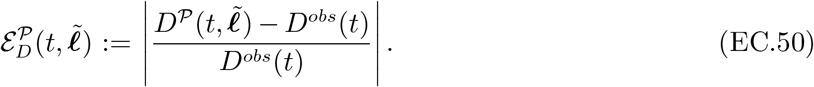

We define the total expected fitting error as a sum of these errors over different time intervals:

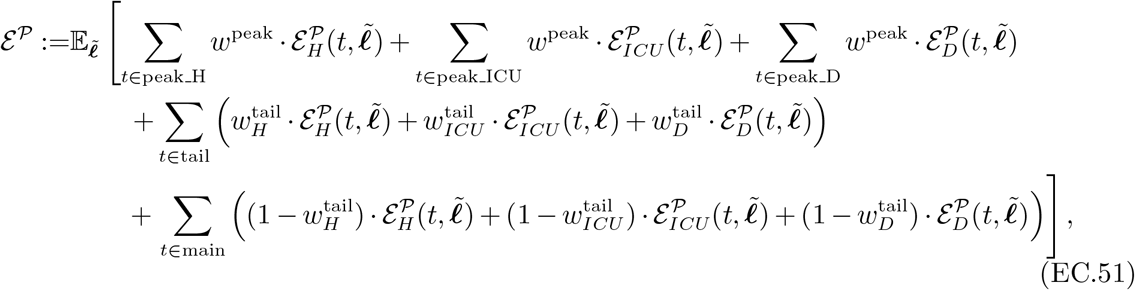

where the expectation is taken with respect to random vector 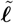, and where the time intervals are defined in Table EC.4 and comprise the entire period between March 17 2020 and October 21 2020. Our approach penalizes the errors at the peak times for hospital beds utilization, ICU beds utilization, and deaths; it also penalizes errors over the last 14 days of the considered period, to ensure an accurate fit at the end of the calibration horizon. We use different weights to account for the different errors. We use *w*^peak^ = 1*/*6 to account for the relatively smaller period of the peaks. We use 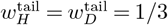 and 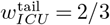, with a higher weight for ICU beds utilization so as to ensure low error in the tail predictions of ICU utilization, as ICU beds utilization towards the tail of the calibration window plays an important role in the dynamics of the model right after the calibration window.

**Table EC.4.**
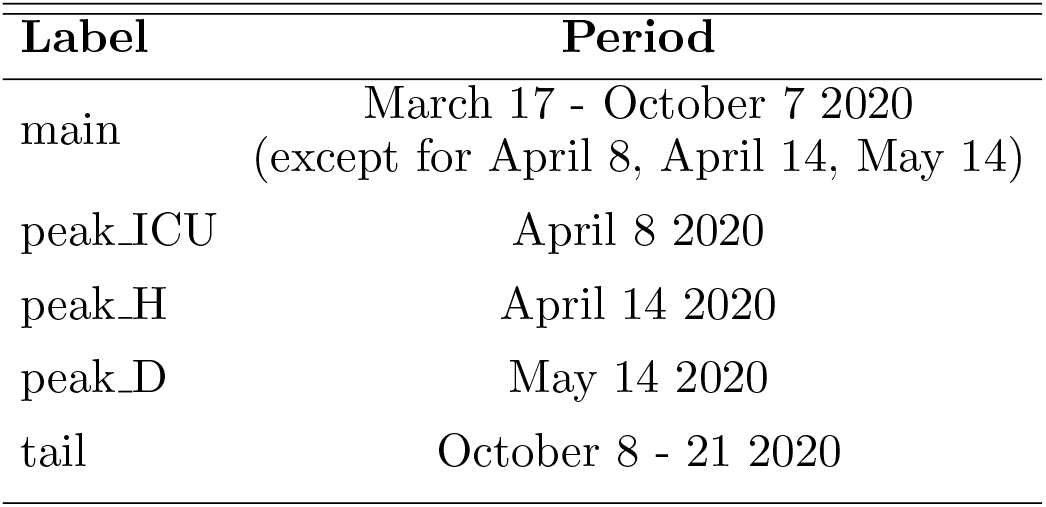
Time intervals used in the calibration

We seek to determine the set of parameters 𝒫 that minimize ℰ^𝒫^ in (EC.51). We approximate the expectation in (EC.51) through a Monte Carlo sample-average approximation, using 100 samples. The set 𝒫 contains both discrete and continuous parameters. To minimize ℰ^𝒫^, we first do a grid search over all possible combinations for the discrete parameters, and then for each such combination, we perform a gradient descent procedure over the space of the continuous parameters. Each parameter is optimized within an allowed range, which is informed from the context of Île-de-France in 2020. We used a wide allowed range if we had no information on what could be reasonable values for a given parameter.

Our calibration procedure yields the parameter fitting summarized in Table EC.5. We compare the fitted values of the SEIR model with the values reported by the French Public Health Agency in Figure 2. Figure EC.1 presents the results from the out-of-sample model validation analysis.

#### EC.4.3. Economic Model Parameter Fitting

We obtain data on population, employment, and wages from the French National Institute of Statistics and Economic Studies (Institut national de la statistique et des études économiques — INSEE). Where relevant, we discount all cash flows at a 3% annualized rate. We set the retirement age to be 65 (i.e., 64 is the last working year of age.) We first obtain the initial population data *N*_*g*_(0) for each age group in Île-de-France at the end of 2019 from INSEE (2020).

##### Estimation of *w*_*g*_

Recall that *w*_*g*_ in (EC.18) corresponds to the employment value for a member of group *g*, under normal conditions. To estimate *w*_*g*_, we use two datasets from INSEE:

- Yearly full time equivalent (FTE)14. wages and employed population count for Île-de-France in 2016, broken up into the age groups “under 26 years old”, “26 to 49 years old” and “more than 50 years old” (INSEE 2016b).
- FTE employment rates across the entire economy for the fourth quarter of 2019, bucketed by age groups “15 to 24 years old”, “25 to 49 years old”, “50 to 64 years old”, and “55 to 64 years old” (INSEE 2019).

**Table EC.5.** Calibration: fitted epidemiological and economic parameters and allowed estimation ranges

| Parameter | Estimated Value | Allowed range |
| --- | --- | --- |
| Date of patient zero | Jan 1 2020 | Dec 8 2019 - Jan 27 2020 |
| Date $d$ when epidemiological parameters change | Apr 16 2020 | Mar 1 2020 - Apr 30 2020 |
| Multiplier $m_\beta$ | 0.60 | 0.50-2.00 |
| Multiplier $m_{p_g^{ICU}}$ | 1.66 | 0.50-2.00 |
| Multiplier $m_{p_g^D}$ | 1.13 | 0.50-2.00 |
| Multiplier $m_{\lambda_H}$ | 1.50 | 0.50-2.00 |
| Multiplier $m_{\lambda_{ICU}}$ | 2.00 | 0.50-2.00 |
| Weight for activities $\alpha_{\text{other}}$ | 0.96 | 0.00-1.00 |
| School activity $\ell^{\text{school}}$ Mar 17 - May 10 2020 | 0.00 | 0.00-0.10 |
| School activity $\ell^{\text{school}}$ May 11 - Jun 30 2020 | 0.00 | 0.00-1.00 |
| School activity $\ell^{\text{school}}$ Jul 1 - Aug 31 2020 | 0.20 | 0.00-0.20 |
| School activity $\ell^{\text{school}}$ Sep 1 - Oct 21 2020 | 1.00 | 0.50-1.00 |
| Social mixing $\alpha$ | 0.39 | 0.10-2.00 |
| Sensitivity of econ. value $\nu^{\text{work}}$ | 0.50 | |
| Sensitivity of econ. value $\nu^{\text{fixed}}$ | 0.47 | |

Since we do not have a consolidated data source for economic data split by our exact age group definitions, we use the above datasets to interpolate values for *w*_*g*_. At a high level, we derive wage curves across age ranges.

We next explain the general procedure, as well as the additional assumptions we have made for the interpolation. First, for the construction of wage curve by age bucket:

- We assume that the national level employment rates from INSEE (2019) are equal to those of the Île-de-France region. Because the age bucketing for our age groups is finer than the age bucketing in the data, we use interpolation. Specifically, we fit a piece-wise linear model (consisting of three pieces) to the four employment rates reported for the “15 to 24 years old”, “25 to 49 years old”, “50 to 64 years old”, and “55 to 64 years old” groups. We take the midpoint of the age group as the *x* value of the datapoint; for example, for “50 to 64 years old” we use a midpoint of 57.5. With this model, we can infer an employment rate for any arbitrary age and construct an employment rate curve.
- We perform a similar fitting procedure for the age group wage information from INSEE (2016b); since the wage progression by age is much smoother, we use simple linear regression to construct a wage curve for each one of our age buckets.
- The previous wage curve only accounts for the employed population, whereas our age groups count the entire population. We thus combine the wage curve with employment rate and population data to arrive at a wage number blended across an entire age group’s population.

When doing this, we treat the 10-19 y.o. and 60-69 y.o. age groups specially by assuming the employment rates are reported only with respect to the work-eligible population in that bucket (15-19 and 60-64 year olds, respectively). We also set the work-eligible population for the 0-9, 70-79, and 80+ age buckets to 0. The formula we use is

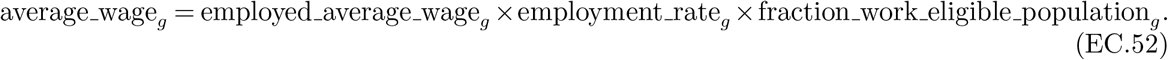

**Figure EC.1.**
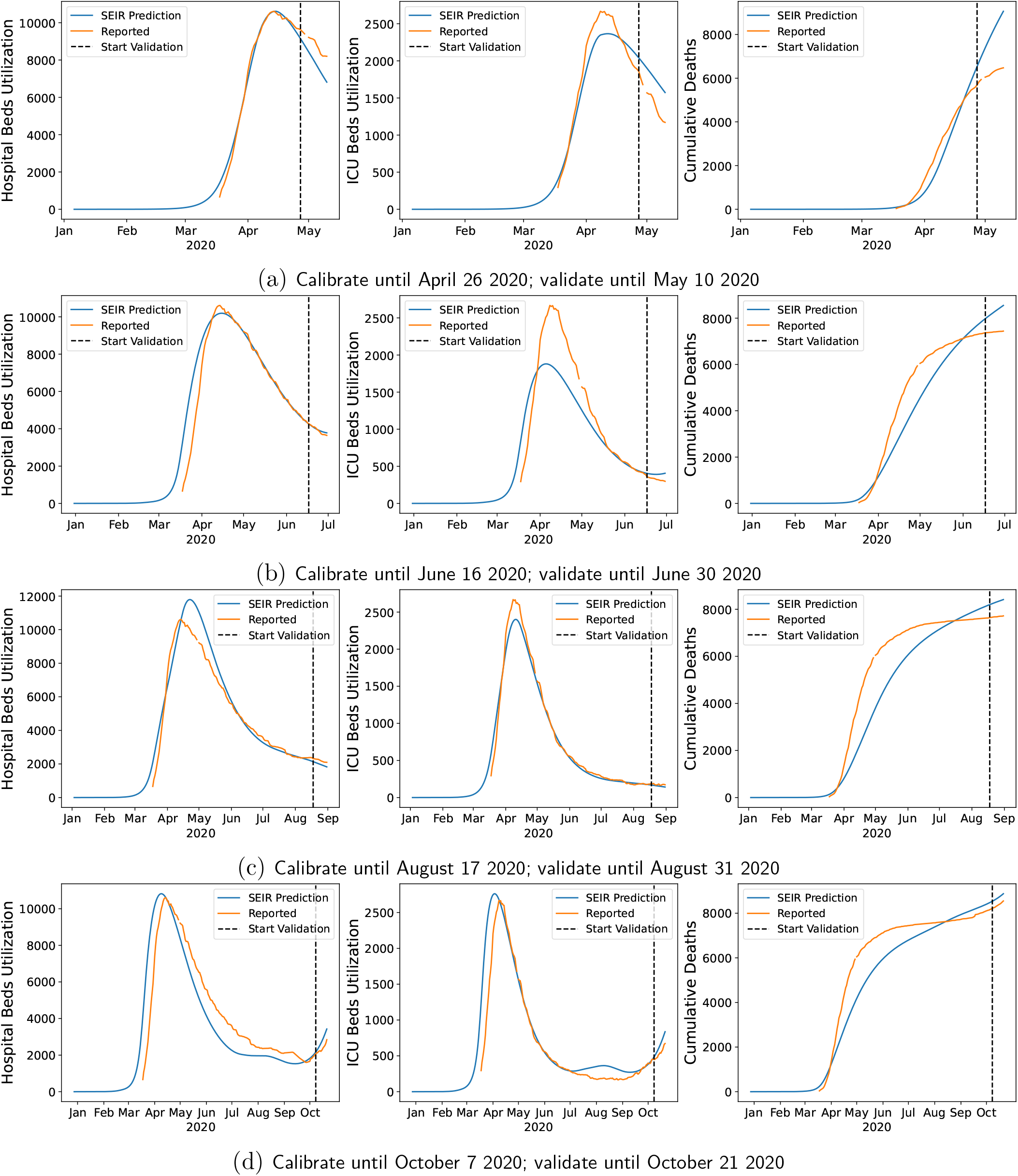
Out-of-sample predictions of the fitted SEIR model vs. reported values by the French Public Health Agency for hospital beds utilization (left), ICU beds utilization (middle) and cumulative deaths (right). For each of four dates *t* (**(a)**: May 10 2020; **(b)**: June 30 2020; **(c)**: August 31 2020; **(d): October 21 2020**) we calibrate the SEIR model using data up until day *t* −14; and then run the SEIR model up to day *t*. The dashed line indicates the start of the out-of-sample validation (i.e., *t* − 13).

- The interpolations we use introduce errors: in particular, if we aggregate the wages inferred by our constructed curve across the entire population, we overestimate the real total wages by 5.12%. We scale all wages average wage_*g*_ proportionally so as to retrieve the real total wage amount *w*_*g*_.

Table EC.6 summarizes the year-based employment contribution parameters per age group. We note that when using them in the objective of the optimization problem, we divide these year-based values by 365, in order to capture employment value on a daily basis.

**Table EC.6.** Year-based economic value parameters under normal activity, per age group (in €)

| Age Group $g$ (y.o.) | $w_g \times 365$ (€) |
| --- | --- |
| 0-9 | 0.00 |
| 10-19 | 2084.79 |
| 20-29 | 11 743.20 |
| 30-39 | 25 799.29 |
| 40-49 | 31 746.68 |
| 50-59 | 32 573.79 |
| 60-69 | 12 640.83 |
| 70-79 | 0.00 |
| 80+ | 0.00 |

##### Estimation of *ν*^work^, *ν*^other activities^, *ν*^fixed^

We move on to the estimation of parameters *ν*^work^, *ν*^other activities^, *ν*^fixed^ in (EC.18). These measure the sensitivity of economic value to the confinement pattern (*t*). We estimate them from data on lost economic output during the first lockdown phase employed in Île-de-France, and in particular using the month of April 2020. We break up the approach into a few steps:

- We use survey data of French managers regarding business activity during the lockdown starting March 17 2020 from the Bank of France. This is sentiment data where managers are asked to compare current business conditions to normal conditions for the same relevant time period (Banque de France 2020a,b). These data are reported by industry, and we aggregate them into a single number weighting by industry size. We use FTE wages and employed population count for the Île-de-France region in 2016 (INSEE 2016a) to figure out the appropriate weights to use in the aggregation. We then use these monthly readings as proxies for the economic activity level due to confinements in the month of April 2020, as compared to normal activity. The economic activity level for the month of April is 58.51%.
- A requirement for our estimation are the precise levels of confinement in April 2020. We retrieve these from Google mobility data (Google 2020), as explained in Section EC.4.2. To simplify the estimation, we set *ν*^other activities^ = 0 and then determine parameters *ν*^work^, *ν*^fixed^ solving the system of equations

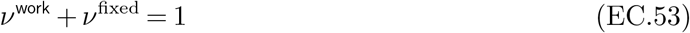

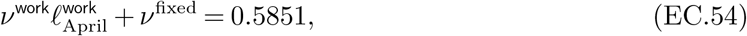

where 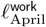 corresponds to the average value of *ℓ*^work^(*t*) through the month of April 2020. In our experiments, we also test our algorithm in alternative scenarios where we set *ν*^other activities^ *>* 0, keep the value for *ν*^fixed^ from the system (EC.53)-(EC.54), and adjust *ν*^work^ = 1 − *ν*^other activities^ − *ν*^fixed^. The specific values we test are *ν*^other activities^ ∈ {0, 0.1, 0.2}.

### EC.5. Benchmark Policies

We compare ROLD to several simpler classes of policies drawing inspiration by real life confinement management rules:

#### ICU admissions trigger policy — ICU-t

This class of policies is similar in spirit to the trigger rule proposed by Duque et al. (2020) for the Austin metropolitan area. This rule places all age groups and activities (except home) at a strict level of confinement when the average seven-day hospital admissions (i.e., inflow of patients admitted into the hospital due to COVID-19 complications) exceeds a pre-determined threshold, and then changes the confinement to a relaxed level when the average seven-day hospital admissions and the hospital utilization rate drop below pre-determined thresholds.

Since Duque et al. (2020) does not differentiate between hospital and ICU beds, we define our policy class on ICU admissions and utilization instead of hospitalizations. Specifically, the ICU admissions trigger policy is defined as in Algorithm 5.

##### Algorithm 5

ICU Admissions Trigger Policy — ICU-t

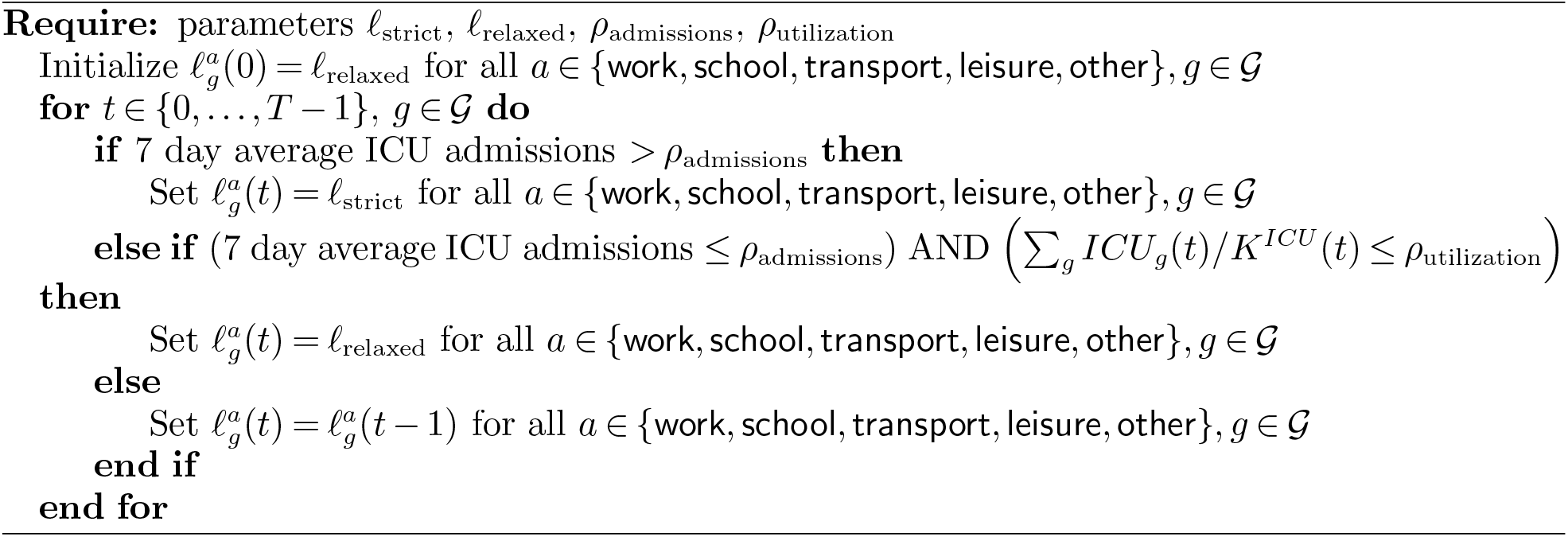

We optimize over the parameters *ℓ*_strict_, *ℓ*_relaxed_, *ρ*_admissions_ and *ρ*_utilization_ via grid search with the goal of minimizing the objective in (8) corresponding to the total economic and death loss due to the pandemic, and we report the performance of the best policy.

#### Hybrid Trigger Policy — Hybrid-t

This policy resembles the rule used in France for declaring a region in “maximum alert”15. (Lehot and Borgne 2020). Like the previous policy, this policy also switches between a (uniform) strict and a more relaxed confinement level, but the trigger condition combines ICU utilization with the rate of new infections in the population. Specifically, the policy switches to strict confinement if the average seven-day incidence rate in the population, defined16. as ∑_*t*−6≤ *τ* ≤*t*_ ∑_*g*_ new infections_*g*_(*τ*)*/* ∑_*g*_ *N*_*g*_(0), is greater than a threshold *ρ*_incidence_, *and* the incidence rate in age groups corresponding to the population that is 60 y.o. and above, ∑_*t*−6≤*τ* ≤*t*_ ∑_*g*≤60 y.o._ new infections_*g*_(*τ*)*/* ∑_*g*≤60 y.o._ *N*_*g*_(0), is greater than a threshold *ρ*_incidence 60+_, *and* the ICU utilization rate is greater than a threshold *ρ*_utilization_. We optimize over all parameters *ℓ*_strict_, *ℓ*_relaxed_, *ρ*_incidence_, *ρ*_incidence 60+_ and *ρ*_utilization_ with the goal of minimizing the total loss objective in (8), and we report the performance of the best policy.

This is the Hybrid-t AND policy, and it is described in Algorithm 6. We also test a stricter version of this policy that takes the logical *or* of the three conditions (Hybrid-t OR), instead of taking the *and*, as the trigger for setting the strict confinement level.

#### Fully open — FO

This corresponds to the normal conditions where 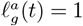 for all *a* ∈ {home, work, school, transport, leisure, other }, *g* ∈ 𝒢 and *t* ∈ {0, …, *T* − 1}.

#### Full confinement — FC

In this policy, all activities except home are fully restricted. That is, we set 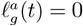 for all *a* ∈ {work, school, transport, leisure, other}, *g* ∈ 𝒢, *t* ∈ {0, …, *T* − 1}, and 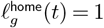 for all *g* ∈ 𝒢 and *t* ∈ {0, …, *T* − 1}.

##### Algorithm 6

Hybrid Trigger Policy — Hybrid-t AND

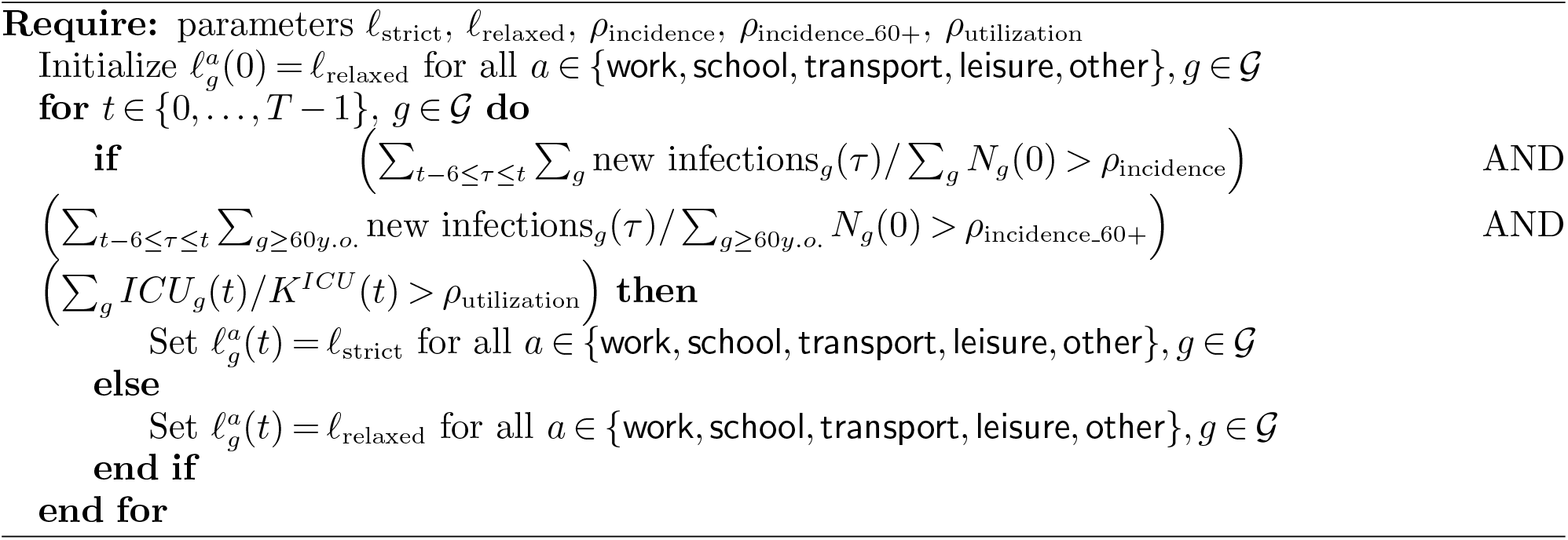

### EC.6. Additional Results

#### EC.6.1. Robustness Analyses

We analyze additional problem instances by changing the value of each of 13 estimated parameters within a sensitivity range, as shown in Table EC.7. For each parameter, we sample 40 values uniformly at random from a specified sensitivity range. In each problem instance, one parameter is changed from its estimated value, for a total of 13 *×* 40 = 520 problem instances.

**Table EC.7.** Robustness analysis: parameters and sensitivity analysis ranges

| Parameter | Estimated Value | Sensitivity Range |
| --- | --- | --- |
| Multiplier $m_\beta$ | 0.60 | 0.45-0.75 |
| Multiplier $m_{p_g^{ICU}}$ | 1.66 | 1.51-1.81 |
| Multiplier $m_{p_g^D}$ | 1.13 | 0.98-1.28 |
| Multiplier $m_{\lambda_H}$ | 1.50 | 1.35-1.65 |
| Multiplier $m_{\lambda_{ICU}}$ | 2.00 | 1.85-2.15 |
| Weight for activities $\alpha_{\text{other}}$ | 0.96 | 0.76-1.00 |
| School activity $\ell^{\text{school}}$ Mar 17 - May 10 2020 | 0.00 | 0.00-0.20 |
| School activity $\ell^{\text{school}}$ May 11 - Jun 30 2020 | 0.00 | 0.00-0.20 |
| School activity $\ell^{\text{school}}$ Jul 1 - Aug 31 2020 | 0.20 | 0.00-0.40 |
| School activity $\ell^{\text{school}}$ Sep 1 - Oct 21 2020 | 1.00 | 0.80-1.00 |
| Social mixing $\alpha$ | 0.39 | 0.19-0.59 |
| Work activity $\ell_{\text{April}}^{\text{work}}$ April 2020 | 0.213 | 0.113-0.313 |
| Economic activity level April 2020 | 58.51% | 48.51%-68.51% |

Figure EC.2 shows robustness results for seven values of the economic cost of death *χ*: [0, 10, 15, 25, 50, 100, 150] *×* the annual GDP per capita in France. The shown boxplots summarize results over the 520 problem instances, for each value of *χ*. These results reinforce our findings from Section 6 on the gains of dual targeting, as well as the observation that dual targeting unlocks complementarities which may not be available under targeting age groups or activities separately.

**Figure EC.2.**
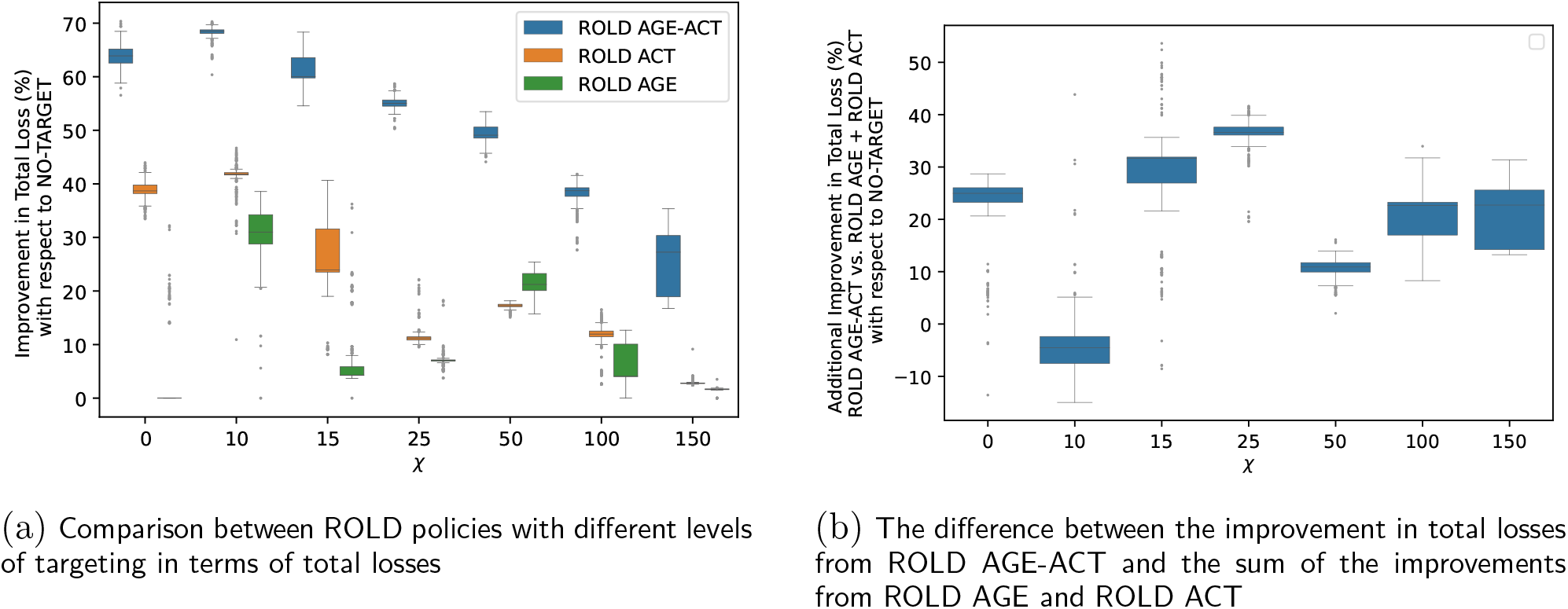
Robustness analyses showing the superiority of ROLD AGE-ACT over ROLD policies with less granular targeting, and the super-additive improvements of ROLD AGE-ACT over the sum of the improvements of ROLD AGE and ROLD ACT, for different values for the cost of death *χ*, over a wide set of problem instances. All improvements are with respect to ROLD NO-TARGET. For each value of *χ*, the boxplots summarize results over 520 different problem instances.

#### EC.6.2. Can Dual Targeting Increase Average Activity Levels for Each Age Group?

For each ROLD policy, we calculate the time-average activity level of each age group, averaged over the activities relevant to that age group (listed in Table EC.8), and weighting different activities by the respective participation rates. We split the 24-hour day in 48 slots of 30 minutes each, indexed *τ* = 1, …, 48, and let 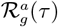 denote the participation rate of age group *g* in activity *a* during time slot *τ* of a generic day (so that 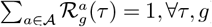). We retrieve the participation rates from time-use survey data we obtained from the French National Archive of Data from Official Statistics (ADISP, INSEE 2010). We detail our procedure for estimating participation rates in Appendix EC.8.1. Then, for group 0-9 y.o., the time-average activity level is defined as

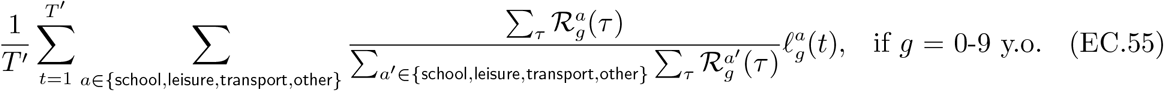

and similarly for other age groups.

#### EC.6.3. Additional Details for How Gains Arise from Dual Targeting

Figure EC.3 shows the activity levels of different (age group, activity) pairs, with pairs divided into three buckets (high, medium, low) in terms of their econ-to-contacts-ratio. The ROLD policy tends to enforce stricter confinements in earlier periods and subsequently relaxes these through time.

Figure EC.4 visualizes the optimized confinement policy for the value *χ* = 60*×*, which is in the mid-range of estimates used in the economics literature on COVID-19 (Alvarez et al. 2020) and is representative of the overall behavior we observe across all experiments. Groups 20-69 y.o. remain more open in work but face confinement in leisure for up to the first ten weeks; on the contrary, the 10-19 y.o. group is confined in work for a long period while remaining open in leisure, and the 70+ y.o. groups also remain open in leisure.

**Table EC.8.** Relevant activities for each age group, excluding home

| Age Group $g$ (y.o.) | Relevant Activities |
| --- | --- |
| 0-9 | school, leisure, transport, other |
| 10-19 | school, work, leisure, transport, other |
| 20-29 | work, leisure, transport, other |
| 30-39 | work, leisure, transport, other |
| 40-49 | work, leisure, transport, other |
| 50-59 | work, leisure, transport, other |
| 60-69 | work, leisure, transport, other |
| 70-79 | leisure, transport, other |
| 80+ | leisure, transport, other |

**Figure EC.3.**
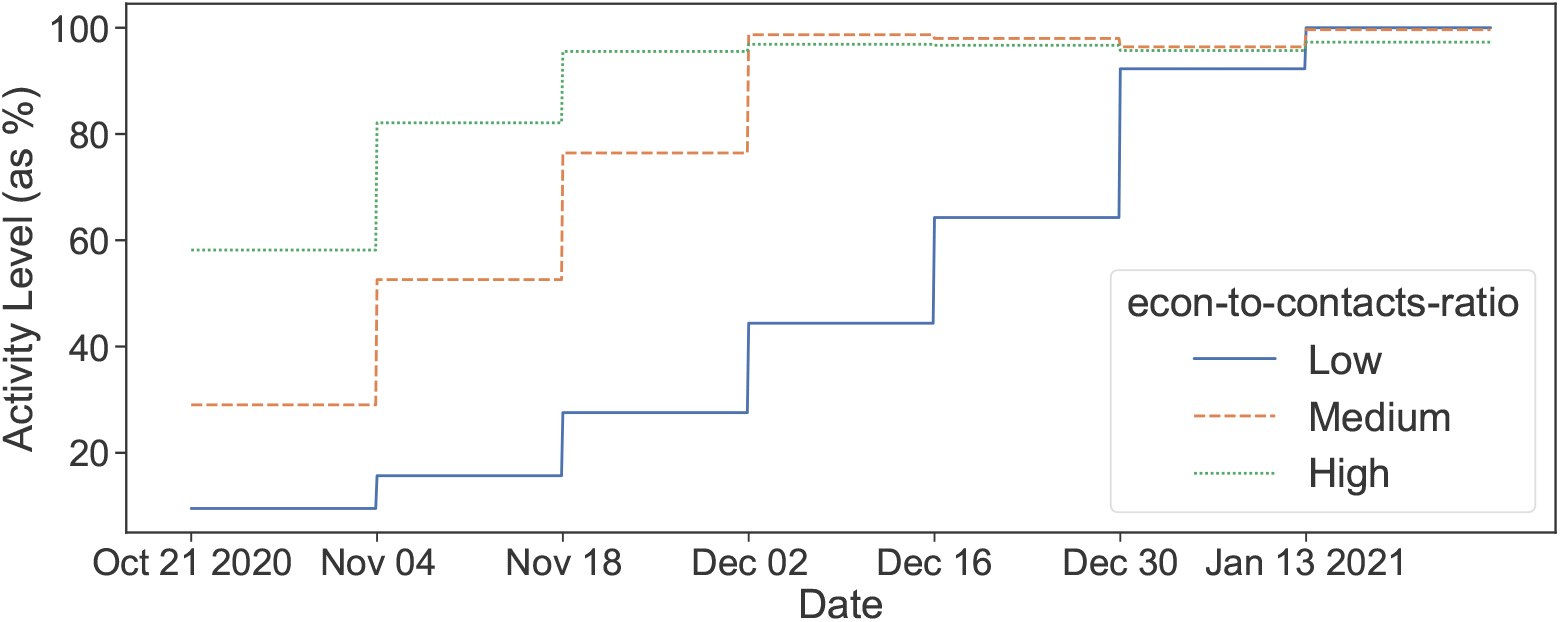
The activity levels of optimized ROLD AGE-ACT policies for different (age group, activity) pairs, bucketed in three equally-sized buckets according to their econ-to-contacts-ratio. The lines indicate the mean for each bucket, where for each time *t* the mean is taken over the different values of the cost of death *χ* and over the relevant (age group, activity) pairs.

#### EC.6.4. Additional Details for the Regression Decision Trees

To understand how the ROLD policies target confinements across different age groups and activities, we train an interpretable machine learning model – a regression decision tree – to predict the optimal ROLD confinement in each activity as a function of several features.

To build a training set, we first create a larger set of problem instances built using a wide range of problem parameters. We vary the following parameters: basic reproduction number *R*_0_, the scaling of the group-specific probabilities *p*_*ss,g*_ of having severe symptoms conditioned on being in state *I*, the social mixing parameter *α*, the cost of death *χ*, the ICU capacity *K*^ICU^, the economic model parameter *ν*^fixed^, the ratio of economic model parameters *ν*^other activities^*/ν*^work^, and the activity weights *w*^leisure^, *w*^transport^, *w*^other^ used in the model for 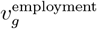 in (EC.18). We create each instance by drawing each of these parameters independently and uniformly at random from a range that is specified in Table EC.9.

All resulting instances use an optimization horizon of *T*′ = 90 days and change the confinement decisions every 14 days. For each problem instance, we compute the optimal ROLD AGE-ACT, ACT, and NO-TARGET activity levels, i.e., the decisions 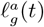 for *g* ∈ *𝒢, a* ∈ *𝒜*, 0 ≤ *t* ≤ *T* − 1, which we then use to simulate the SEIR dynamics and calculate all the corresponding values of the SEIR states.

**Figure EC.4.**
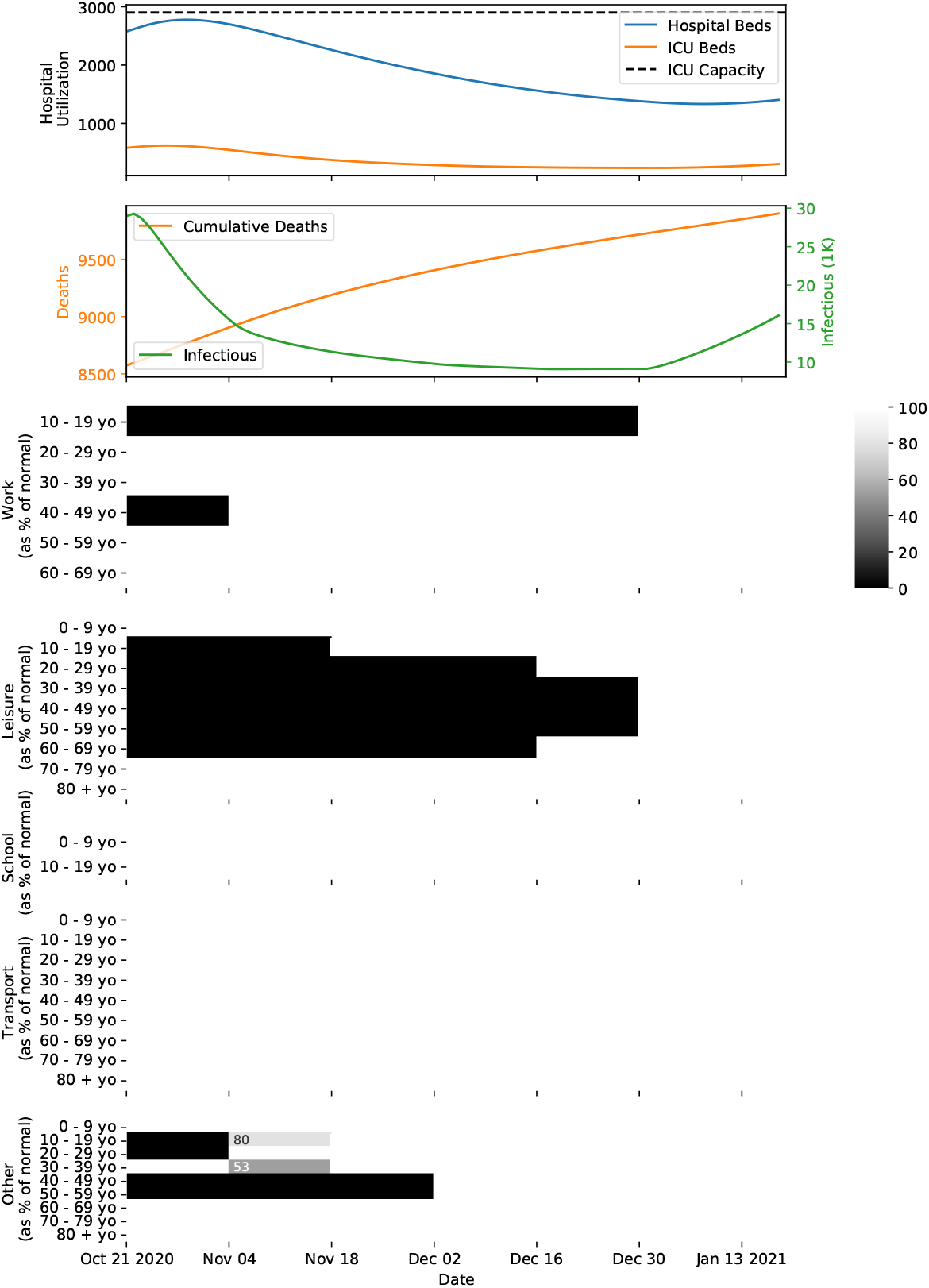
The optimized ROLD AGE-ACT policy for a problem instance with a 90-day optimization horizon starting on October 21, 2020, with 2900 ICU beds, and with cost of death *χ* = 60*×*. From top to bottom, the seven panels depict the time evolution for the occupation of hospital and ICU beds, the number of infectious individuals, the cumulative number of deaths in the population, and the confinement policy imposed by ROLD in each age group and activity. In panels 3-7, the values correspond to the activity levels allowed and are color-coded so that darker shades capture a stricter confinement.

##### EC.6.4.1. Additional Details for the Regression Decision Trees for the ROLD AGE-ACT Policy

We create a training set of 5,524,785 samples — with one sample for every 14-day period (i.e., for *t* = 0, 14, 28, etc.), each age group *g*, and each activity *a* relevant to that age group — where we include several direct and derived features based on the parameter values characterizing the instance and the induced SEIR states, and a target corresponding to the optimal ROLD AGE-ACT decision 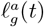. We consider both targeted features, by which we mean features that differentiate on either group *g* or activity *a*,17. or on both, and non-targeted features (Tables EC.10, EC.11). Some of the non-targeted features are allowed to depend on *t*.

Using this data as training set, we employ the CART algorithm (implemented in the scikitlearn Python library) to train regression decision trees to predict the optimal ROLD AGE-ACT confinement decisions across all activities and age groups as a function of the considered features, using the traditional mean-squared-error (MSE) criterion as a goodness of fit metric. When using *k*-fold cross-validation to assess the optimal depth of the tree, we noticed that the optimal depth exceeded 10. As such, we decided to train a tree of depth four for interpretability purposes, and we also trained trees with maximum depth ranging from four to ten to test the robustness of our results.

**Table EC.9.** Parameter variation used for fitting trees. Each problem instance is defined by drawing each parameter independently from the uniform distribution over the indicated range for that parameter.

| Parameter | Description | Min | Max | Notes |
| --- | --- | --- | --- | --- |
| $R_0$ | Basic reproduction number | 1 | 8 | |
| severity-scaling | Scales the probability of $p_{ss,g}$ for all groups $g$ | 0.1 | 3.125 | Max value ensures $p_{ss,80+} = 1$ |
| $\alpha$ | Social mixing parameter | 0.16 | 1.16 | Min/max values correspond resp. to contacts dropping to 80% and 20% of their level |
| $\chi$ | Cost of death | 0 | 400 | In multiples of GDP/capita |
| $K^{\text{ICU}}$ | ICU capacity | 592 | 4175 | Min(max) corresponds to the ICU filling completely in 10 (60) days from day 0 of the optimization horizon, assuming a fully open policy |
| $\nu^{\text{fixed}}$ | Economic model parameter | 0.1 | 0.9 | |
| $\frac{\nu^{\text{other activities}}}{\nu^{\text{work}}}$ | Ratio of economic model parameters | 0.01 | 0.36 | The estimated ratio of 3/50 was scaled up and down by a factor of 6 |
| $w^{\text{leisure}}, w^{\text{transport}}, w^{\text{other}}$ | Weights for econ. model for $v_g^{\text{employment}}$ in (EC.18) | - | - | Sample uniformly at random from the simplex defined by $w^{\text{leisure}} + w^{\text{transport}} + w^{\text{other}} = 1$; $w^{\text{leisure}}, w^{\text{transport}}, w^{\text{other}} \geq 0$ |

**Table EC.10.** Targeted features used for fitting trees for ROLD AGE-ACT

| Feature Name | Description |
| --- | --- |
| $g, a$ | Age group, activity |
| $p_{ss,g}$ | Probability of having severe symptoms conditional on infection for group $g$ |
| $p_g^D$ | Probability of dying conditional on severe symptoms for group $g$ |
| $v_g^{\text{life}}$ | Lost wages for group $g$ |
| $N_g(t)$ | $S_g(t) + E_g(t) + I_g(t) + R_g(t)$ |
| $\text{econ-gradient}(g, a, t)$ | The gradient of the economic value with respect to the activity level $\ell_g^a(t)$ normalized by $N_g(t)$, i.e., $\frac{\partial \text{Economic Value}(t) / \partial \ell_g^a(t)}{N_g(t)}$ |
| $\text{contacts-given}(g, a)$ | The number of social contacts given to other groups by group $g$ in activity $a$, i.e., $\sum_{h \in \mathcal{G}} C_{h,g}^a$ |
| $\text{econ-to-contacts-ratio}(g, a, t)$ | The ratio of $\text{econ-gradient}(g, a, t)$ and $\text{contacts-given}(g, a)$ |

**Table EC.11.** Non-targeted features used for fitting trees for ROLD AGE-ACT

exceeded 10. As such, we decided to train a tree of depth four for interpretability purposes, and **Table EC.11** Non-targeted features used for fitting trees for ROLD AGE-ACT
| Feature Name | Description |
| --- | --- |
| $t$ | Time period |
| $R_0$ | Basic reproduction number |
| contacts-elasticity | The percentage drop in total contacts when activity levels drop by 50%. Equal to $1 - 4^{-\alpha}$, where $\alpha$ is the social mixing parameter. |
| $\chi$ | Cost of death |
| $K^{\text{ICU}}$ | ICU capacity |
| R-perc($t$) | The percentage of the population estimated by the model to be in the recovered state at time $t$, i.e., $\sum_{g \in \mathcal{G}} R_g(t) / [\sum_{g \in \mathcal{G}} N_g(t) + R_g^q(t)]$ |
| ICU-utilization($t$) | The percentage of ICU beds occupied at time $t$ |
| infection-rate-new-seven-day-avg($t$) | The rate of new infections averaged over the last seven days, i.e., $\frac{1}{7} \sum_{\tau \in [t-6, t]} \sum_{g \in \mathcal{G}} [\sigma E_g(\tau)] / [\sum_{g \in \mathcal{G}} N_g(\tau) + R_g^q(\tau)]$ |
| infection-rate-new-elderly-seven-day-avg($t$) | The rate of new infections in elderly groups averaged over the last seven days, i.e., $\frac{1}{7} \sum_{\tau \in [t-6, t]} [\sum_{g \in \mathcal{G} : g \geq 60y.o.} \sigma E_g(\tau)] / [\sum_{g \in \mathcal{G} : g \geq 60y.o.} N_g(\tau) + R_g^q(\tau)]$ |
| ICU-daily-admissions($t$) | New daily admissions into the ICU at time $t$, i.e., the sum of the second and third term in the right-hand-side of (EC.8) over all age groups |
| ICU-daily-admissions-seven-day-avg($t$) | Seven-day average of new daily admissions into the ICU, i.e., ICU-daily-admissions($t$) averaged over the last seven days |

In addition, we also report in Figure EC.5 the tree of depth five trained using the features described above. Beyond appearing in the first four levels of the tree (which is consistent with Figure 10), it is worth noting that the econ-to-contacts-ratio continues to be used for splits in the fifth level, which further supports the importance of that feature.

Lastly, we perform a few other tests of feature importance. Figure EC.6 reports permutation importance for a very high depth (equal to 10) tree. While at this higher depth there are more features with non-zero permutation importance, econ-to-contacts-ratio continues to overwhelmingly dominate the others. We also train another set of depth-four trees to verify this finding through a different means than permutation feature importance. Specifically, we train a tree with *only* the non-targeted features and trees with (all) the non-targeted features plus a single targeted feature.

We do this to see which targeted feature most improves goodness-of-fit versus the non-targeted tree. Figure EC.7 shows the econ-to-contacts-ratio dominates the other trees, and is very close in RMSE to the tree using all features reported in Figure 10.

**Figure EC.5.**
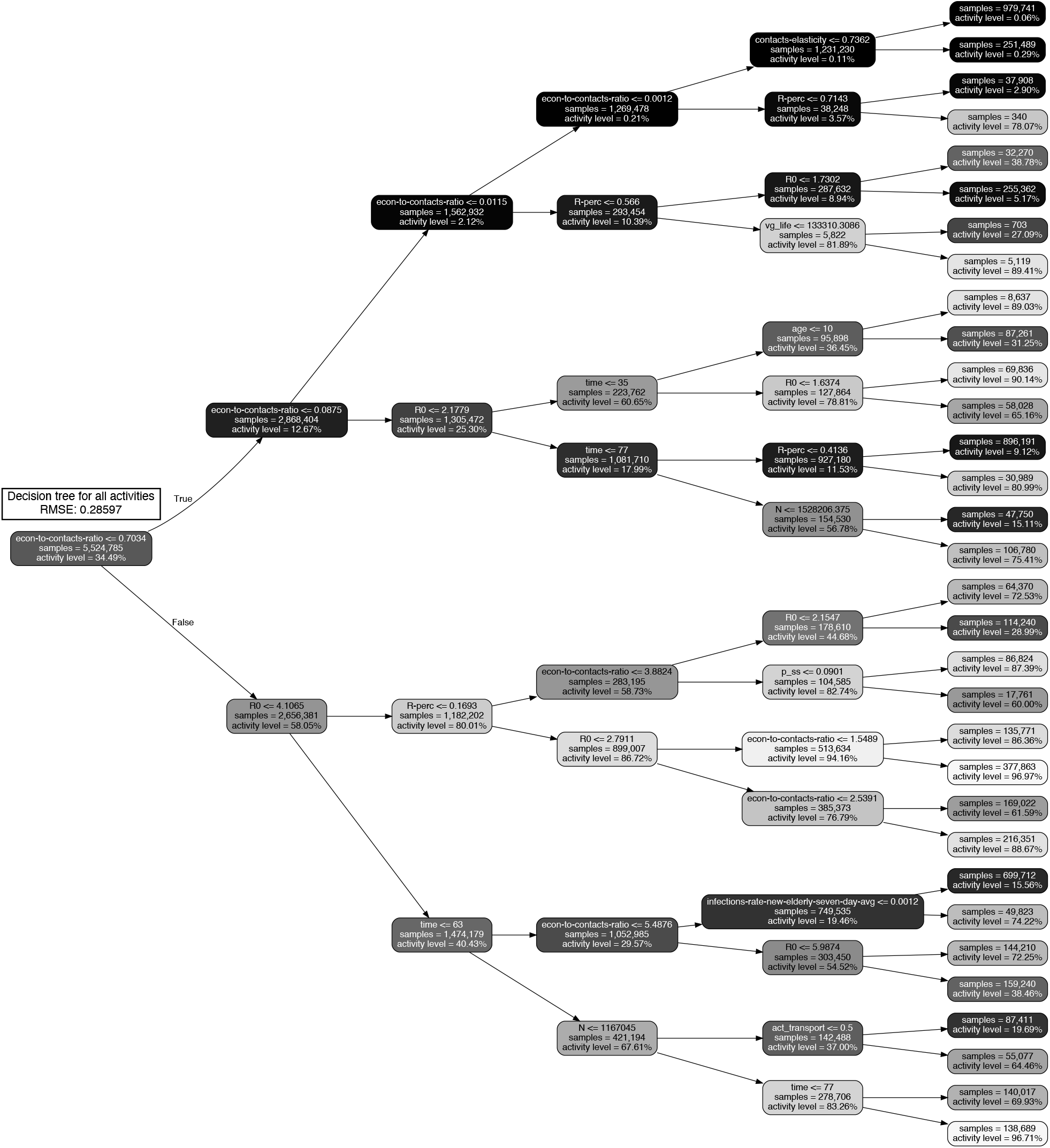
Decision tree of depth five approximating the optimized ROLD AGE-ACT confinement decisions (trained with 5,524,785 samples), with an optimization horizon of *T*′ = 90 days.

##### EC.6.4.2. Additional Details for the Regression Decision Trees for the Gains of Dual Targeting

The training set consists of 14,039 data samples (i.e., one sample for each problem instance), where we include several features, and targets corresponding to the relative gains of ROLD AGE-ACT over ROLD NO-TARGET and over ROLD ACT. The considered features are listed in Table EC.12. Note that the economic features are calculated as aggregated statistics (standard deviation) of the econ-to-contacts-ratio over group-activity pairs. When calculating these statistics, we only use relevant (age group, activity) pairs, where an activity is relevant for an age group if that age group engages in that activity, as detailed in Table EC.8.

**Figure EC.6.**
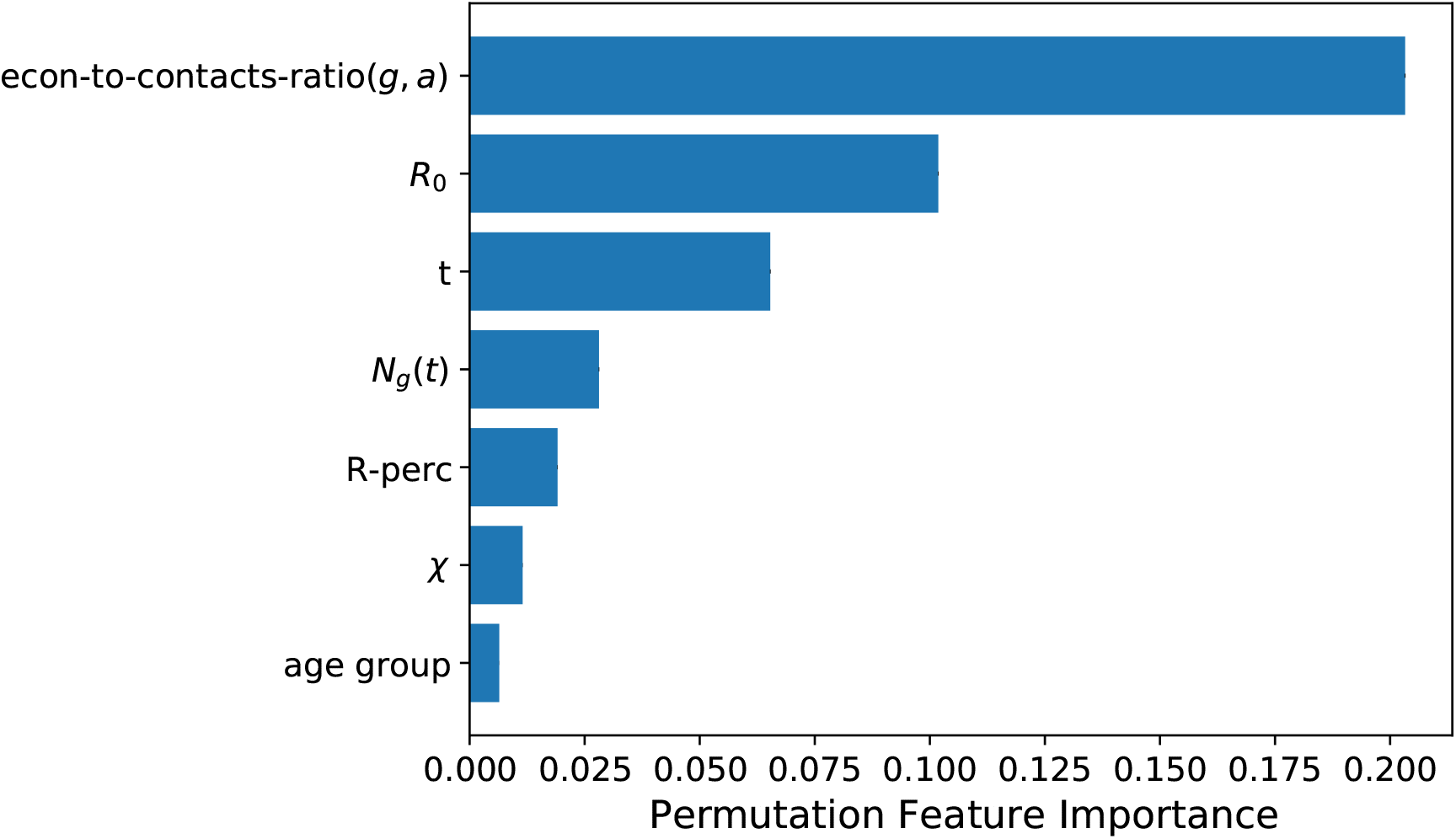
Permutation feature importance for different variables, for the depth-10 decision tree for the ROLD AGE-ACT policy. We show all variables with importance score above 0.005.

**Figure EC.7.**
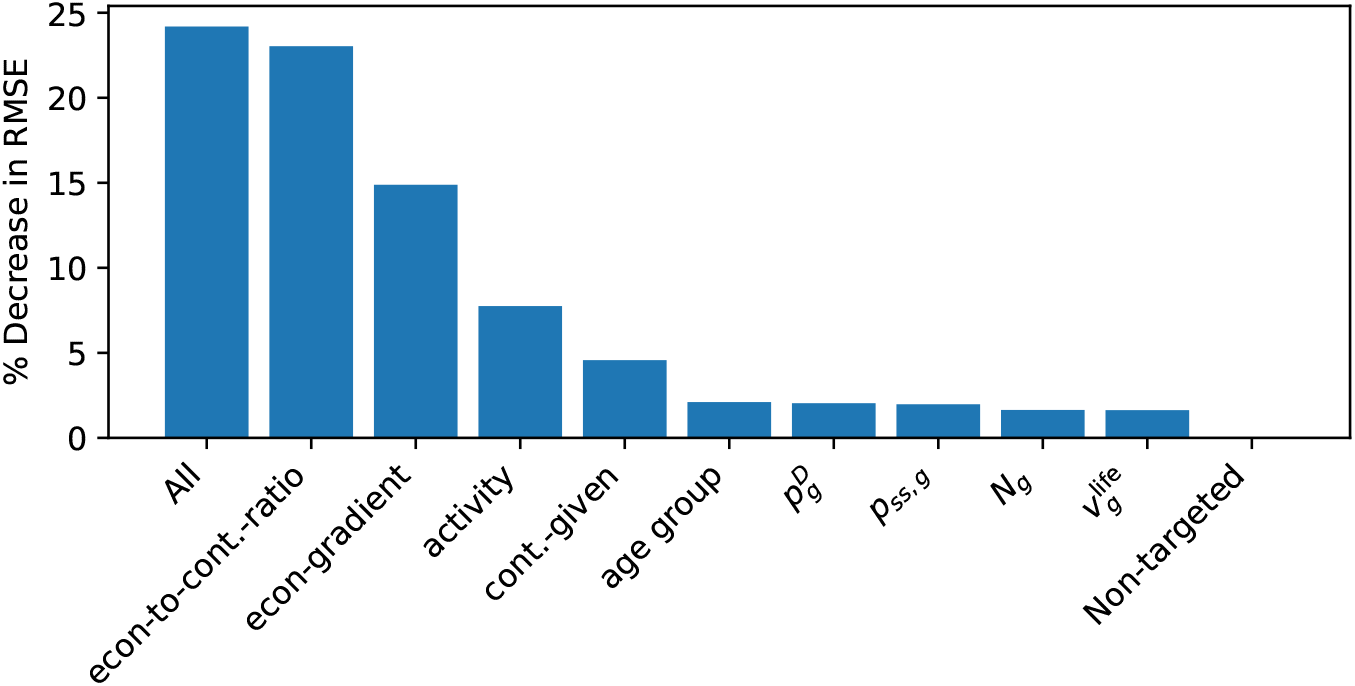
Percent decrease in RMSE for the single targeted feature trees and the all features tree, versus the tree using only non-targeted features, for the ROLD AGE-ACT policy.

Using this data as training set, we then employ the CART algorithm (implemented in the scikit-learn Python library) to train regression decision trees to predict the relative gains of ROLD AGE-ACT over ROLD NO-TARGET, and ROLD AGE-ACT over ROLD ACT, as a function of the considered features, using the mean-squared-error (MSE) criterion as a goodness of fit metric. We use 100 as the minimum number of data points in order to split a node.

We report in Figures EC.8 and EC.9 the trees of depth five trained using the features described above, for the gains of AGE-ACT over NO-TARGET, and AGE-ACT over ACT, respectively.

**Table EC.12.** Features used for fitting trees for gains of dual targeting

| Feature Name | Description |
| --- | --- |
| $R_0$ | Basic reproduction number |
| contacts-elasticity | The percentage drop in total contacts when activity levels drop by 50% |
| $\chi$ | Cost of death |
| severity-scaling | Multiplicative scaling of probabilities $p_{ss,g}$ for all groups $g$ |
| $K^{\text{ICU}}$ | ICU capacity |
| econ-to-contacts-ratio-activity-stdev | The standard deviation of $\text{econ-to-contacts-ratio}(g, a, t_0)$ across relevant age groups for an activity $a$. We include this feature for each activity $a \in \{\text{work, school, transport, leisure, other}\}$. |
| econ-to-contacts-ratio-agegroup-stdev | The standard deviation of $\text{econ-to-contacts-ratio}(g, a, t_0)$ across relevant activities for an age group $g$. We include this feature for each age group $g \in \mathcal{G}$. |

### EC.7. Stylized Model of Linearized Dynamics

This section generates further insight into the structure of the confinement policies produced by ROLD by examining a significantly simpler version of the problem we study, for which an analytical characterization is possible. Our goal is to understand how the optimal ROLD policy depends on various problem primitives (such as economic parameters, contact matrices, etc.), which in turn will confirm our selection of features for training the tree policies in Section 7 and the prominence of econ-to-contacts-ratio.

#### Simplified SEIR model

We consider a simplified compartmental model in which there is a single population group engaging in a single activity, and there are only susceptible (*S*), exposed (*E*), infectious (*I*), recovered (*R*) and deceased (*D*) compartments. Also for the sake of simplicity, we consider the *continuous* time dynamics of this model, namely:

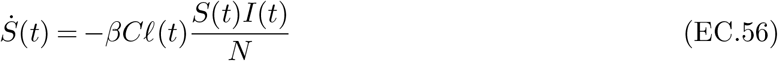

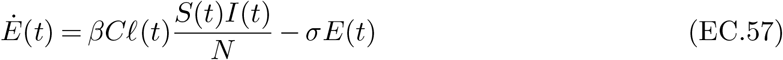

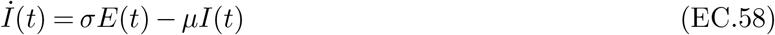

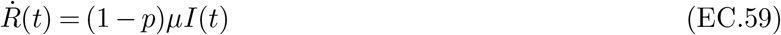

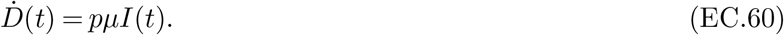

We pause to explain the new dynamical equations. Since there is a single group and activity, the new infections term *βCℓ*(*t*)*S*(*t*)*I*(*t*) is significantly simplified. Here, *β* is the transmission rate and *C* is the rate at which social contacts occur. Furthermore, *σ* and *μ* are the transition rates defined analogously to our original model. Since in this stylized model we remove hospitalized states, we have a direct transition from *I* to *R* or *D*; we denote by *p* the probability that an individual dies given that they are infectious. We denote by *N* the total population size.

#### Activity levels

The control is the activity level of the population and is one-dimensional, so we denote it by *ℓ*, in analogy to the original model. We simplify the economic model by taking the economic value as a linear function of the activity level *ℓ*, i.e., *wℓ*, where *w* is the economic value generated per capita and per unit time under no confinement. Given the same cost of death parameter *χ*, the objective is to maximize:18.

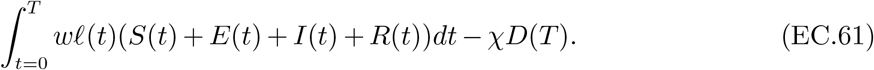

**Figure EC.8.**
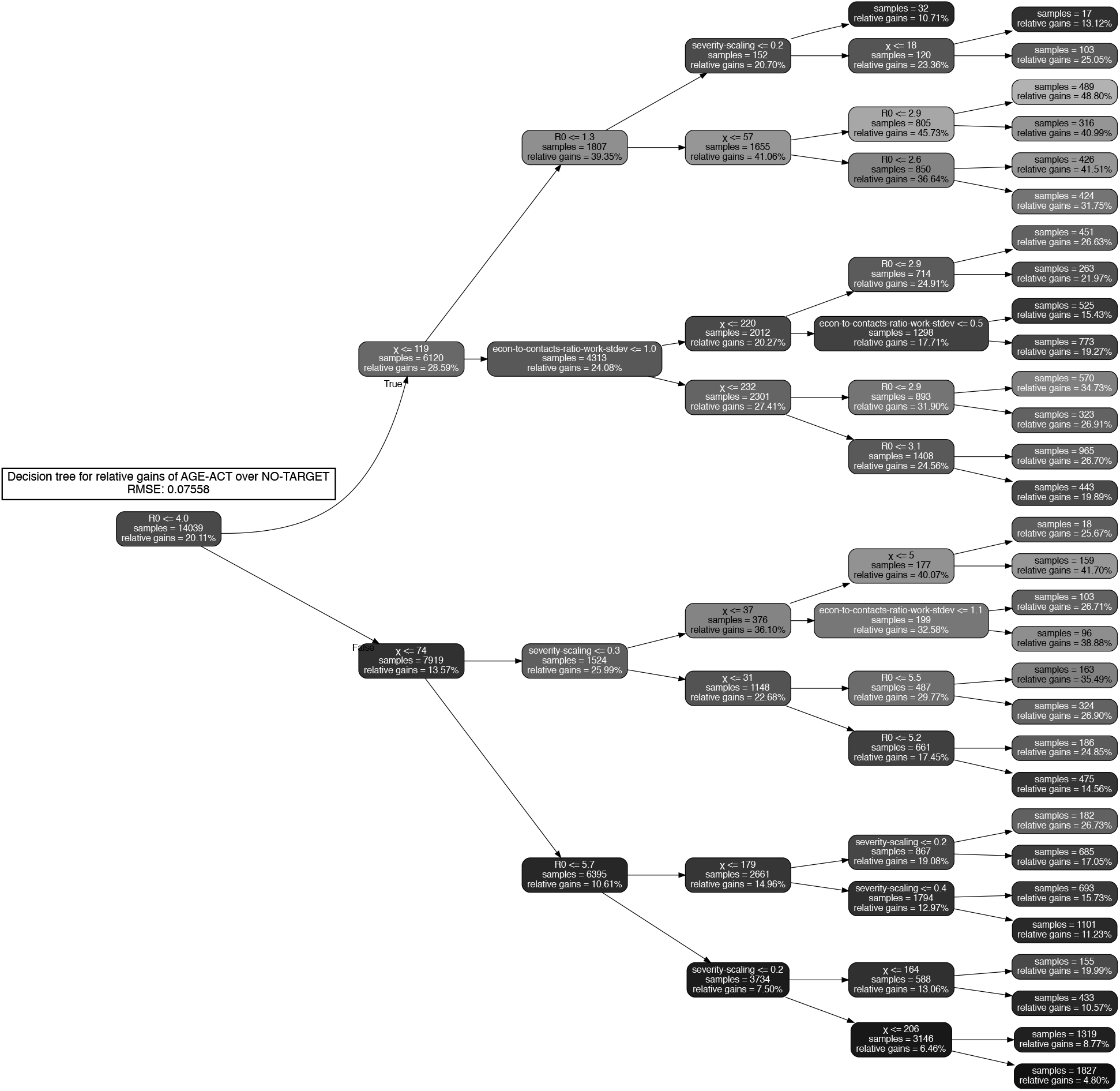
Decision tree of depth five approximating the relative gains of the dual-targeted ROLD AGE-ACT over ROLD NO-TARGET for the total loss, trained on a total of 14,039 problem instances with an optimization horizon of *T*^′^ = 90 days. The nodes are color-coded based on the relative gains, with darker colors corresponding to smaller gains.

#### Solving the linearized system

To facilitate the analysis, we make the assumption that throughout the time horizon and for any activity level, *S*(*t*) ≈ *N*. In that case, the new infections term can be approximated as

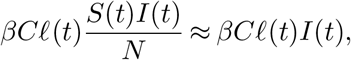

leading to the approximate dynamics:

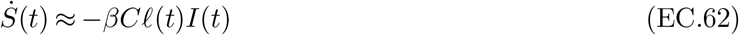

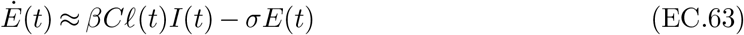

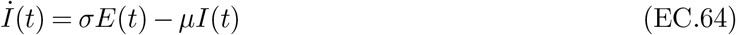

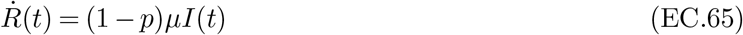

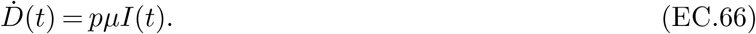

**Figure EC.9.**
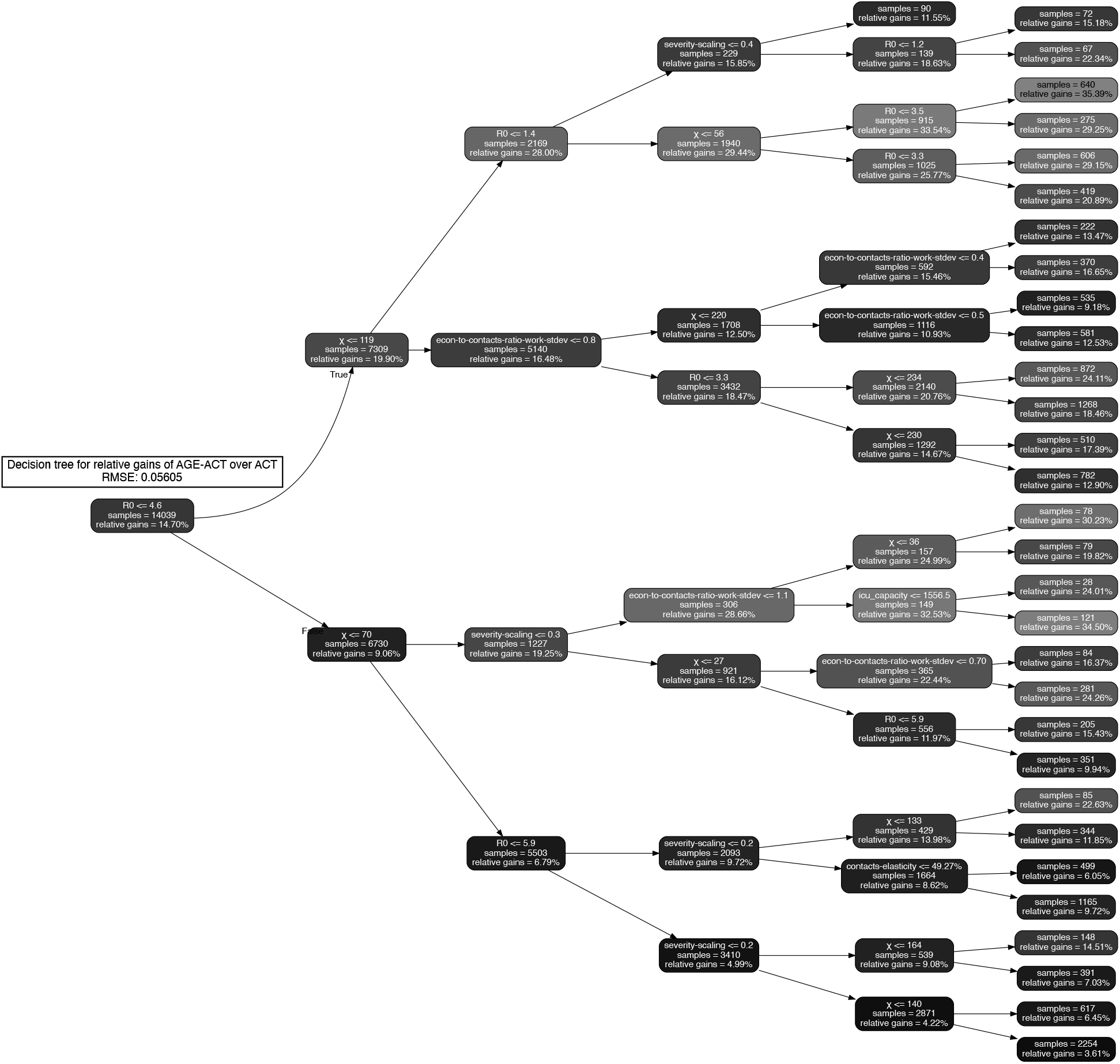
Decision tree of depth five approximating the relative gains of the dual-targeted ROLD AGE-ACT over ROLD ACT for the total loss, trained on a total of 14,039 problem instances with an optimization horizon of *T*^*′*^= 90 days. The nodes are color-coded based on the relative gains, with darker colors corresponding to smaller gains.

In compact notation, these approximate SEIR dynamics can be written as 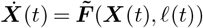, where ***X***(*t*) = (*S*(*t*), *E*(*t*), *I*(*t*), *R*(*t*), *D*(*t*)) and the function 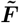 captures (EC.62)-(EC.66). Additionally, under the assumption *S*(*t*) ≈ *N*, the objective simplifies to:

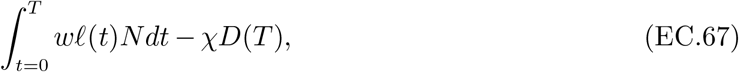

since *S*(*t*) + *E*(*t*) + *I*(*t*) + *R*(*t*) ≈*N*.

With respect to this linearized dynamic and approximate objective, the control problem is:

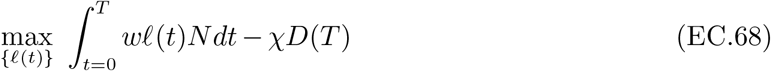

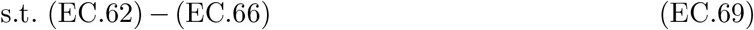

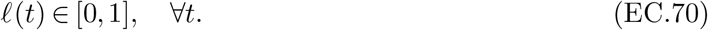

Note that the control problem under the simplified model does not include capacity constraints, as we have removed the hospitalized states from the model.

Although the dynamics in (EC.62) - (EC.66) are linear, the bilinear terms coming from multiplying *ℓ* with the SEIR states still make the problem difficult to solve. We thus proceed with a linearization of the problem which mimics the workings of the ROLD algorithm in Section 4, and characterize the optimal policy for this linearized problem. Analogously to Section 4, consider a nominal time-invariant control 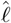, and a nominal trajectory 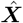. We build a linear approximation of the system dynamics as:

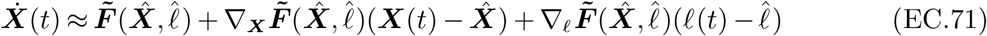

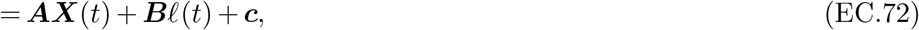

where

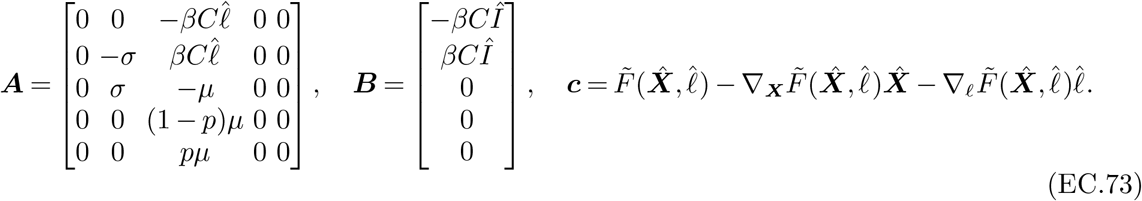

This allows us to express the state at any *t* as a function of the confinement decision *ℓ*(*τ*) for all 0 ≤ *τ* ≤ *t*. In particular, the solution of the dynamical system 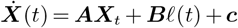 is

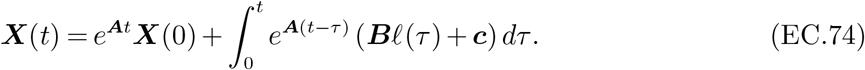

Note that we can readily write the objective (EC.68) as maximizing

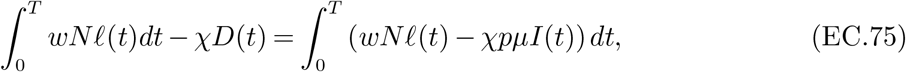

without resorting to any Taylor approximations for linearization.

By plugging in the solution for the dynamical system (EC.74), we have

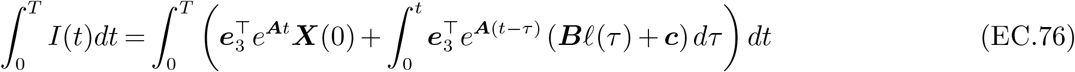

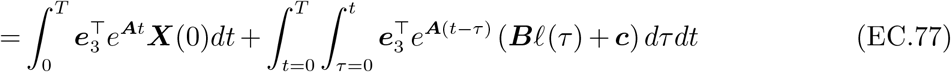

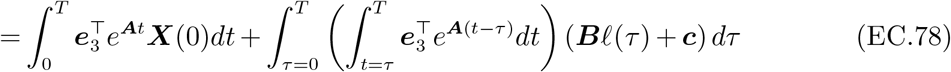

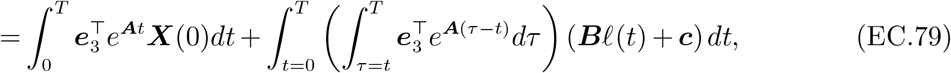

where ***e***_3_ := [0, 0, 1, 0, 0].

We can now rewrite the objective in (EC.75) as

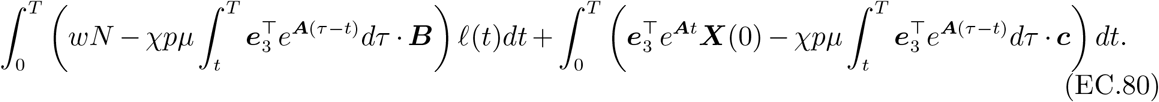

The second summand is a constant with respect to the control. The coefficient of *ℓ*(*t*) in the integral in the first summand calculates to

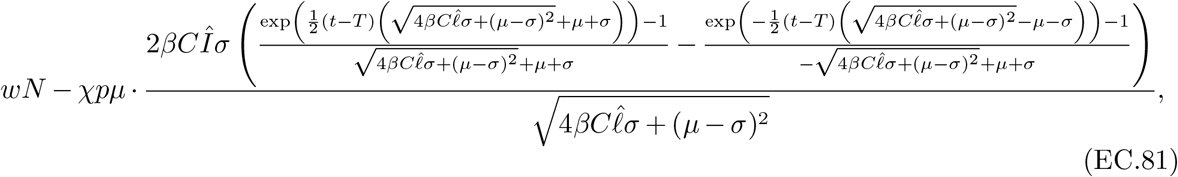

and the optimal policy is to set *ℓ* to 1 if and only if the expression above is non-negative. It is perhaps most useful to focus on understanding the resulting policy for the case that 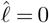, in which case the coefficient of *ℓ*(*t*) takes a simpler form equal to

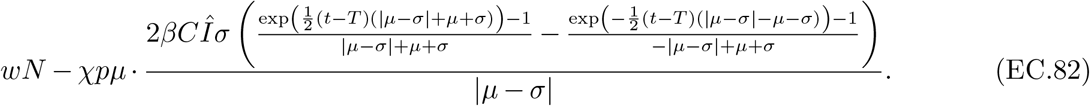

The optimal decision then is

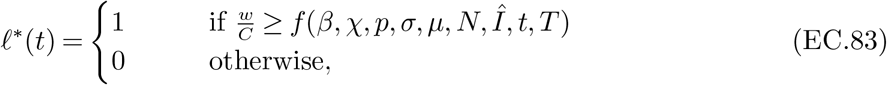

where

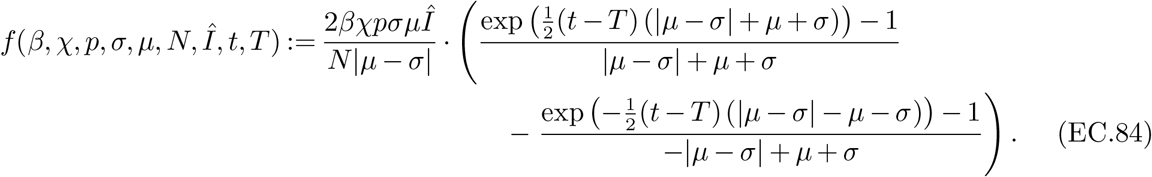

(We note that19. *f* (*β, χ, p, σ, γ, N, Î, t, T*) ≥0 for 0 ≤*t* ≤*T*.)

Inequality (EC.83) uncovers a natural logic for the confinement decisions in this linearized formulation. Specifically, the left-hand side exactly corresponds to the econ-to-contacts-ratio that we identified in Section 7: it is given by the gradient of (per-capita) economic value generated with respect to the level of activity *ℓ*(*t*)20. divided by the rate of social contacts generated. Thus, the optimal policy is governed by a threshold on the value of the econ-to-contacts-ratio, allowing normal activity levels (*ℓ*(*t*) = 1) when the econ-to-contacts-ratio exceeds the threshold and completely confining the entire population (*ℓ*(*t*) = 0) otherwise. The threshold, given by the function *f* (*β, χ, p, σ, γ, N, Î, t, T*), is increasing in parameters such as the probability of infection given a contact *β*, the probability of death given infection *p*, and the cost of death *χ*, and is decreasing in the size of the overall population *N*, which matches intuition.

We remark that the simple threshold policy relying on econ-to-contacts-ratio emerges when we impose 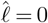 in (EC.81); for general 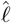, the policy still follows a threshold rule based on the marginal economic value exceeding a function that depends on problem parameters and the rate of contacts, but the simple ratio no longer emerges in the rule.

There are two conclusions we highlight from this stylized model analysis. First, even with significant simplifications such as a single group and activity, the policies output by ROLD or similar procedures are quite complex and elusive to completely characterize in closed form. Second, there are however interesting regimes where these policies intuitively depend on a quantity resembling the econ-to-contacts-ratio. In this light, selecting the group-activity pairs to be confined in decreasing order of their econ-to-contacts-ratios, as our trees from Section 7 do, naturally parallels the policy in (EC.83). This provides some theoretical backing to the feature that the trees consider to be important in explaining ROLD’s decisions.

### EC.8. Details on the Practically Implementable Policies

#### EC.8.1. Retrieving Participation Rates from the Time-Use Survey Data

The time-use survey data by INSEE (2010) follows a data format that is harmonized with EU standards for time-use surveys. Each row in the main data file is a “diary” that corresponds to a specific day of a specific survey participant. Among other information, a diary has information on the self-reported location of the participant, as well as the self-reported main activity of the participant, per 10-minute interval (for a total of 144 intervals over the 24-hour day). The survey participant selects both the reported location and the reported main activity from a given menu of options.

Most of the reported locations in the diaries map naturally into one of our six activities in the set 𝒜= {work, transport, leisure, school, home, other }. However, for four of the reported locations in the diaries (“Unspecified location/transport mode”, “Unspecified location (not travelling)”, “Workplace or school”, “Other specified location (not travelling)”), we use supplemental information on the main activity pursued, as well as on the activity (i.e., employment) status of the individual, to decide on the exact mapping. In particular:

- For reported locations “Unspecified location/transport mode”, “Unspecified location (not travelling)”, “Other specified location (not travelling)”, we map the location to one or more of our activities in set 𝒜 on the basis of the reported time-use activity, using our judgment on which of our activities are highly relevant for the respective pair of reported location-activity in the survey.
- For reported location “Workplace or school”:
  — If the activity (employment) status of the survey participant is “Pupil, student, further training, unpaid traineeship”, then for most reported activities we assign to our activity school. For a few reported activities that relate to transport, we assign to both school and transport.
  — If the activity (employment) status of the survey participant is “Employed full-time” or “Employed part-time”, then for most reported activities we assign to our activity work. For a few reported activities, we assign either to school; work and school; work and transport; or school and transport.
  — Otherwise (i.e., if the activity status of the survey participant is neither of the above), then for most reported activities we assign to both our activities work and school. For a few reported activities, we assign either to only work; only school; work and transport; or school and transport.

We detail how we calculate the participation rates 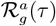 in Algorithm 7. When the proposed mapping assigns to more than one of our activities, then we split the 10 minutes of the corresponding 10-minute interval equally across the proposed activities. We note that the time-use survey data does not include responses from participants aged below 10 y.o., and therefore for our 0-9 y.o. age group we propose an interpolation from survey participants aged 10, 11, and 12 y.o.

##### Algorithm 7

Calculating Participation Rates 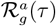

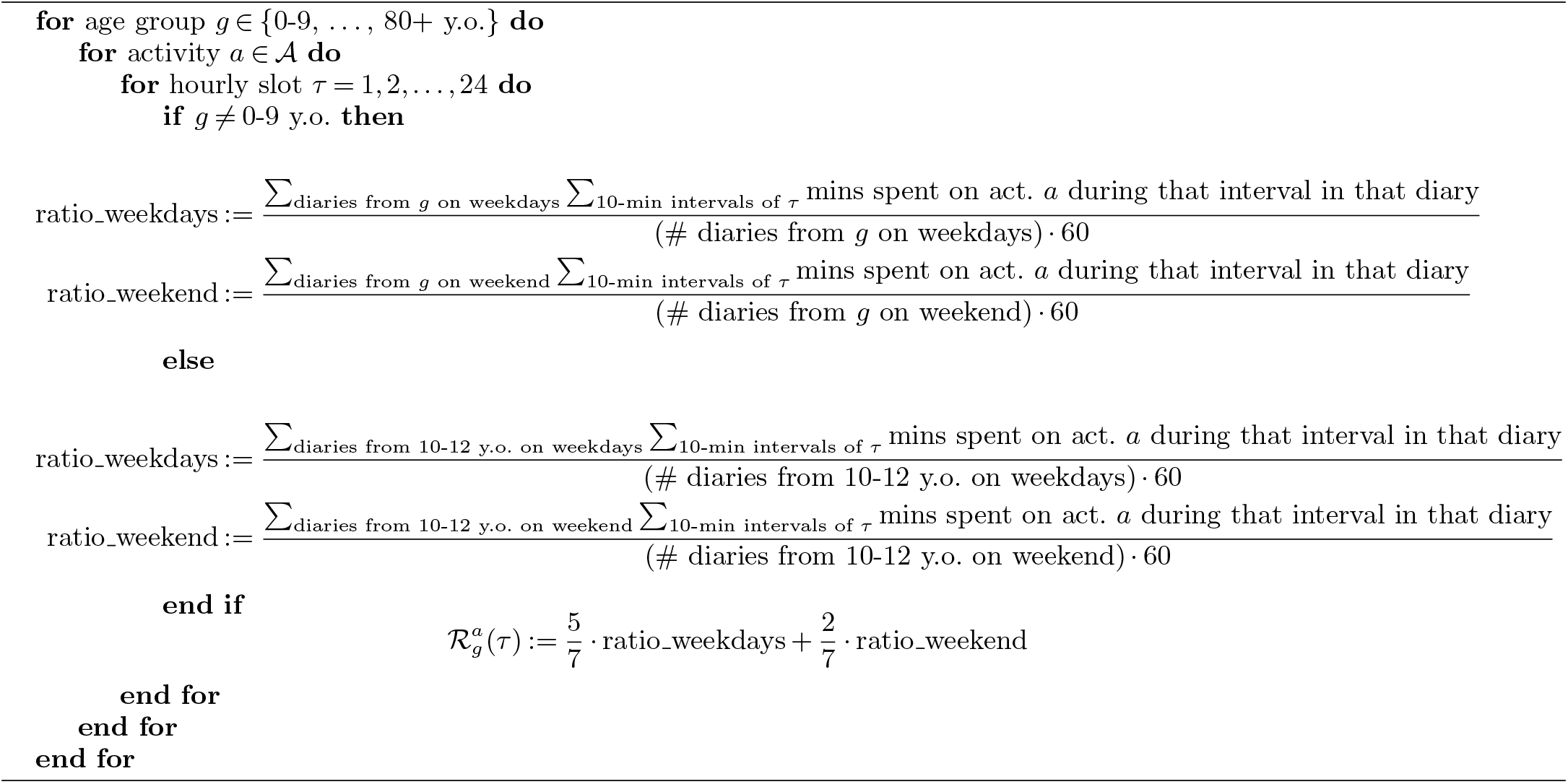

#### EC.8.2. Details on the Gradient Descent Procedure for Optimizing Curfews

We used the ‘trust-constr’ method in the SCIPY-OPTIMIZE Python library, and employed its default parameter settings. For a given problem instance, we initialized the optimization algorithm using two different strategies:

- Random initialization: For each activity, we sample the start and duration of the curfew uniformly at random, both from the 0 − 24 hours interval. We repeat this exercise for a total of 120 iterations.
- ROLD ACT solution initialization, randomly perturbed: For each activity, we draw an i.i.d. Bernoulli variable with probability of success *p*, where *p* is a parameter explained below. In case of success, the curfew is initialized at the ROLD ACT solution. In case of failure, we sample the start and duration of the curfew uniformly at random, both from the 0 − 24 hours interval. We vary the parameter *p* in the range [0, 1/6, 2/6, 3/6, 4/6, 5/6] and repeat this exercise for 20 iterations for each of the values for parameter *p*. This gives a total of 120 iterations.

Each of the iterations is run for 24 hours using two strategies:

- Full completion: The algorithm runs until local optimality, which, on average, is achieved after eight hours.
- Partial completion: The algorithm runs for two hours and is restarted with a new random seed for a total of 12 times.

In total, we used 600 CPUs, each running for 24 hours, where the processors used in the cluster were Intel E5-2640v4, Intel 5118, and AMD 7502. We note that we did not optimize the code for more efficient computation.

#### EC.8.3. Time-Average Aggregate Activity Levels for Age Groups

We calculate the time-average activity level of each age group, averaged over the activities relevant to that age group (Table EC.8), and weighting different activities by the respective participation rates. Specifically, for AGE-ACT, ACT, and NO-TARGET, the proposed calculation is the one in (EC.55) (and similarly for the other age groups). For CURFEW, the proposed calculation just replaces 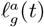 in these equations with 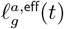 from (15).

#### EC.8.4. Details on Limited Compliance Policies with Age-Group-Specific Compliance Fractions

We use the estimates for age-group-specific non-compliance probabilities by Ganslmeier et al. (2022) to estimate a relative scaling of compliance probabilities across age groups. Ganslmeier et al. (2022) estimate non-compliance probabilities based on data from the first COVID-19 lockdown in the UK. Because the age groups used by Ganslmeier et al. (2022) are different from ours, to find the compliance probability *p*_*g*_ for an age group *g* ∈ 𝒢 we take the conservative approach of using the minimum over the compliance probabilities of the age groups of Ganslmeier et al. (2022) that intersect with age group *g*. For our 0-9 y.o. age group, for which Ganslmeier et al. (2022) do not estimate compliance, we use the compliance probability from Ganslmeier et al. (2022) for their youngest group, 18-24 y.o. Table EC.13 summarizes the age-group specific compliance probabilities we interpolate for our age groups. Based on these, we estimate a relative scaling between compliance probabilities of different age groups, assuming that the relative scaling in France and in particular Île-de-France would be similar to the relative scaling in the UK.

**Table EC.13.** Fraction of the population that complies with recommended restrictions by age group as interpolated from study by Ganslmeier et al. (2022)

| Age Group $g$ (y.o.) | Compliance Fraction $p_g$ (%) |
| --- | --- |
| 0-9 | 96.10 |
| 10-19 | 96.10 |
| 20-29 | 94.66 |
| 30-39 | 94.66 |
| 40-49 | 95.50 |
| 50-59 | 96.27 |
| 60-69 | 96.40 |
| 70-79 | 96.45 |
| 80+ | 96.45 |

Restricting to that fixed relative scaling, we refer to the resulting effective activity levels as the ROLD COMP-NH(*p*) policy, where NH stands for non-homogeneous across age groups, and where *p* is the compliance fraction for the least compliant age group according to the scaling. We look at the performance of the ROLD COMP-NH(*p*) policy as the compliance fraction *p* varies. The results are similar to the results for limited compliance policies with a population-wide compliance fraction (ROLD COMP(*p*)), and are summarized in Figure EC.10.

---

**Figure EC.10.**
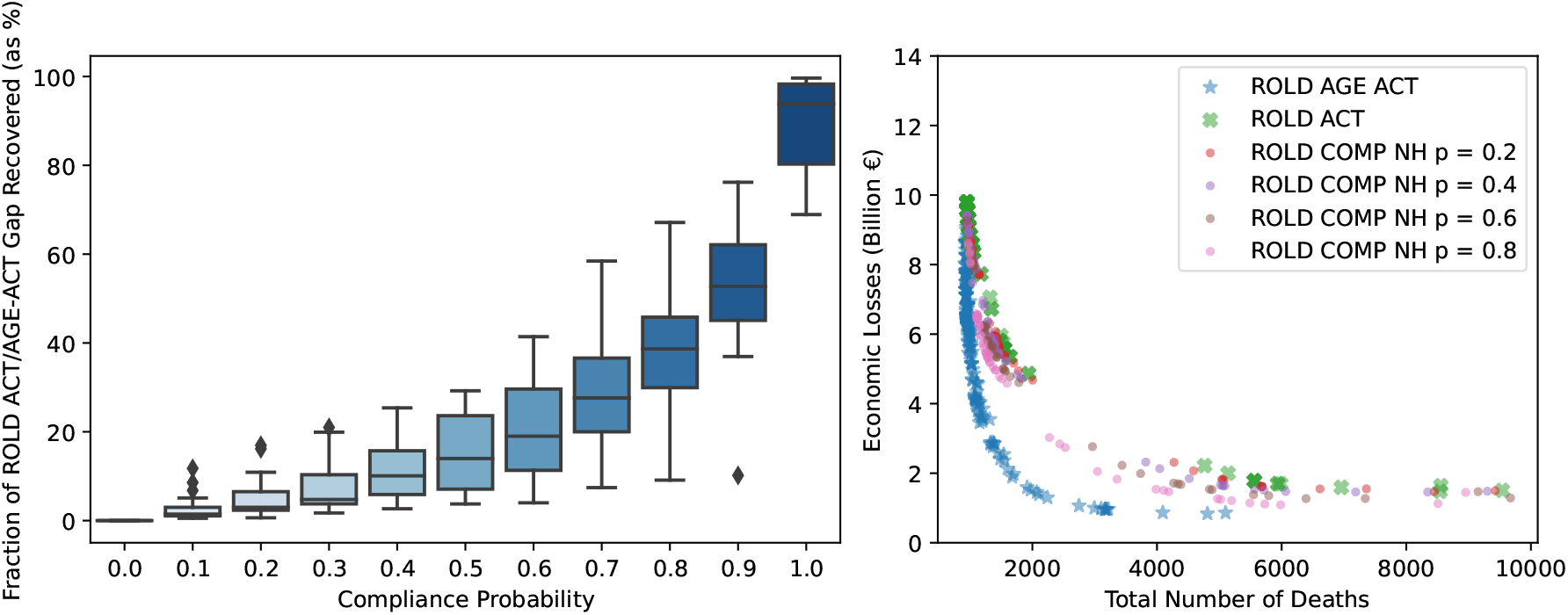
The performance of limited compliance policies with age-group-specific compliance, for different compliance levels. We estimate a relative scaling of compliance fractions across age groups using the estimates in Ganslmeier et al. (2022). Keeping the relative scaling across age groups fixed, and denoting the compliance fraction for the least compliant age group by *p*, we look at the performance of ROLD COMP-NH(*p*) as *p* varies. In the left panel, the boxplot at each *p* summarizes the fraction of the ROLD ACT/AGE-ACT performance gap that ROLD COMP-NH(*p*) recovers, across different values of the cost of death *χ*. We include 201 distinct values of *χ* from 0 to 1000*×*. The right panel illustrates the total number of deaths and the economic losses generated by ROLD COMP-NH(*p*) for different compliance levels *p*, compared to ROLD AGE-ACT and ROLD ACT. Each marker corresponds to a different problem instance parameterized by *χ*.

**Table EC.14:**
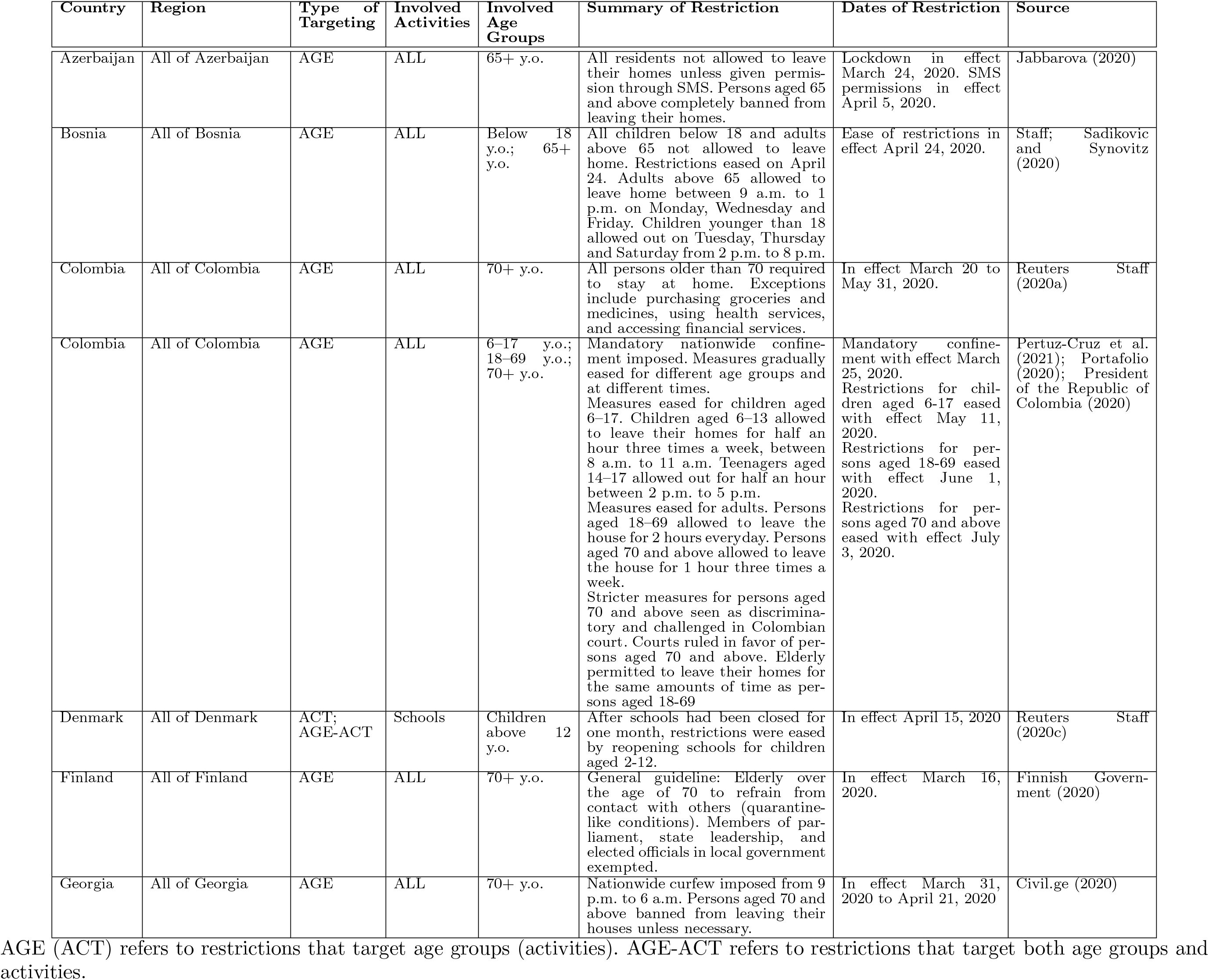
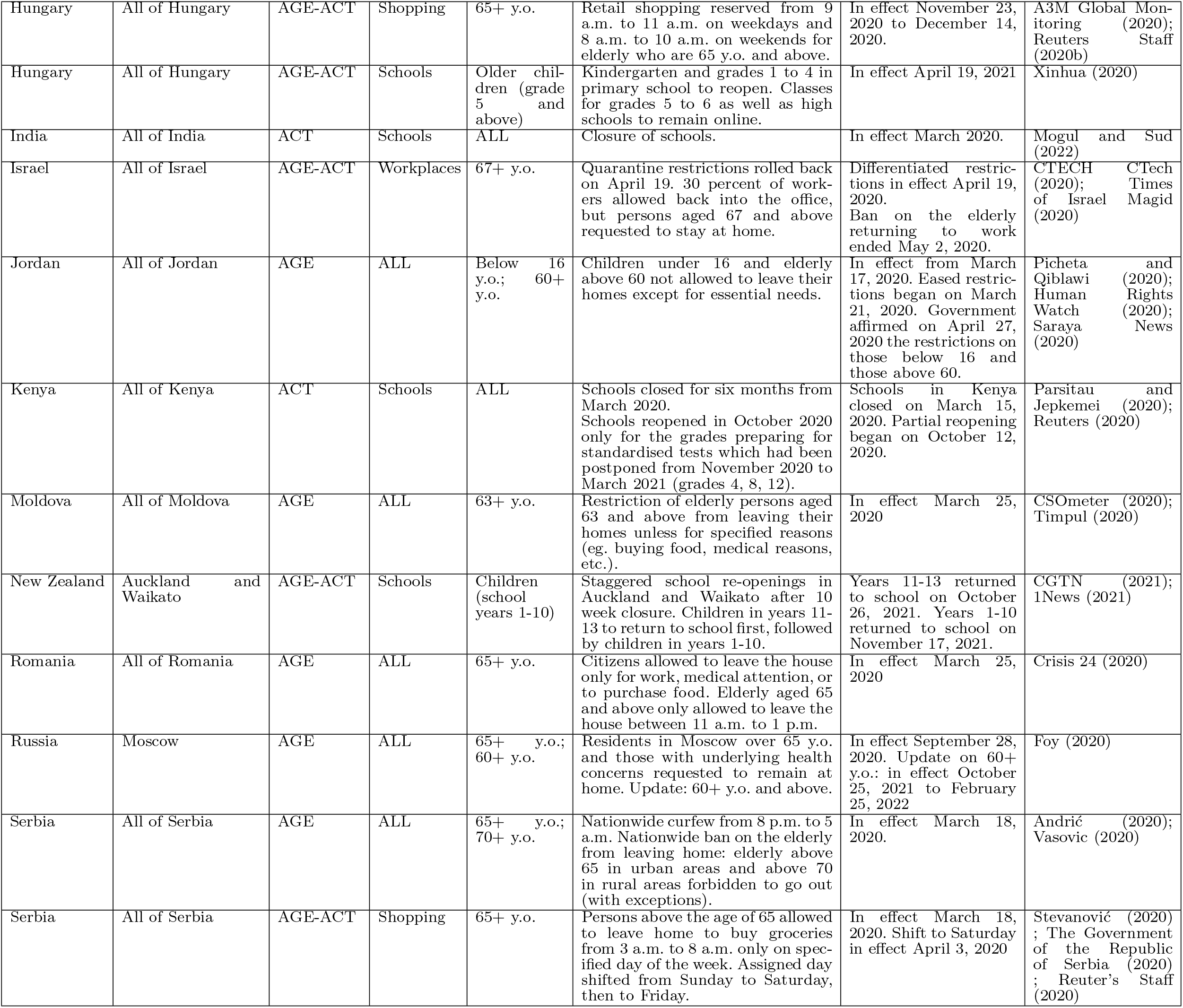
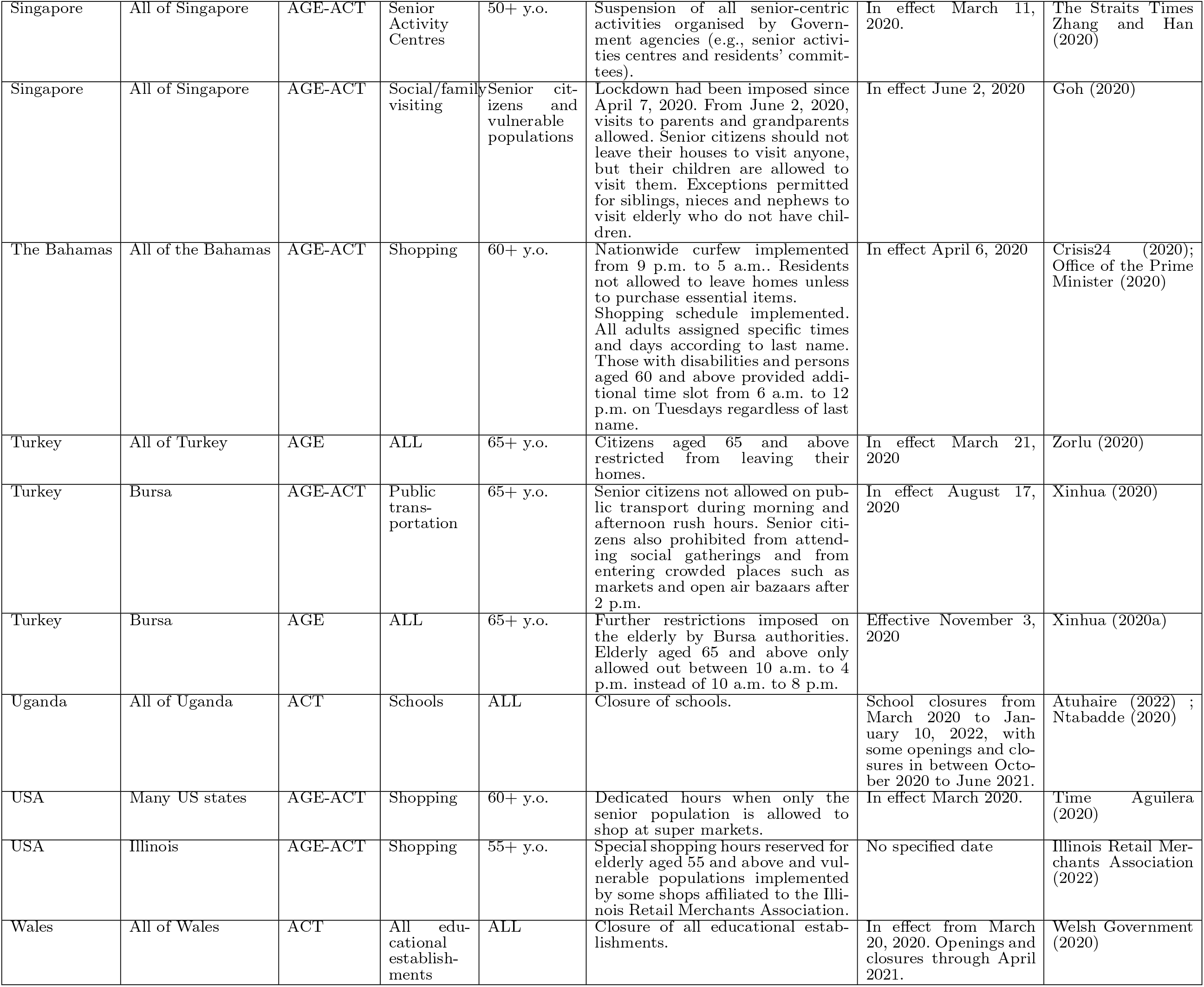
Examples of enforced targeted restrictions

**Table EC.15:** Examples of recommended targeted restrictions

| Country | Region | Type of Targeting | Involved Activities | Involved Age Groups | Summary of Recommended Restriction | Dates of Restriction | Source |
| --- | --- | --- | --- | --- | --- | --- | --- |
| China | All of China | AGE-ACT | Travel | Elderly | Elderly and high-risk groups advised against travelling during China's week long May holiday | In effect April 23, 2020 | Xinhua (2020b); National Working Committee on Children and Women under State Council (2020) |
| Croatia | All of Croatia | AGE | ALL | 60+ y.o. | Elderly above the age of 60 advised to avoid crowded places. Routine visits to the doctor should be skipped unless urgent. | In effect March 3, 2020 | Government of the Republic of Croatia (2020) |
| Czech Republic | All of Czech Republic | AGE | ALL | 60+ y.o. | Seniors advised to leave the house as little as possible. | In effect March 20, 2020 | Ministry of Health (2020) |
| Ireland | All of Ireland | AGE | ALL | 70+ y.o. | Elderly above the age of 70 and medically vulnerable persons advised to cocoon (i.e., stay home as much as possible). | In effect March 27, 2020 | Department of Health (2020); McGreevy and Gleeson (2020) |
| New Zealand | All of New Zealand | AGE | ALL | Older people | Older and vulnerable people encouraged to "hunker down". | Peak of Omicron outbreak, early 2022. | ? |
| Thailand | All of Thailand | AGE | ALL | Elderly | Elderly and their primary caregivers advised to stay at home as much as possible | In effect April 6, 2020 | Department of Medical Services |
| The Bahamas | All of the Bahamas | AGE-ACT | Work | 65-74 y.o. | Elderly between the age of 65-74 recommended to work from home. | In effect March 29, 2020 | Office of the Prime Minister Communications Unit (2020) |
| The Bahamas | All of the Bahamas | AGE | ALL | 75+ y.o. | Elderly above the age of 75 asked not to leave home. | In effect March 29, 2020 | Office of the Prime Minister Communications Unit (2020) |

AGE (ACT) refers to restrictions that target age groups (activities). AGE-ACT refers to restrictions that target both age groups and activities.

## References

Acemoglu D, Chernozhukov V, Werning I, Whinston MD (2020) Optimal targeted lockdowns in a multi-group SIR model. Working Paper 27102, NBER, URL http://dx.doi.org/10.3386/w27102.

Adelman D (2020) Thousands of lives could be saved in the US during the COVID-19 pandemic if states exchanged ventilators. Health Affairs 39(7):1247–1252, URL http://dx.doi.org/10.1377/hlthaff.2020.00505, pMID: 32352846.

Aguilera J (2020) Some supermarkets are launching senior-only hours during the coronavirus pandemic. Not all retailers think that’s a good idea. URL https://time.com/5804574/grocery-store-chains-seniors-coronavirus/, accessed January 12, 2021.

Ahn HS, Silberholz J, Song X, Wu X (2021) Optimal covid-19 containment strategies: Evidence across multiple mathematical models. Available at SSRN 3834668.

Alagoz O, Sethi AK, Patterson BW, Churpek M, Alhanaee G, Scaria E, Safdar N (2021) The impact of vaccination to control COVID-19 burden in the United States: A simulation modeling approach. PLOS ONE 16(7):1–12, URL http://dx.doi.org/10.1371/journal.pone.0254456.

Alagoz O, Sethi AK, Patterson BW, Churpek M, Safdar N (2020) Effect of timing of and adherence to social distancing measures on COVID-19 burden in the United States. Annals of Internal Medicine 174(1):50–57, URL http://dx.doi.org/10.7326/M20-4096.

Alvarez FE, Argente D, Lippi F (2020) A simple planning problem for COVID-19 lockdown. Working Paper 26981, National Bureau of Economic Research, URL http://dx.doi.org/10.3386/w26981.

Anderson RM, May RM (1992) Infectious Diseases of Humans: Dynamics and Control (Oxford University Press).

Bastani H, Drakopoulos K, Gupta V, Vlachogiannis I, Hadjicristodoulou C, Lagiou P, Magiorkinis G, Paraskevis D, Tsiodras S (2021) Efficient and targeted COVID-19 border testing via reinforcement learning. Nature 599(7883):108–113, URL http://dx.doi.org/10.1038/s41586-021-04014-z.

Bemporad A (2006) Model predictive control design: New trends and tools. Proceedings of the 45th IEEE Conference on Decision and Control, 6678–6683 (IEEE).

Béraud G, et al. (2015) The French connection: The first large population-based contact survey in France relevant for the spread of infectious diseases. PLOS ONE 10(7):1–22.

Bertsimas D, Boussioux L, Cory-Wright R, Delarue A, Digalakis V, Jacquillat A, Kitane DL, Lukin G, Li M, Mingardi L, Nohadani O, Orfanoudaki A, Papalexopoulos T, Paskov I, Pauphilet J, Lami OS, Stellato B, Bouardi HT, Carballo KV, Wiberg H, Zeng C (2021) From predictions to prescriptions: A data-driven response to COVID-19. Health Care Management Science 24(2):253–272, URL http://dx.doi.org/10.1007/s10729-020-09542-0.

Bertsimas D, Ivanhoe JK, Jacquillat A, Li ML, Previero A, Lami OS, Bouardi HT (2020) Optimizing vaccine allocation to combat the COVID-19 pandemic. medRxiv.

Birge JR, Candogan O, Feng Y (2020) Reducing economic losses with targeted closures. Working Paper 2020-57, University of Chicago, Becker Friedman Institute for Economics.

Boloori A, Saghafian S (2023) Health and economic impacts of lockdown policies in the early stage of COVID-19 in the United States. Service Science 0(0):null, URL http://dx.doi.org/10.1287/serv.2023.0321.

Bose S, Souyris S, Mukherjee UK, Ivanov A, Seshadri S, III ACE, Xu Y (2021) Control of epidemic spreads via testing and lock-down. Technical report.

Brandeau ML, Zaric GS, Richter A (2003) Resource allocation for control of infectious diseases in multiple independent populations: beyond cost-effectiveness analysis. Journal of Health Economics 22(4):575–598, ISSN 0167-6296, URL http://dx.doi.org/https://doi.org/10.1016/S0167-6296(03)00043-2.

Camacho EF, Alba CB (2013) Model predictive control (Springer science & business media).

Chang S, Pierson E, Koh PW, Gerardin J, Redbird B, Grusky D, Leskovec J (2020) Mobility network models of COVID-19 explain inequities and inform reopening. Nature 589:1–6.

Chhatwal J, Dalgic O, Mueller P, Adee M, Xiao Y, Ladd MA, Linas BP, Ayer T (2020) PIN68 COVID-19 simulator: An interactive tool to inform COVID-19 intervention policy decisions in the United States. Value in Health 23:S556, URL http://dx.doi.org/10.1016/j.jval.2020.08.909.

Cipriano LE, Haddara WMR, Zaric GS, Enns EA (2021) Impact of university re-opening on total community COVID-19 burden. PLOS ONE 16(8):1–16, URL http://dx.doi.org/10.1371/journal.pone.0255782.

Cot C, Cacciapaglia G, Sannino F (2021) Mining Google and Apple mobility data: Temporal anatomy for COVID-19 social distancing. Scientific reports 11(1):1–8.

Di Domenico L, Pullano G, Sabbatini CE, Böelle PY, Colizza V (2020) Impact of lockdown on COVID-19 epidemic in Île-de-France and possible exit strategies. BMC medicine 18(1):1–13.

Du Z, Xu X, Wu Y, Wang L, Cowling BJ, Meyers L (2020) Serial interval of COVID-19 among publicly reported confirmed cases. Emerging Infectious Diseases 26(6).

Duque D, Morton DP, Singh B, Du Z, Pasco R, Meyers LA (2020) Timing social distancing to avert unmanageable COVID-19 hospital surges. Proceedings of the National Academy of Sciences 117(33):19873–19878.

Dutta R, Gomes SN, Kalise D, Pacchiardi L (2021) Using mobility data in the design of optimal lockdown strategies for the COVID-19 pandemic. PLoS Computational Biology 17(8):e1009236.

El Housni O, Sumida M, Rusmevichientong P, Topaloglu H, Ziya S (2020) Can testing ease social distancing measures? Future evolution of COVID-19 in NYC. arXiv preprint arXiv:2005.14700.

Evgeniou T, Fekom M, Ovchinnikov A, Porcher R, Pouchol C, Vayatis N (2020) Epidemic models for personalised covid-19 isolation and exit policies using clinical risk predictions. medRxiv.

Favero CA, Ichino A, Rustichini A (2020) Restarting the economy while saving lives under COVID-19. C.E.P.R Discussion Paper 14664, CEPR.

Fotouhi H, Mori N, Miller-Hooks E, Sokolov V, Sahasrabudhe S (2021) Assessing the effects of limited curbside pickup capacity in meal delivery operations for increased safety during a pandemic. Transportation Research Record 2675(5):436–452, URL http://dx.doi.org/10.1177/0361198121991840.

Foy H (2020) Russian businesses prepare for fresh lockdowns as Covid-19 cases soar. The Financial Times URL https://www.ft.com/content/f0696f19-fe0b-425c-8e73-3703bd44dec3, accessed January 12, 2021.

French Government (2020) Données hospitalières relatives à l’épidémie de COVID-19. https://www.data.gouv.fr/fr/datasets/donnees-hospitalieres-relatives-a-lepidemie-de-covid-19/, accessed October 21, 2020.

Ganslmeier M, Van Parys J, Vlandas T (2022) Compliance with the first uk covid-19 lockdown and the compounding effects of weather. Scientific Reports 12(1):3821, URL http://dx.doi.org/10.1038/s41598-022-07857-2.

Giordano G, Colaneri M, Di Filippo A, Blanchini F, Bolzern P, De Nicolao G, Sacchi P, Colaneri P, Bruno R (2021) Modeling vaccination rollouts, SARS-CoV-2 variants and the requirement for non-pharmaceutical interventions in Italy. Nature Medicine 27(6):993–998.

Goldstein JR, Cassidy T, Wachter KW (2021) Vaccinating the oldest against COVID-19 saves both the most lives and most years of life. Proceedings of the National Academy of Sciences 118(11), ISSN 0027-8424.

Google (2020) COVID-19 community mobility report. https://www.google.com/covid19/mobility/, accessed October 21, 2020.

Harrison S (2020) Coronavirus: Ireland’s restrictions eased for over 70s. BBC URL https://www.bbc.com/news/uk-northern-ireland-52547169, accessed January 12, 2021.

Huang HC, Araz OM, Morton DP, Johnson GP, Damien P, Clements B, Meyers LA (2017) Stockpiling ventilators for influenza pandemics. Emerging Infectious Diseases 23(6):914–921, URL http://dx.doi.org/10.3201/eid2306.161417.

Ilin C, Annan-Phan S, Tai XH, Mehra S, Hsiang S, Blumenstock JE (2021) Public mobility data enables COVID-19 forecasting and management at local and global scales. Scientific reports 11(1):1–11.

INSEE (2010) Emploi du temps (version pour Eurostat). 2009-2010, INSEE [producer], ADISP [distributor].

Kanbur N, Ankgül S (2020) Quaranteenagers: A single country pandemic curfew targeting adolescents in Turkey. Journal of Adolescent Health 67(2).

Kaplan EH (2020) Om forum—covid-19 scratch models to support local decisions. Manufacturing & Service Operations Management 22(4):645–655, URL http://dx.doi.org/10.1287/msom.2020.0891.

Kucharski AJ, et al. (2020) Effectiveness of isolation, testing, contact tracing, and physical distancing on reducing transmission of SARS-CoV-2 in different settings: a mathematical modelling study. The Lancet Infectious Diseases 20(10):1151–1160.

Lehot M, Borgne BL (2020) Covid-19 : ces chiffres qui montrent que Paris a dépassé le seuil d’alerte maximale depuis le 25 septembre. Franceinfo Accessed October 5, 2020.

Li ML, Bouardi HT, Lami OS, Trikalinos TA, Trichakis N, Bertsimas D (2022) Forecasting COVID-19 and analyzing the effect of government interventions. Operations Research 71(1):184–201.

Linas BP, Xiao J, Dalgic OO, Mueller PP, Adee M, Aaron A, Ayer T, Chhatwal J (2022) Projecting COVID-19 Mortality as States Relax Nonpharmacologic Interventions. JAMA Health Forum 3(4):e220760–e220760, ISSN 2689-0186, URL http://dx.doi.org/10.1001/jamahealthforum.2022.0760.

Magid J (2020) Cabinet approves removal of age restriction for those returning to work. The Times of Israel URL https://www.timesofisrael.com/liveblog-may-2-2020/, accessed January 12, 2021.

Mandal S, Das H, Deo S, Arinaminpathy N (2021) Combining serology with case-detection, to allow the easing of restrictions against SARS-CoV-2: a modelling-based study in India. Scientific Reports 11(1):1835, URL http://dx.doi.org/10.1038/s41598-021-81405-2.

Mas-Colell A, Whinston MD, Green JR, et al. (1995) Microeconomic theory, volume 1 (Oxford University Press New York).

Matrajt L, Eaton J, Leung T, Brown ER (2021) Vaccine optimization for COVID-19: Who to vaccinate first? Science Advances 7(6):eabf1374, URL http://dx.doi.org/10.1126/sciadv.abf1374.

Mehrotra S, Rahimian H, Barah M, Luo F, Schantz K (2020) A model of supply-chain decisions for resource sharing with an application to ventilator allocation to combat COVID-19. Naval Research Logistics (NRL) 67(5):303–320, URL http://dx.doi.org/https://doi.org/10.1002/nav.21905.

Morris DH, Rossine FW, Plotkin JB, Levin SA (2021) Optimal, near-optimal, and robust epidemic control. Communications Physics 4(1):1–8.

Navabi-Shirazi M, El Tonbari M, Boland N, Nazzal D, Steimle LN (2022) Multicriteria course mode selection and classroom assignment under sudden space scarcity. Manufacturing & Service Operations Management 24(6):3252–3268, URL http://dx.doi.org/10.1287/msom.2022.1131.

Pataro IML, et al. (2021) A control framework to optimize public health policies in the course of the COVID-19 pandemic. Scientific Reports 11(1):13403.

Patel MD, Rosenstrom E, Ivy JS, Mayorga ME, Keskinocak P, Boyce RM, Hassmiller Lich K, Smith I Raymond L, Johnson KT, Delamater PL, Swann JL (2021) Association of Simulated COVID-19 Vaccination and Nonpharmaceutical Interventions With Infections, Hospitalizations, and Mortality. JAMA Network Open 4(6):e2110782–e2110782, ISSN 2574-3805, URL http://dx.doi.org/10.1001/jamanetworkopen.2021.10782.

Perakis G, Singhvi D, Skali Lami O, Thayaparan L (2021) COVID-19: A multipeak SIR based model for learning waves and optimizing testing. Available at SSRN 3817680.

Prem K, Cook AR, Jit M (2017) Projecting social contact matrices in 152 countries using contact surveys and demographic data. PLoS computational biology 13(9):e1005697.

Prem K, et al. (2020) The effect of control strategies to reduce social mixing on outcomes of the COVID-19 epidemic in Wuhan, China: a modelling study. The Lancet Public Health 5(5):e261–e270.

Reuters (2021) France goes under nationwide 6pm curfew as Covid-19 death toll surpasses 70,000. France24 URL https://www.france24.com/en/europe/20210116-france-set-for-nationwide-6pm-curfew-in-effort-to-stem-covid-19, accessed January 12, 2021.

Reuters Staff (2020) Bosnian region eases lockdown on seniors, children after court ruling. Reuters URL https://www.reuters.com/article/us-health-coronavirus-bosnia-idUSKCN2261RA, accessed January 12, 2021.

Rosenstrom ET, Ivy JS, Mayorga ME, Swann JL (2022) Could earlier availability of boosters and pediatric vaccines have reduced impact of COVID-19? 2022 Winter Simulation Conference (WSC), 1–12, URL http://dx.doi.org/10.1109/WSC57314.2022.10015236.

Salje H, et al. (2020) Estimating the burden of SARS-CoV-2 in France. Science 38.

Tiirinki H, et al. (2020) COVID-19 pandemic in Finland–preliminary analysis on health system response and economic consequences. Health policy and technology 9(4):649–662.

Wellenius GA, et al. (2020) Impacts of state-level policies on social distancing in the United States using aggregated mobility data during the COVID-19 pandemic. arXiv e-prints arXiv–2004.

Wille L, Van Hoang T, Funk S, Coletti P, Beutels P, Hens N (2020) SOCRATES: an online tool leveraging a social contact data sharing initiative to assess mitigation strategies for COVID-19. BMC Research Notes 13(293).

Xiong C, Hu S, Yang M, Luo W, Zhang L (2020) Mobile device data reveal the dynamics in a positive relationship between human mobility and COVID-19 infections. Proceedings of the National Academy of Sciences 117(44):27087–27089.

Young HP (1994) Equity (Princeton University Press).

Yuan Yx (2015) Recent advances in trust region algorithms. Mathematical Programming 171:249–281.

Zaric GS, Brandeau ML (2001) Resource allocation for epidemic control over short time horizons. Mathematical Biosciences 171(1):33–58, ISSN 0025-5564, URL http://dx.doi.org/https://doi.org/10.1016/S0025-5564(01)00050-5.

Zaric GS, Brandeau ML (2002) Dynamic resource allocation for epidemic control in multiple populations. Mathematical Medicine and Biology: A Journal of the IMA 19(4):235–255, URL http://dx.doi.org/10.1093/imammb/19.4.235.

## E-companion References

Banque de France (2020a) Point sur la conjoncture française à fin avril 2020. https://www.banque-france.fr/sites/ efault/files/media/2020/06/10/point-conjoncture_avril-2020.pdf, accessed June 19, 2020.

Banque de France (2020b) Point sur la conjoncture française à fin mai 2020. https://www.banque-france.fr/sites/default/files/media/2020/06/11/point-conjoncture-09_juin-2020-20200609.pdf, accessed June 19, 2020.

Bi Q, et al. (2020) Epidemiology and transmission of COVID-19 in 391 cases and 1286 of their close contacts in Shenzhen, China: a retrospective cohort study. The Lancet Infectious Diseases 20(8).

Diekmann O, Heesterbeek JAP, Roberts MG (2010) The construction of next-generation matrices for compartmental epidemic models. Journal of The Royal Society Interface 7(47):873–885.

INSEE (2016a) T402 : Salaire brut en équivalent temps plein, par secteur d’activité, région et département. https://www.insee.fr/fr/statistiques/fichier/4204500/T402.xls, accessed June 19, 2020.

INSEE (2016b) T404 : Salaire brut enéquivalent temps plein, par tranche d’âge simplifiée, catégorie socioprofessionnelle simplifiée et région. https://www.insee.fr/fr/statistiques/fichier/4204500/T404.xls, accessed June 19, 2020.

INSEE (2019) Au quatrieme trimestre 2019, le taux de chômage passe de 8,5 % a 8,1 %. https://www.insee.fr/fr/statistiques/4309346, accessed June 19, 2020.

INSEE (2020) Estimation de population par département, sexe etâge quinquennal - années 1975 à 2020. https://www.insee.fr/fr/statistiques/fichier/1893198/estim-pop-dep-sexe-aq-1975-2020.xls, accessed June 19, 2020.

Mohammad H (2020) Coronavirus : un habitant de Bobigny considéré comme le nou-veau “patient zéro”. France Bleu, URL https://www.francebleu.fr/infos/sante-sciences/coronavirus-un-habitant-de-bobigny-considere-comme-le-nouveau-patient-zero-1588796619, accessed January 9, 2021.

Perasso A (2018) An introduction to the basic reproduction number in mathematical epidemiology. ESAIM: Proceedings and Surveys 62:123–138.

Salje H, et al. (2020) Estimating the burden of SARS-CoV-2 in France. Science 38.

## References for Examples of Targeted Restrictions

1News (2021) Auckland high school seniors prepare to return to class. 1News URL https://www.1news.co.nz/2021/10/25/auckland-high-school-seniors-prepare-to-return-to-class/, accessed April 29, 2022.

A3M Global Monitoring (2020) Covid-19 pandemic - hungary. A3M Global Monitoring URL https://global-monitoring.com/gm/page/events/epidemic-0001988.5I3CRSSqviVB.html?lang=en, accessed April 30, 2022.

Aguilera J (2020) Some supermarkets are launching senior-only hours during the coronavirus pandemic. not all retailers think that’s a good idea. Time URL https://time.com/5804574/grocery-store-chains-seniors-coronavirus/, accessed April 28, 2022.

Andrić G (2020) Corona virus: “you work all day, so you run to get home before curfew - how serbia stopped for 50 days”. BBC URL https://www-bbc-com.translate.goog/serbian/cyr/srbija-60733573?_x_tr_sl=sr&_x_tr_tl=en&_x_tr_hl=en&_x_tr_pto=sc, accessed April 29, 2022.

Atuhaire P (2022) Uganda schools reopen after almost two years of covid closure. BBC URL https://www.bbc.com/news/world-africa-59935605, accessed April 29, 2022.

CGTN (2021) Schools in new zealand’s auckland to reopen from november 17. CGTN URL https://newsaf.cgtn.com/news/2021-11-11/Schools-in-New-Zealand-s-Auckland-to-reopen-from-November-17-155kca6Frdm/index.html, accessed on April 29, 2022.

Civilge (2020) Covid-19: Georgia announces nationwide lockdown, partial curfew. UNA.ge URL https://civil.ge/archives/344761, accessed April 30, 2022.

Crisis 24 (2020) Romania: Government announces lockdown measures on March 25 /update 2. Crisis 24 URL https://crisis24.garda.com/alerts/2020/03/romania-government-announces-lockdown-measures-on-march-25-update-2, accessed April 30, 2022.

Crisis24 (2020) Bahamas: Additional restrictions for shopping introduced as of April 6 /update 4. Crisis24 URL https://crisis24.garda.com/alerts/2020/04/bahamas-additional-restrictions-for-shopping-introduced-as-of-april-6-update-4, accessed April 30, 2022.

CSOmeter (2020) State of emergency in the republic of moldova. CSOmeter URL https://csometer.info/updates/state-emergency-republic-moldova, accessed April 30, 2022.

CTech (2020) Israel rolls back covid-19 restrictions as number of sick stabilizes. CTech URL https://www.calcalistech.com/ctech/articles/0,7340,L-3809065,00.html, accessed April 28, 2022.

Department of Health (2020) Guidance on cocooning to protect people over 70 years and those extremely medically vulnerable from covid-19 - updated guidance from 29 june. Government of Ireland URL https://www.gov.ie/en/publication/923825-guidance-on-cocooning-to-protect-people-over-70-years-and-those-extr/?fbclid=IwAR2TkpZADAmZpaFd6MCkmk2OElQzZ1gEuIq9BOPF3p9K2AF2yRU29PuqIKY, accessed April 30, 2022.

Department of Medical Services (????) Guidelines for caring for the elderly during the covid-19 outbreak.

Finnish Government (2020) Government, in cooperation with the president of the republic, declares a state of emergency in finland over coronavirus outbreak. Finnish Government URL https://valtioneuvosto.fi/en/-/10616/hallitus-totesi-suomen-olevan-poikkeusoloissa-koronavirustilanteen-vuoksi, accessed April 30, 2022.

Foy H (2020) Russian businesses prepare for fresh lockdowns as covid-19 cases soar. The Financial Times URL https://www.ft.com/content/f0696f19-fe0b-425c-8e73-3703bd44dec3, accessed April 30, 2022.

Goh YH (2020) Limited visits to parents or grandparents to be allowed from June 2 after circuit breaker. Straits Times URL https://www.straitstimes.com/singapore/limited-visits-to-parents-or-grandparents-to-be-allowed-from-june-2-seniors-urged-to-stay, accessed April 30, 2022.

Government of the Republic of Croatia (2020) Coronavirus protection measures. Government of the Republic of Croatia URL https://vlada.gov.hr/coronavirus-protection-measures/28950, accessed April 30, 2022.

Human Rights Watch (2020) Jordan: State of emergency declared. Human Rights Watch URL https://www.hrw.org/news/2020/03/20/jordan-state-emergency-declared, accessed April 30, 2022.

Illinois Retail Merchants Association (2022) Special shopping hours for vulnerable populations. IRMA URL https://irma.org/covid-19-senior-shopping/, accessed April 29, 2022.

Jabbarova A (2020) Azerbaijanis required to send sms notification before leaving their homes. Global Voices URL https://globalvoices.org/2020/04/10/azerbaijanis-required-to-send-sms-notification-before-leaving-homes/, accessed April 30, 2022.

Magid J (2020) Cabinet approves removal of age restriction for those returning to work. Times of Israel URL https://www.timesofisrael.com/liveblog-may-2-2020/, accessed April 28, 2022.

McGreevy R, Gleeson C (2020) Cocooning is advisory, not mandatory, government confirms. The Irish Times URL https://www.irishtimes.com/news/ireland/irish-news/cocooning-is-advisory-not-mandatory-government-confirms-1.4229569, accessed April 30, 2022.

Ministry of Health (2020) Senioři by měli co nejvíce omezit vychézení ven. Ministry of Health of the Czech Republic URL https://koronavirus.mzcr.cz/semiori-by-meli-co-nejvice-omezit-vychazeni-ven/, accessed April 30, 2022.

Mogul R, Sud V (2022) After more than 600 days shut out, delhi’s students just want to go back to school. CNN URL https://edition.cnn.com/2022/01/27/india/india-delhi-schools-reopen-600-days-intl-hnk/index.html, accessed April 29, 2022.

National Working Committee on Children and Women under State Council (2020) Experts from the chinese center for disease control and prevention remind that high-risk groups such as pregnant women, the elderly, and patients with chronic diseases are not recommended to travel. National Working Committee on Children and Women under State Council URL http://www.nwccw.gov.cn/2020-04/24/content_283708.htm, accessed April 30, 2022.

Ntabadde C (2020) “i wish i could have some magic, i would just say let schools open right away”. UNICEF URL https://www.unicef.org/uganda/stories/i-wish-i-could-have-some-magic-i-would-just-say-let-schools-open-right-away, accessed April 29, 2022.

Office of the Prime Minister (2020) Food shopping schedule. Office of the Prime Minister URL https://www.bahamas.gov.bs/wps/wcm/connect/df5a4569-9c5b-40e4-85f3-d764584aef5c/Statement+-+Food+shopping+schedule.pdf?MOD=AJPERES, accessed April 30, 2022.

Office of the Prime Minister Communications Unit (2020) Prime minister announces three new cases of covid-19 and plans to extend state of emergency for eight days. The Government of the Bahamas URL https://tinyurl.com/3vppjmm6, accessed April 30, 2022.

Parsitau DS, Jepkemei E (2020) How school closures during covid-19 further marginalize vulnerable children in kenya. Brookings; Education Plus Development URL https://www.brookings.edu/blog/education-plus-development/2020/05/06/how-school-closures-during-covid-19-further-marginalize-vulnerable-children-in-kenya/#:∼:text=On%20March%2015%2C%202020%2C%20the,devastating%20consequences%20for%20marginalized%20learners., accessed April 30, 2022.

Pertuz-Cruz SL, Molina-Montes E, Rodríguez-Pérez C, Guerra-Hernälndez EJ, Cobos de Rangel OP, Artacho R, Verardo V, Ruiz-Lopez MD, García-Villanova B (2021) Exploring dietary behavior changes due to the covid-19 confinement in colombia: A national and regional survey study. Frontiers in Nutrition URL https://www.frontiersin.org/articles/10.3389/fnut.2021.644800/full#B5, accessed April 29, 2022.

Picheta R, Qiblawi T (2020) Jordan eases lockdown after total curfew leads to chaos. CNN URL https://edition.cnn.com/2020/03/25/middleeast/jordan-lockdown-coronavirus-intl/index.html, accessed April 30, 2022.

Portafolio (2020) Tumban cuarentena obligada para adultos mayores de 70 años. Portafolio URL https://www.portafolio.co/economia/gobierno/tumban-cuarentena-obligatoria-para-adultos-mayores-de-70-anos-coronavirus-en-colombia-hoy-12-agosto-202 accessed April 29, 2022.

President of the Republic of Colombia (2020) Estas son las medidas que se deben tener en cuenta para la salida de los niños durante la cuarentena. President of the Republic of Colombia URL https://id.presidencia.gov.co/Paginas/prensa/2020/Estas-son-las-medidas-que-se-deben-tener-en-cuenta-para-la-salida-de-los-ninos-durante-la-cuarentena-200507.aspx3, accessed April 29, 2022.

Reuters (2020) Kenya partially reopens schools, 6 months after covid shuttered them. Vox URL https://www.voanews.com/a/covid-19-pandemic_kenya-partially-reopens-schools-6-months-after-covid-shuttered-them/6197024.html, accessed April 30, 2022.

Reuters Staff (2020a) Colombia declares coronavirus state of emergency, orders elderly to stay home. Reuters URL https://www.reuters.com/article/us-health-coronavirus-colombia-idUSKBN2150AI, accessed April 30, 2022.

Reuters Staff (2020b) Hungary imposes restricted shopping hours to protect elderly in pandemic. Reuters URL https://www.reuters.com/article/uk-health-coronavirus-hungary-idUKKBN2831OW, accessed April 30, 2022.

Reuters Staff (2020c) Reopening schools in Denmark did not worsen outbreak, data shows. Reuters URL https://www.reuters.com/article/us-health-coronavirus-denmark-reopenig/reopening-schools-in-denmark-did-not-worsen-outbreak-data-shows-idUSKBN2341N7, accessed April 30, 2022.

Reuter’s Staff (2020) Serbia’s elderly venture out for dawn food run. Reuters URL https://www.reuters.com/article/health-coronavirus-serbia-elderly-idUSL8N2BM0A5, accessed April 29, 2022.

Sadikovic M, Synovitz R (2020) Coronavirus in court: Bosnia’s age-based lockdowns are ruled discriminatory. RadioFreeEurope/RadioLiberty URL https://www.rferl.org/a/coronavirus-in-court-bosnia-s-age-based-lockdowns-are-ruled-discriminatory/30574453.html, accessed April 30, 2022.

Saraya News (2020) Adaileh: The government continues to ban children and the elderly from leaving. Saraya News URL https://www.sarayanews.com/article/611788?fbclid=IwAR0Bfhu348B9y0BAnVM6UN5tIOSj-kRa9r3i3wYenXbrIeQSmMN-GgE6c, accessed April 30, 2022.

Staff R (????) Bosnian region eases lockdown on seniors, children after court ruling.

Stevanović K (2020) Corona virus and over 65 in serbia: “it never occurs to me to vampire”. BBC URL https://www.bbc.com/serbian/cyr/srbija-52266134, accessed April 29, 2022.

The Government of the Republic of Serbia (2020) Senior citizens to go grocery shopping on saturday instead of sunday. The Government of the Republic of Serbia URL https://www.srbija.gov.rs/vest/en/153257/senior-citizens-to-go-grocery-shopping-on-saturday-instead-of-sunday.php, accessed April 29, 2022.

Timpul (2020) Drastic decisions /// the government prohibits people over the age of 63 from being outside. it is forbidden for all people to be in public spaces (parks, recreational areas) / quarantine in the villages of bă lceana and sofia. Timpul URL https://timpul.md/articol/decizii-drastice-guvernul-interzice-persoanelor-in-varsta-de-peste-63-ani-aflarea-in-afara-domiciliului-se-interzice-tuturor-persoanelor-aflarea-in-spatii-publice-parcuri-zone-de-agrement-ca. html, accessed April 30, 2022.

Vasovic A (2020) Serbia imposes night curfew, orders elderly indoors. Reuters URL https://www.reuters.com/article/us-health-coronavirus-serbia/serbia-imposes-night-curfew-orders-elderly-indoors-idUSKBN2143XR, accessed April 29, 2022.

Welsh Government (2020) Timeline of school closures during the coronavirus (covid-19) pandemic, march 2020 to april 2021. Welsh Government URL https://gov.wales/timeline-school-closures-during-coronavirus-covid-19-pandemic, accessed April 30, 2022.

Xinhua (2020) Hungary set to cautiously reopen schools, kindergartens. Xinhua URL http://www.xinhuanet.com/english/europe/2021-04/16/c_139885633.htm, accessed April 30, 2022.

Xinhua (2020) Turkey’s bursa province adopts new covid-19 restrictions for elderly. Xinhua URL http://www.xinhuanet.com/english/2020-08/17/c_139297485.htm, accessed April 30, 2022.

Xinhua (2020a) Turkey’s bursa tightens covid-19 lockdown on elderly. Xinhua URL http://www.news.cn/english/2020-11/04/c_139488890.htm, accessed April 29, 2022.

Xinhua (2020b) Xinhua headlines: China’s tourism rebounds during may day holiday as coronavirus eases. Xinhua URL http://www.xinhuanet.com/english/2020-05/05/c_139032433.htm, accessed April 30, 2022.

Zhang LM, Han GY (2020) Coronavirus: Govt agencies to suspend activities for seniors for 14 days to cut risk of transmission. The Straits Times URL https://www.straitstimes.com/singapore/health/govt-agencies-to-suspend-activities-for-seniors-for-14-days-starting-march-11-to, accessed April 27, 2022.

Zorlu F (2020) Thousands of citizens age 65 and above, restricted to their homes due to covid-19, take breath of fresh air. Anadolu Agency URL https://www.aa.com.tr/en/latest-on-coronavirus-outbreak/turkey-elderly-given-one-time-permission-to-go-out/1835850, accessed April 30, 2022.

